# Phase I study of a SARS-CoV-2 mRNA vaccine PTX-COVID19-B

**DOI:** 10.1101/2022.05.06.22274690

**Authors:** Natalia Martin Orozco, Noah Vale, Alan Mihic, Talya Amor, Lawrence Reiter, Yuko Arita, Reuben Samson, Queenie Hu, Anne-Claude Gingras, Brad Sorenson, Eric G. Marcusson, Piyush Patel

## Abstract

PTX-COVID19-B mRNA vaccine encodes for SARS-CoV-2 Spike protein G614 variant and lacks the proline-proline (986-987 position) mutation present in other COVID-19 vaccines. This Phase 1 observer-blinded, randomized, placebo-controlled, ascending dose study evaluated the safety, tolerability, and immunogenicity of two doses of PTX-COVID19-B vaccine in healthy seronegative adults. Participants received two intramuscular doses, 4 weeks apart, of 16-μg, 40-μg, or 100-μg PTX-COVID19-B. Adverse events were generally mild to moderate, self-resolving, and transient. The most common solicited local and systemic adverse event was pain at the injection site and headache, respectively. After the first immunization, all participants seroconverted, producing high titers of anti-receptor-binding-domain, anti-Spike, and neutralizing antibodies, including neutralizing antibodies against the ancestral viral strain and the Alpha, Beta, and Delta variants of concern, in a dose-dependent way, further increasing over 10-20 times after the second dose. All tested doses of PTX-COVID19-B were safe, well-tolerated, and provided a strong immunogenicity response. The 40-μg dose showed fewer adverse reactions than the 100-μg dose, supporting further investigation of the 40-μg dose.

**Clinical Trial Registration:** ClinicalTrials.gov identifier: NCT04765436 (https://clinicaltrials.gov/ct2/show/NCT04765436)

## INTRODUCTION

The coronavirus (severe acute respiratory syndrome coronavirus 2 [SARS-CoV-2]) world pandemic that started in 2019 continues to affect multiple countries around the world where access to vaccines is limited.^1^

To provide an alternative prophylactic vaccine against SARS-CoV-2, Providence Therapeutics Holdings, Inc. (PT) developed an mRNA vaccine, PTX-COVID19-B, composed of a lipid nanoparticle containing modified mRNA that encodes for full-length Spike (S) protein with glycine in position 614 (G614). By March of 2020, the D614G variant of SARS-CoV-2 was the predominant strain worldwide and this substitution has prevailed in current, more transmissible variants of concern (VOCs).^2,3,4^ The recently described crystal structure of the full-length G614 S trimers indicates that G614 variant forms a more stable S trimer with one receptor binding domain (RBD) remaining in the up conformation longer than D614 in the pre-fusion state. The study posited that G614 S trimer stability could provide a superior immunogen for eliciting neutralizing antibody responses, largely targeting the RBD and N-terminal domain, which stay exposed longer than with the D614 S trimer. Moreover, full-length G614 S protein vaccines could have a relative increase in potency because this change compensates for the lack of engineered stabilizing proline-proline (986-987 position) mutations present in other COVID-19 vaccines.^5,6,7,8^ Preclinical studies showed PTX-COVID19-B was safe, highly immunogenic, and completely protected animals from SARS-CoV-2 infection.^9^ Based on the preclinical results, PTX-COVID19-B was authorized by Health Canada to enter clinical trials in December 2020.

Reported here are interim findings of the Phase 1 study to evaluate the safety, tolerability, and immunogenicity of PTX-COVID19-B vaccine in healthy seronegative adults aged 18 to 64. The study began in January 2021. Enrollment was completed in April 2021. The safety database was locked, and analysis was performed on data up to day 42 after the first dose in May 2021. Immunological data is reported up to week 26 (day 180).

## RESULTS

### Study Demographics

Sixty healthy adults were randomized into three groups comprising 20 participants each (Fig. 1). In each group, 15 participants received the PTX-COVID19-B vaccine and 5 received the placebo. All 60 study participants received their first dose of vaccine on day 1 and all but 1 participant received a second dose on day 28. This participant, in the 40-μg dose group, withdrew from the study due to a non-study related consideration and was removed from final analysis.

**Figure 1:**
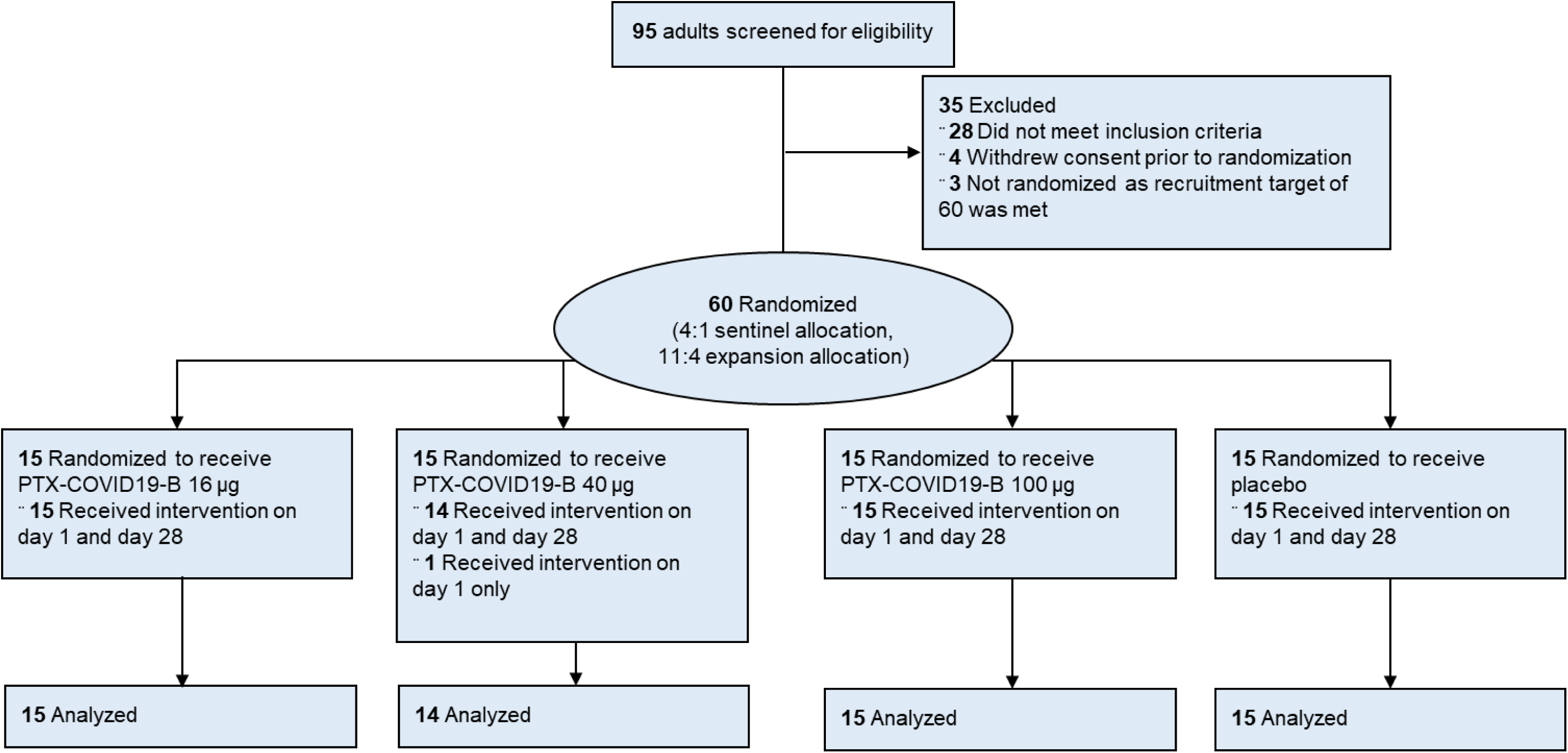
Study flow chart.

The study included 30 (50%) female and 30 (50%) male participants. The mean age was 34.9 ± 13.0 years. Key demographic characteristics of the participants at enrollment are presented in Table 1.

**Table 1.** Demographic characteristics of the participants in the Phase 1 PTX-COVID19-B trial at enrollment.

| Characteristic | PTX-COVID19-B<br>16 µg<br>(N=15) | PTX-COVID19-B<br>40 µg<br>(N=15) | PTX-COVID19-B<br>100 µg<br>(N=15) | Placebo<br>(Pooled)<br>(N=15) | Overall<br>(N=60) |
| --- | --- | --- | --- | --- | --- |
| Sex – No. (%) |  |  |  |  |  |
| Male | 7 (47) | 8 (53) | 7 (47) | 8 (53) | 30 (50) |
| Female | 8 (53) | 7 (47) | 8 (53) | 7 (47) | 30 (50) |
| Age – yr | 38.3±13.1 | 36.1±15.5 | 32.5±12.0 | 32.7±11.1 | 34.9±13.0 |
| Race or ethnic group – No.<br>(%)<sup>a</sup> |  |  |  |  |  |
| American Indian or<br>Alaskan Native | 0 | 0 | 0 | 0 | 0 |
| Asian | 1 (7) | 1 (7) | 4 (27) | 2 (13) | 8 (13) |
| Black or African<br>American | 0 | 0 | 0 | 0 | 0 |
| Native Hawaiian/Other<br>Pacific Islander | 0 | 0 | 0 | 0 | 0 |
| White | 13 (87) | 13 (87) | 11 (73) | 13 (87) | 50 (83) |
| Other | 1 (7) | 1 (7) | 0 | 0 | 2 (3) |
| Hispanic or Latino | 0 | 3 (20) | 2 (13) | 2 (13) | 7 (12) |
| Body-mass index, kg/m<sup>2b</sup> | 23.8±3.34 | 25.3±3.80 | 23.8±3.11 | 23.9±2.88 | 24.2±3.28 |

Plus-minus values are means ± standard deviation
<sup>a</sup> Race or ethnic group was reported by the participant.
<sup>b</sup> Body-mass index is the weight in kilograms divided by the square of the height in meters. This calculation was based on the weight and height measured at the time of screening.

### Safety

No serious adverse events, events leading to study discontinuation, events of special interest, new onset chronic disease, or potential immune-mediated medical conditions were reported.

After the first dose, solicited systemic and local reactions were absent or mild in most participants. Two participants (13.3%) in Cohort 2 (40-μg dose) experienced severe solicited systemic reactions (muscle pain and joint pain) and 1 participant (6.7%) in Cohort 2 experienced a severe solicited local reaction (pain) (Fig. 2).

**Figure 2.**
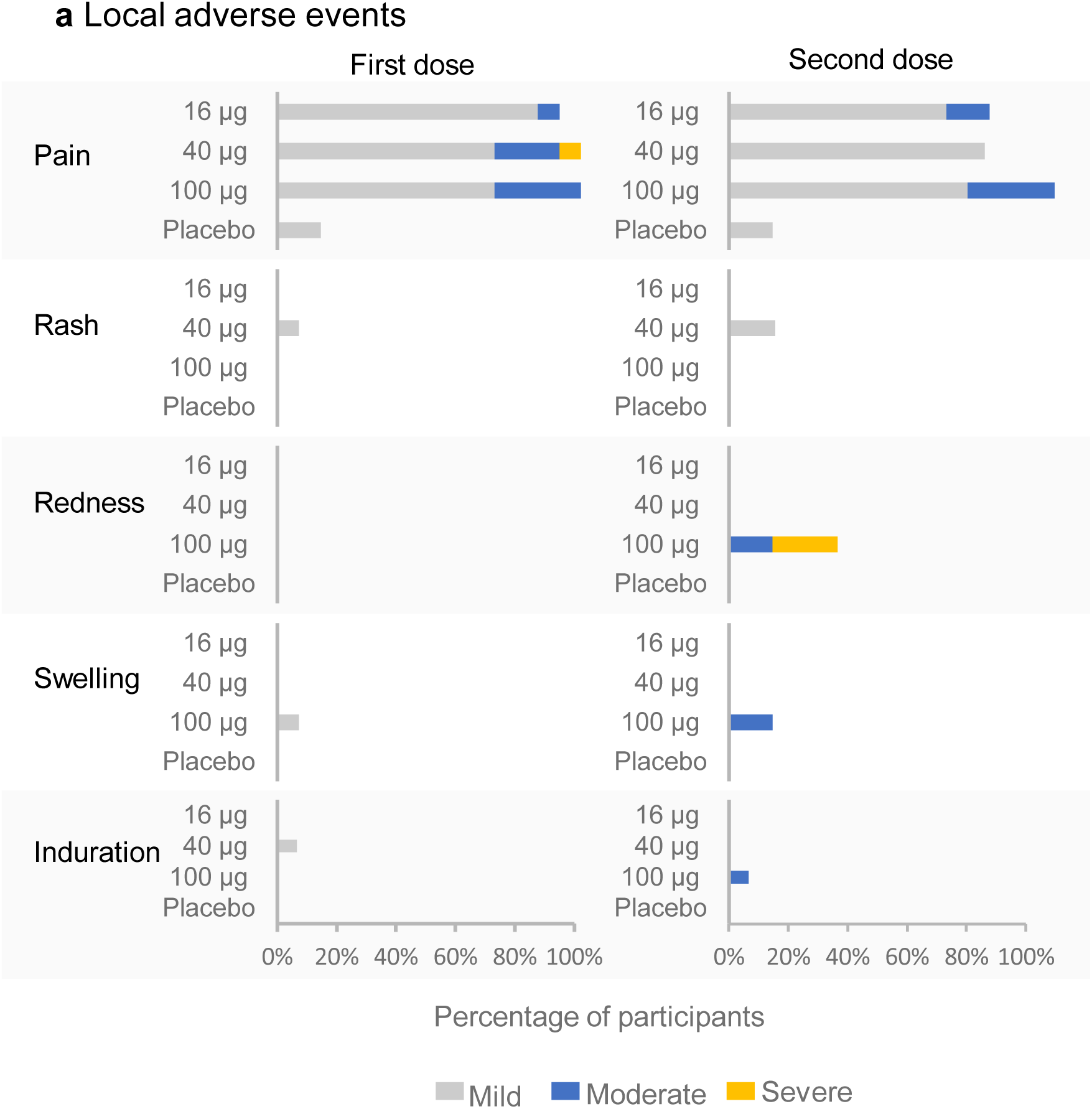
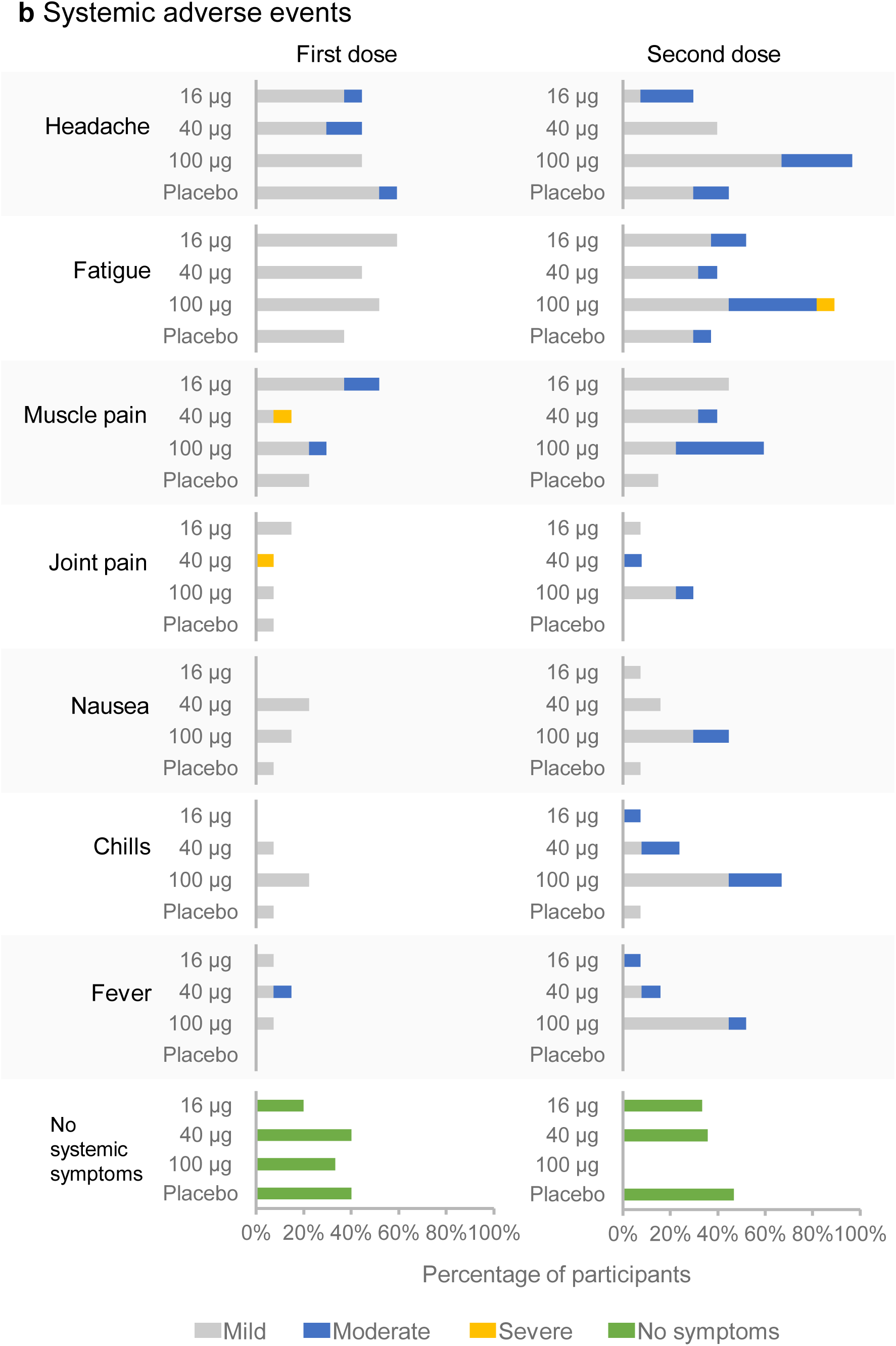
Summary of solicited systemic and local reactions. The plot represents the percentage of participants in each vaccine group (Cohort 1, 2, 3, and placebo) with local (Panel A) adverse events and solicited systemic (Panel B) adverse events through the third day post each vaccination according to FDA’s Toxicity Grading Scale (Grade 1, mild; Grade 2, moderate; Grade 3, severe; and Grade 4, potentially life threatening). There were no Grade 4 events. Participants who reported 0 events make up the remainder of the 100% calculation (not displayed). Participants that did not show any systemic symptom after vaccination are represented in the lower graph.

After the second dose, solicited systemic and local reactions were absent or mild in most participants. One participant (6.7%) in Cohort 3 (100-μg dose) experienced a severe solicited systemic reaction (fatigue) and 3 participants (20.0%) in Cohort 3 experienced severe solicited local reactions (redness). The incidence of solicited local and systemic reactions was highest at the 100-μg dose level, particularly after the second dose. Overall, the most commonly reported solicited local and systemic adverse events in vaccinated subjects were pain at the injection site and headache, respectively (Fig. 2).

The most frequently reported (>5.0% across all cohorts) unsolicited treatment-emergent adverse events, defined as adverse events which occurred or worsened after the start of study vaccination, from day 1 to day 42, in the PTX-COVID19-B groups were injection site reaction (33.3% of participants in Cohort 3), diarrhoea (13.3% and 6.7% of participants in Cohorts 2 and 3, respectively), rhinorrhea (13.3% of participants in Cohort 1), and rash (13.3% of participants in Cohort 2). Most adverse events were mild in severity and unrelated to the study drug (except injection site reaction) (Fig. 2).

Four participants had medically-attended adverse events, none of which were considered related to the study drug except severe injection site reactions in two participants in Cohort 2.

There have been no cases of COVID-19 reported up to week 26.

### Immunogenicity

#### Antibody Response

Eight days after immunization with PTX-COVID19-B there were no detectable antibodies against Spike or RBD either determined by enzyme-linked immunosorbent assay (ELISA) or Mesoscale Discovery (MSD) platform. By day 28 after the first immunization anti-S and anti-RBD immunoglobulin G (IgG) antibodies were detected across the three cohorts indicating seroconversion of all participants (Fig. 3) These antibodies were further significantly increased two weeks after the second dose of PTX-COVID19-B (day 42), in all three cohorts (*p <* 0.0001). Antibody levels were monitored up to week 26 after the first vaccination (Fig. 3B). Although the antibody levels decreased slightly by week 12 (day 84) and modestly by week 26 (day 180), they stayed above the levels observed after the first immunization (day 28). The week 26 antibody levels were also higher than COVID-19 convalescence serum except the low dose cohort (16-μg of dose, Cohort 1) (Fig. 3). Both peak antibody level and week 26 antibody level were dose dependent.

**Figure 3.**
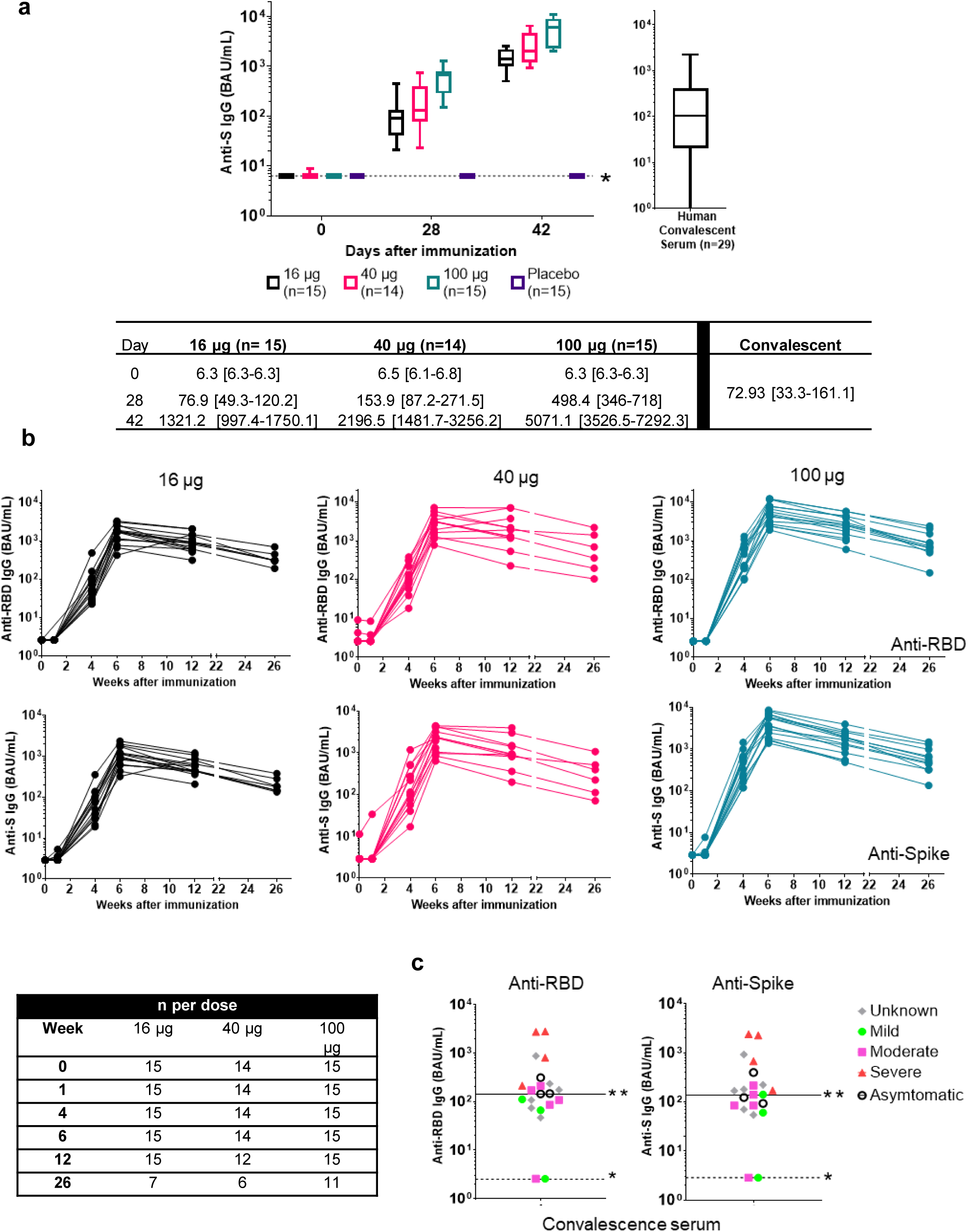
PTX-COVID19-B induction of anti-S and anti-RBD antibodies. Anti-S IgG antibody levels measured by ELISA are shown in Panel A. Anti-S antibody levels in sera from 29 subjects infected with COVID-19 were also determined by ELISA. The geometric mean and 95% confidence intervals are presented (P<0.0001). Anti-RBD and anti-S IgG antibody levels measured by MSD assay up to week 26 after immunization with PTX-COVID19-B are shown in Panel B. Control anti-RBD and anti-S antibody levels in sera from 29 subjects infected with COVID-19 are shown in Panel C. The number of subjects whose sera were tested at each timepoint are indicated in the table.

The two assays for determination of anti-Spike antibodies, ELISA and MSD, showed a strong positive correlation (correlation, 0.96 [95% confidence interval [CI] 0.95-0.98], *p <* 0.0001) (Supplementary Figure 1).

#### COVID-19 Neutralization Levels

Neutralization activity measured with a pseudovirus neutralization assay (pseudotype virus neutralization activity [pVNA] expressed in IU/mL as the assay was calibrated with the National Institute for Biological Standards and Control [NIBSC] standard) was not detected in the participants before vaccination. After the first vaccination by day 28, neutralization activity was present in all participants of the three cohorts (Fig. 4A). Moreover, after the second vaccination pVNA levels increased 16-fold for the low dose, and 30-fold for the mid and high dose. The mean neutralizing antibody titers in PTX-COVID19-B vaccinated groups were at least 8-fold higher at all doses compared to control serum samples from convalescent patients (HCS) (Fig. 4A). The anti-S and anti-RBD antibody levels correlate with the neutralization units in all three assays (Supplementary Figure 2). Also, the determination of angiotensin-converting enzyme 2 (ACE2):S blocking antibodies by MSD assay correlates with the neutralization units (Supplementary Figure 3).

**Figure 4.**
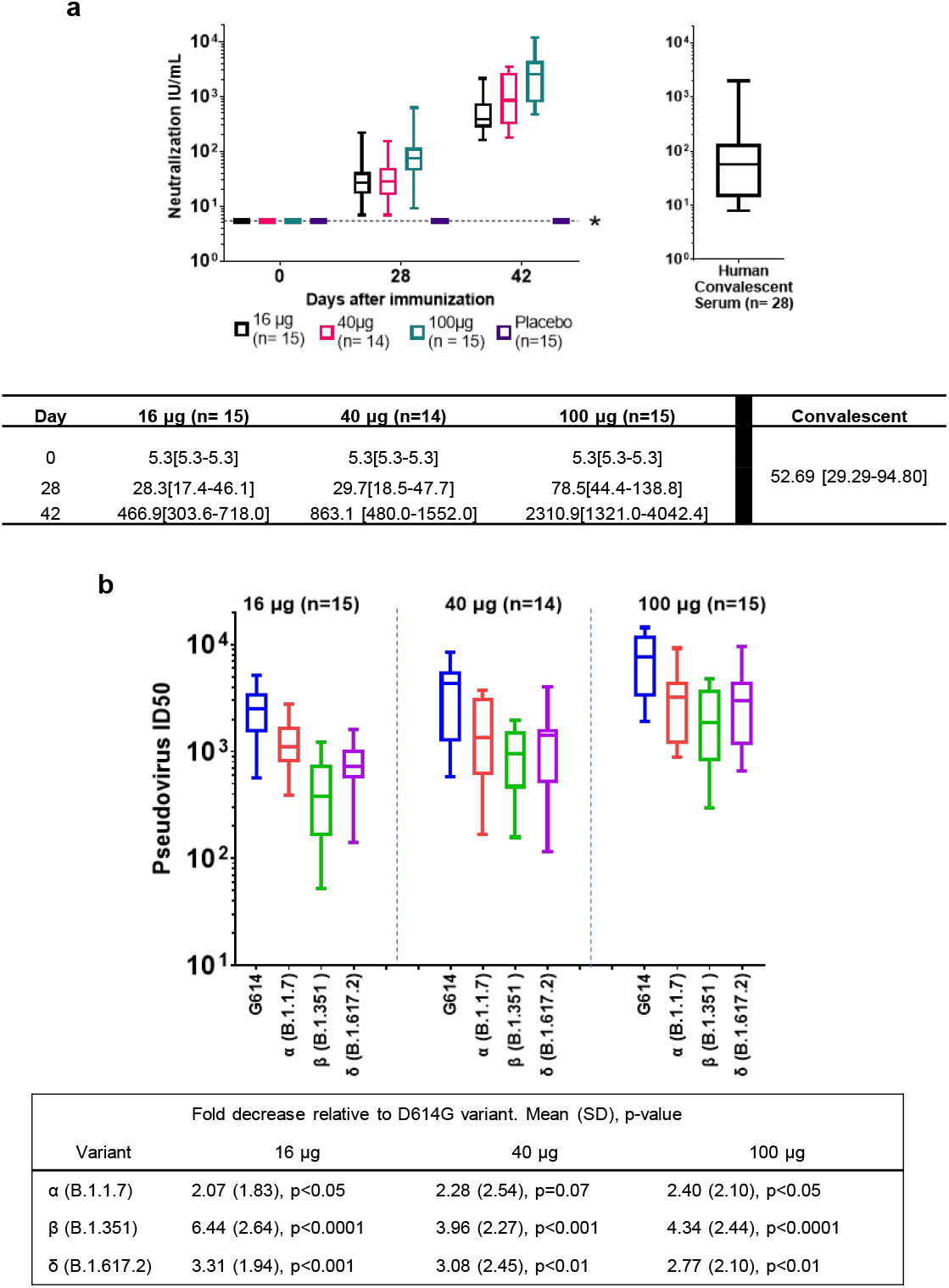
PTX-COVID19-B vaccine induction of neutralizing antibodies against G614 SARS-CoV-2 and VOCs. Levels of neutralization (IU/mL) of sera from PTX-COVID19-B subjects from day 42 against G614 are shown in Panel A. Levels of neutralization from sera from 28 subjects infected with COVID-19 were also determined. Levels of neutralization (ID50) of sera from PTX-COVID19-B subjects from day 42 against VOCs Alpha, Beta, and Delta are shown in Panel B. Mean (SD) fold decrease relative to D614G variant are shown in the table, where values were calculated by dividing the average ID50 value of G614G by ID50 concentration of other variants indicated.

Levels of neutralization against wild-type D614G and the Alpha (B.1.1.7), Beta (1.351), and Delta (B.1.617.2) VOCs were evaluated at day 42 for all three cohorts (Fig. 4B) using a pseudovirus neutralization assay. However, since there is no standard available for VOCs, the results are expressed as ID50 (Inhibitory dilution 50%). All participants vaccinated with PTX-COVID-19-B produced neutralizing antibodies against the VOCs tested. As the vaccine dose increased, the ID50 against the VOCs also increased. While the neutralization response was lower for the VOCs compared to the G614 strain, especially for VOC Beta, all three doses of PTX-COVID-19-B can elicit a strong neutralizing antibody response against the VOCs with nominal titers comparable to the current approved mRNA vaccines.^10,11^

The magnitude of neutralizing antibody determined by the lentivirus assay and the recombinant Vesicular Stomatitis Virus (rVSV) neutralization assays correlated well (correlation: 0.87 [95% CI 0.80-0.91] *p <* 0.0001) for all samples from the three dose levels at day 28 and day 42 (Supplementary Figure 4). The results indicate comparability of both assays for the interpretation of neutralization activity.

## DISCUSSION

Vaccination with PTX-COVID19-B in 18-to 64-year-old participants evaluated in the Phase 1 trial indicates a safe profile. Local and systemic reactions were mild after the first vaccine dose and increased to moderate after the second dose only in a minority of the participants. So far PTX-COVID19-B vaccination has shown fewer reactions than other approved COVID-19 vaccines.^7,8,12^ This improved safety needs to be confirmed in follow-up clinical trials with a large number of participants.

Neutralizing antibody was observed in 100% of the vaccinated participants after the first immunization at day 28 regardless of the dose level, higher than the conversion rate of neutralizing antibody observed for Moderna and Pfizer’s mRNA vaccines in their phase 1/2 clinical trial.^8,13^ This early induction of neutralizing antibody may be beneficial for vaccinees since it may provide early protection against SARS-CoV-2 after vaccination. At around 6 months (day 180) after vaccination, antibody levels for the 40 μg and 100 μg dose cohorts were higher than COVID-19 convalescence serum, suggesting a more durable protection than current mRNA vaccines. Moreover, the levels of neutralizing antibodies induced by PTX-COVID19-B against the ancestral strain and VOCs are comparable to other available COVID-19 mRNA vaccines and higher than the levels considered protective,^14,15^ which suggest the efficacy of PTX-COVID19-B equivalent to the current approved COVID-19 mRNA vaccines.^8,10,16^ All these potential advantages of PTX-COVID19-B need to be further extended in follow-up clinical trials.

The safety, tolerability, and immunogenicity, including neutralizing antibody response to VOCs, of the 40-μg dose support the selection of this dose for evaluation in Phase 2 clinical trials.

Critical limitations of this trial include a small study population and limited ethnic diversity as compared with the general population. Additionally, as a Phase 1 safety trial, the study population did not include those at increased risk of severe illness from COVID-19, including older adults and immunocompromised individuals that may mount a weaker immune response to vaccination.

Although several COVID-19 vaccines have been approved globally (most under emergency or interim use), issues still remain with supply and delivery of vaccines to combat emerging variants. Additional safe, effective, and easily deployable SARS-CoV-2 vaccines are needed to meet the challenge for global immunization required to end the pandemic. Based on the reported results, PTX-COVID19-B vaccine is a promising candidate that warrants testing in the next phase of trials. A Phase 2 trial of PTX-COVID19-B in 525 healthy adults with the 40-μg dose is ongoing. A large Phase 3 trial will follow in 2022. The vaccine will be also tested as a 3^rd^ dose booster in a Phase 2/3 clinical trial.

## METHODS

### Trial Design

This observer-blinded, randomized, placebo-controlled, ascending dose Phase 1 study is being conducted at one site in Canada (Manna Research, Toronto, Ontario). Eligible participants included healthy men and nonpregnant women between 18 to 64 years of age, with a body mass index of 18 to 30 kg/m^2^ at screening. All participants were seronegative to SARS-CoV-2 and reverse transcription-polymerase chain reaction (RT-PCR)-negative without evidence of recent exposure to SARS-CoV-2 or viral respiratory diseases.

Two doses of 16-μg (Cohort 1), 40-μg (Cohort 2), and 100-μg (Cohort 3) PTX-COVID19-B were compared to placebo with 20 participants in each cohort. Vaccine or placebo were administered on days 1 and 28. The sentinel group had five participants in each of the dose cohorts, who were randomized in a 4:1 ratio (vaccine: placebo). After review of the safety data of the sentinel group, the trial was followed by a cohort expansion of 15 participants randomized in a 11:4 ratio (vaccine:placebo). Randomization codes were generated using a computer program and provided as a paper listing of vaccine or placebo assignment by an unblinded statistician. The pharmacist and staff members that prepared and administered the doses based on the paper randomization were unblinded; all others involved in the conduct of the study (Principal Investigator, site coordinator, site staff) and the participants were blinded.

PT is the sponsor of the study, which is being conducted under a protocol approved by Advarra. This study is conducted in accordance with the provisions of the Declaration of Helsinki, and in accordance with the International Council for Harmonisation of Technical Requirements for Pharmaceuticals for Human Use E6 Guidelines on Good Clinical Practice (GCP). All participants provided written informed consent prior to enrollment. Full text of the trial protocol, including detailed statistical analysis plan are provided in the Supplementary Materials.

### Trial Vaccine and Placebo

PTX-COVID19-B mRNA vaccine, manufactured by PT (lot number: PTX-CB-003), was supplied as a sterile solution (0.2 mg/mL). The mRNA encoding for 1273 amino acid of SARS-COV-2 whole length S protein was formulated in lipid nanoparticles as the final drug product. The placebo was injectable saline solution (sodium chloride 0.9%). Vaccine doses were diluted with injectable saline to reach the 16-μg and 40-μg doses. The 100-μg doses did not require dilution. Vaccine and placebo were administered intramuscularly into the upper arm deltoid muscle of the non-dominant side in a 0.5 mL dose with a 25g needle.

### Safety and Reactogenicity Assessments

Participants were monitored for solicited and unsolicited adverse events (see Supplementary Figure 5 for the schedule of key assessments). The investigator assessed each event for seriousness, intensity, and relationship to the vaccine. Vital signs and site reactions were monitored for an hour after each vaccination. Participants maintained a Diary Card to track reactogenicity and other safety assessments for 7 days after each vaccination. Reactogenicity solicited events included systemic symptoms including headache, fatigue, muscle pain, joint pain, nausea, chills, and fever, and localized reactions including injection site pain, bruising, redness, and swelling. Participants were asked to record oral temperature with their own thermometer if they felt any of the above symptoms and record it in the Diary Card.

Additionally, the overall safety was analyzed, including unsolicited adverse events, medically attended adverse events, new onset chronic disease, serious adverse events, adverse events of special interest, potential immune-mediated medical conditions, and findings from targeted physical examinations, vital signs assessments, and clinical safety laboratory testing. The intensity of a solicited adverse event was graded according to United States Food and Drug Administration (FDA) standards (see Supplementary Table 1).^17^ For the adverse events not covered by the FDA standard, the rating scale from the Division of AIDS (DAIDS) was used (see Supplementary Table 2).^18^ Standard laboratory and vital signs reference ranges were used and assessed for clinical significance by the Investigators.

Safety reviews were performed by the independent Safety Review Committee according to the committee charter.

### Immunogenicity Assessments

#### Anti-S and Anti-RBD MSD Assay

The responses induced by PTX-COVID19-B on anti-COVID-19 S-protein IgG and anti-COVID-19 RBD IgG were analyzed from serum samples using a multiplex immunoassay sandwich-based method with an electrochemiluminescent (ECL) readout on the MSD platform. Diluted samples as well as calibrators and controls were added to MSD plates (multi-spot wells) coated with either the S or RBD SARS-CoV-2 antigens to capture human IgG antibodies present in the samples. Bound IgG antibodies were then detected using sulfo-TAG labeled anti-human IgG antibody. The human IgG antibody concentration was determined by interpolation of the ECL count on the calibration curve. The assay was validated and performed under GLP conditions at CIRION (Laval, Quebec; V-PLEX^®^ SARS-CoV-2 Panel 2 Kit. Report CIR21417.A01). Analysts knew timepoints but not dose levels or regimen of the samples. To convert the results from MSD arbitrary units (AU/mL) to the World Health Organization (WHO) International Standard in binding antibody units (BAU)/mL, a conversion factor of 0.0272 was applied for anti-RBD IgG concentrations and 0.00901 for anti-S IgG concentrations.

#### Anti-S ELISA

An indirect binding ELISA was also performed to determine if the elicited antibodies could bind the S protein. The SARS-CoV-2 pre-fusion S antigen was adsorbed onto a 96-well microplate and standard ELISA procedure was followed with anti-SARS-CoV-2 S IgG specific antibody (primary antibody) and anti-human IgG antibody (secondary antibody) conjugated to peroxidase. The absorbance of each well was measured using a spectrophotometer at a specific wavelength (450/620 nm). A standard on each tested plate was used to calculate the antibodies against SARS-CoV-2 S according to the unit assigned by the standard (ELU/mL). The assay was validated and performed under GLP conditions at Nexelis (Laval, Quebec; Report number 001_ICA_HU_SARX-CoV-2PRE-SPIKE_ELI_SER_AVR). To convert the results to the WHO International Standard in BAU/mL, a conversion factor of 1/7.9815 was applied. Additionally, anti-S IgG ELISA measurements for 29 control serum samples from convalescent patients with a documented positive SARS-CoV-2 RNA diagnosis (HCS) were also performed.

#### Pseudovirus Neutralization Assay

To evaluate the neutralizing effect of the sera from participants treated with PTX-COVID19-B, a pseudotyped virus neutralization assay developed and validated at Nexelis, Canada^19^ to quantify the titer of neutralizing antibody against SARS-CoV-2. The assay used rVSV particles containing modified S protein of SARS-CoV-2 (Wuhan strain) for which the last 19 amino acids of the cytoplasmic tail were removed. The pseudoparticles contained a luciferase reporter for quantification in relative luminescence units (RLU). The dilution of plasma required to achieve 50% neutralization (NT50) when compared to the pseudoparticle control was interpolated from a linear regression using the two dilutions flanking the 50% neutralization. To convert the NT50 titer to the WHO International Standard in IU/mL, a conversion factor of 1/1.872 was applied. Additionally, neutralizing antibodies for 28 control samples from HCS were assessed in the pseudovirus (rVSV) neutralization assay.

#### Pseudovirus Neutralization Assay

To determine neutralization activity against SARS-CoV-2 Wuhan-Hu1 (D614G) and the VOCs, a pseudovirus neutralization assay was performed using a SARS-CoV-2 S protein pseudotyped lentivirus at the Lunenfeld-Tanenbaum Research Institute at Mount Sinai Hospital (Toronto, Ontario). Spike-pseudotyped lentiviral assays were performed as previously described with minor modifications.^9,16^ The pseudotyped virus used in this assay is a lentivirus expressing the S protein of SARS-CoV-2 Wuhan-Hu1 (D614G), or the Alpha (B.1.1.7), Beta (B.1.351), or Delta (B.617.2) variants. Spike-pseudotyped lentivirus particles were generated and used at a virus dilution resulting in >1000 RLU over control. For the neutralization assay, human sera were serially diluted and incubated with diluted pseudovirus at a 1:1 ratio for 1 hour at 37°C followed by transfer to plated HEK293T-ACE2/TMPRSS2 cells and incubated for 48 hours at 37°C and 5% CO2. Cells were then lysed, and Bright Glo luciferase reagent (Promega, Madison, WI) was added for 2 minutes prior to reading with a PerkinElmer Envision instrument (PerkinElmer, Waltham, MA). The ID50 were calculated in GraphPad Prism 9 using a nonlinear regression algorithm (log[inhibitor] versus normalized response – variable slope). The assay was performed in the same manner for all VOCs tested.

#### ACE2:S Neutralization Assay

The neutralizing antibody response was analyzed at days 8, 28, and 42 using a S-ACE2:S blocking assay which has an ECL readout on the MSD platform. The assay measures antibodies that block the binding ofACE2 to the SARS-CoV-2 S protein and RBD antigens. Blocking antibodies bind to S protein in the plate and prevent the binding of ACE2 labeled with MSD-Sulfotag. A reference standard, included in the kit, was used to generate a standard curve. Neutralizing antibody concentrations in samples were calculated by backfitting the measured signals for samples to the standard curve. Correcting for dilution provided the final neutralization antibody concentration in undiluted samples. In this assay, 1 unit/mL corresponded to neutralizing activity of 1000 ng/mL of antibody that can bind to S protein.

### Statistical Analysis

The statistical analysis was conducted using SAS^®^ version 9.04 (SAS Institute, Cary, NC). No formal sample size calculation was performed.

The database was locked and unblinded five weeks after the last participant received their last dose. The data presented here are up to 6 months (day 180) for anti-RBD and anti-S IgG antibody levels by MSD and up to day 42 for all other safety and immunogenicity analyses, two weeks after the last participant received their last dose.

Immunogenicity data were summarized by cohort and time point using descriptive statistics along with geometric mean and 95% CI. Results were analyzed using a mixed effects model with treatment as a fixed effect and participant as a random effect for each visit. Placebo participants were pooled for comparison. ANOVA with Tukey’s multiple comparison was performed (F [3, 55] = 26.90, degrees of freedom = 3). Analysis was performed at a 5% level of significance. The correlation coefficients were calculated using a two-tail Pearson correlation with 95% confidence.

The number and percentage of participants reporting any treatment-emergent adverse event or reactogenicity were summarized by cohort and tabulated by system organ class and preferred term (coded using MedDRA, WHO Global B3 format – MAR 2021).

## DATA AVAILABILITY

The datasets generated during and/or analysed during the current study are available from the corresponding author on reasonable request.

## Supporting information

Clinical Protocol

Statistical Analysis Plan

## Data Availability

All data produced in the present work are contained in the manuscript.The development of the B.1.617.2 (Delta) lentiviral assay was supported by the CIHR operating grant to the Coronavirus Variants Rapid Response Network (CoVaRR-Net) to A.-C.G. A.-C.G. is the Canada Research Chair, Tier 1, in Functional Proteomics.

## AUTHOR CONTRIBUTIONS

N.M.O. analyzed the data, prepared the figures, and wrote the manuscript; N.V., A.M., and T.A. provided clinical assessment, clinical trial coordination, revision of the data, and revision of the manuscript; L.R. provided revision of the data and revision of the manuscript; Y.A., R.S., Q.H., and A.-C.G. created the pseudovirus, performed pseudovirus testing, analyzed the data, and reviewed the manuscript; B.S. and E.G.M. provided the clinical trial design and revision of the manuscript; P.P. provided the clinical trial design, revision of the manuscript, and clinical trial coordination.

## ACKNOWLEDGEMENTS

We thank the PT manufacturing team for their contributions that lead to this Phase 1 trial. We thank Sunnybrook Research Institute for the GMP operational support (see Supplementary Materials). We thank Dr. W Rod Hardy for the Delta (B.1.617.2) spike construct, and Karen Colwill and Bhavisha Rathod for assistance with logistics. A medical writer employed by PT provided the first draft of the manuscript and editorial assistance.

## COMPETING INTERESTS

N.M.O., L.W., Y.A., B.S., E.G.M., and P.P. are employees of PT. PT received funding for the Phase 1 trial from National Research Council Canada. The development of the B.1.617.2 (Delta) lentiviral assay was supported by the CIHR operating grant to the Coronavirus Variants Rapid Response Network (CoVaRR-Net) to A.-C.G. A.-C.G. is the Canada Research Chair, Tier 1, in Functional Proteomics.

## Notes

### Clinical Trial

NCT04765436

### Funding Statement

Providence Therapeutics received funding for the Phase 1 trial from National Research Council Canada.

### Author Declarations

The trial got full ethics approval from Advarra 372 Hollandview Trail, Suite 300 Aurora, ON L4G 0A5 Canada IORG Number: 0000635 IRB Registration: 00000971 FWA Number: 00023875

