## Supplementary material for "Phase I study of a SARS-CoV-2 mRNA vaccine PTX-COVID19-B"

### Supplementary Materials

**PROVIDENCE THERAPEUTICS TEAM**
Nathalie Fabbri, Florian Gossart, Karitza Fuenmayor, Rajesh Krishnan, Aula Al Muslim, Richard Tran, Joseph Bteich, Aisha Yusuf, Priya Rajesh, Jordan Schwartz, Jared Davis

**SUNNYBROOK RESEARCH INSTITUTE TEAM**
Nickett Donaldson, Crystal Bourne, and Rod Engeland

#### SUPPLEMENTARY FIGURES

##### Supplementary Figure 1. Correlation between ELISA and MSD assays for detection of anti-S antibodies

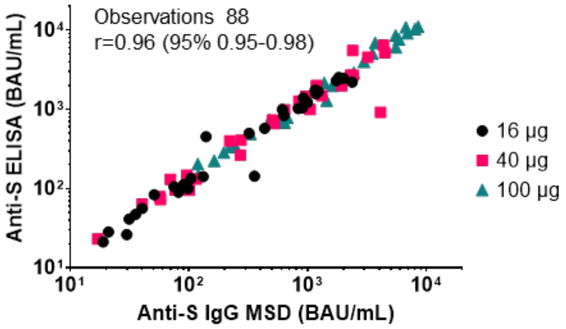


Detection of anti-S antibodies from participants vaccinated with PTX-COVID19 was determined by ELISA and multiplex MSD. Both determinations were calibrated with the WHO standard and results from both assays have a very good correlation (r=0.96).

##### Supplementary Figure 2. Correlation of anti-S IgG and neutralizing antibody response

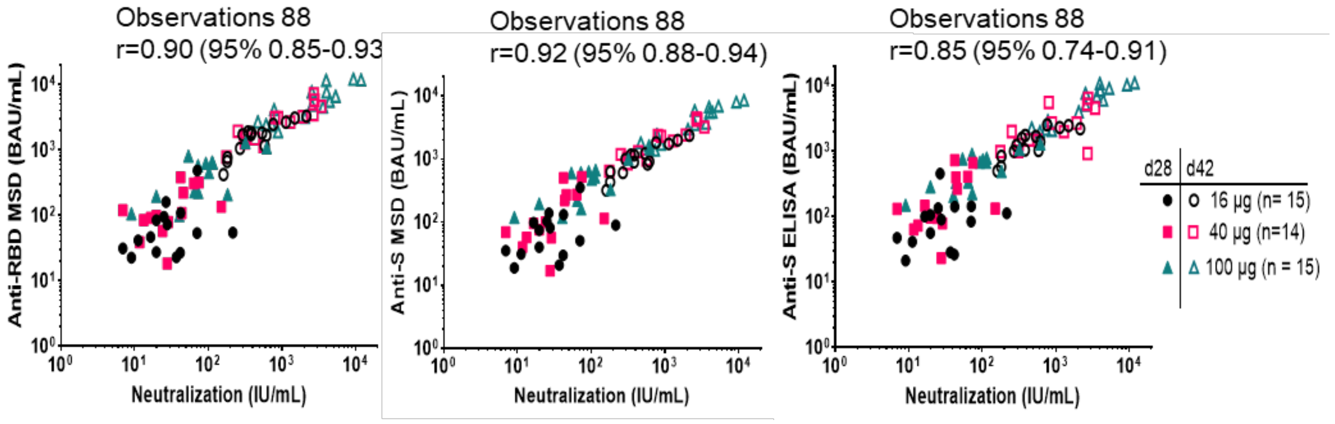

**Supplementary Figure 3. Correlation of anti-Spike IgG and neutralizing antibody response**

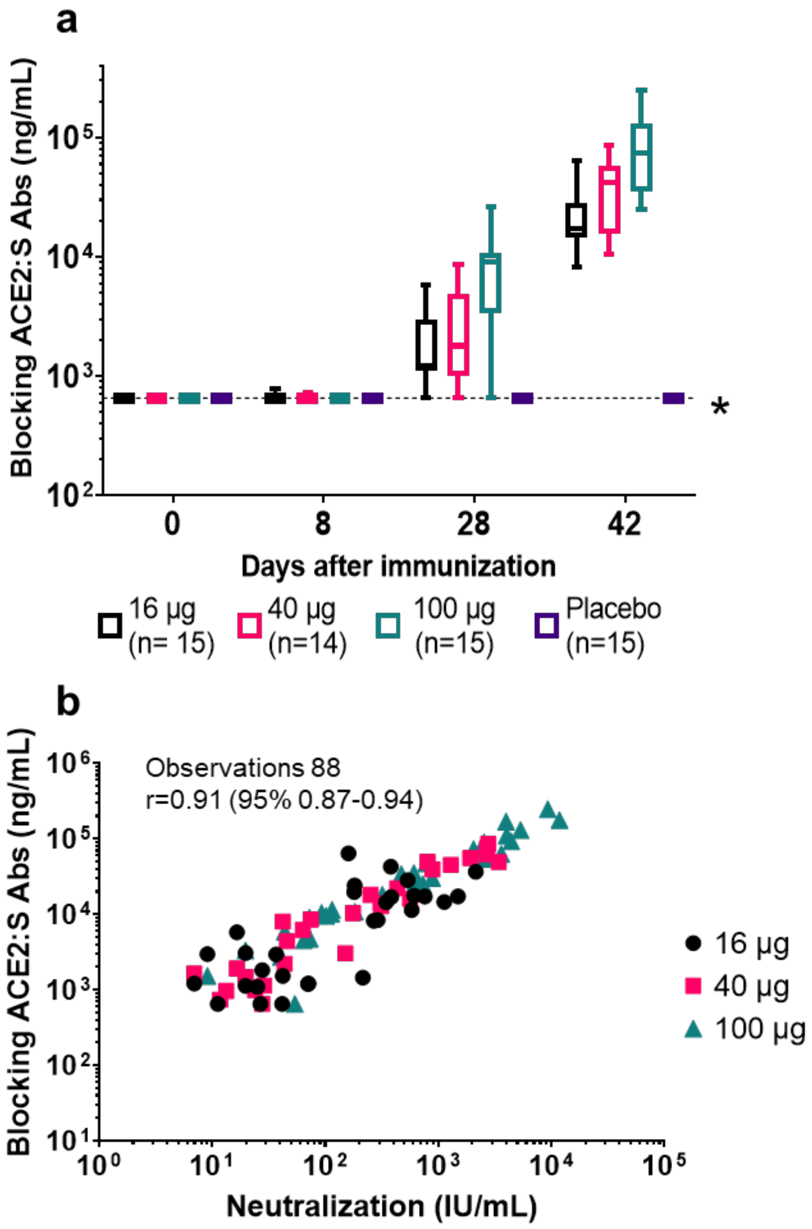


A. Detection of neutralizing activity from the sera of participants vaccinated with PTX-COVID-19-B was determined by ACE2:S blocking assay. Neutralization activity was evident at day 28 after the first immunization and increased by day 42 in all cohorts.
B. The neutralization activity determined by ACE2:S blocking assay correlates to the neutralization activity determined pVNA assay.

**Supplementary Figure 4. Correlation of pseudovirus neutralization assays, lentivirus vs. rVSV**

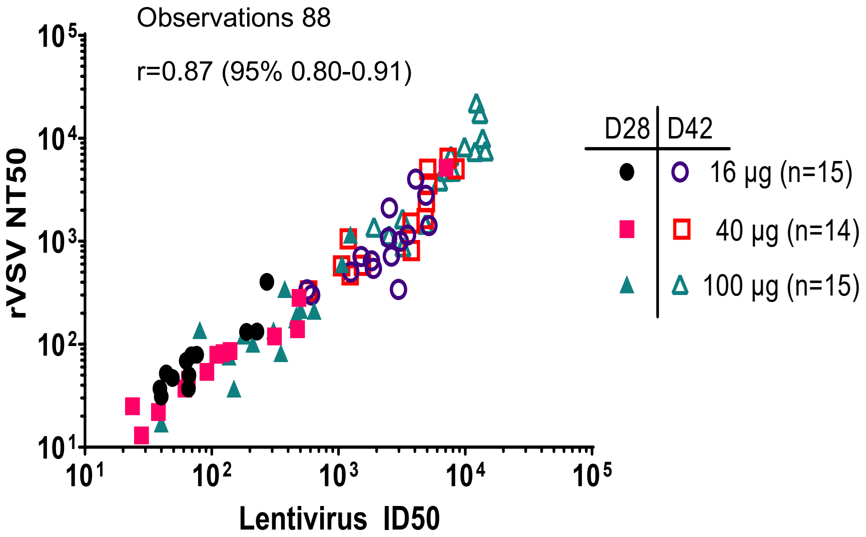


Detection of neutralization activity pVNA from participants vaccinated with PTX-COVID19 was determined by two methods, one method used rVSV expressing ancestral S protein and was performed at Nexelis; the second method used lentivirus expressing G614 S variant and was performed at Mount Sinai Hospital. Both assays have a very good correlation (r=0.87).

#### Supplementary Figure 5. Vaccine Regimens and Schedule of Key Trial Assessments

**A** **Vaccine Regimens**

| **Vaccine Groups** | **No. of Participants** | | **Day 1** | **Day 28** |
|---|---|---|---|---|
| | Randomized (PTX-COVID19-B: placebo) | Randomized Sentinel (PTX-COVID19-B: placebo) | PTX-COVID19-B | PTX-COVID19-B |
| Cohort 1 | 11:4 | 4:1 | 16 µg or placebo | 16 µg or placebo |
| Cohort 2 | 11:4 | 4:1 | 40 µg or placebo | 40 µg or placebo |
| Cohort 3 | 11:4 | 4:1 | 100 µg or placebo | 100 µg or placebo |

**B** **Key Trial Assessments**

| **Procedure** | **Screening** | **Day** 1 | 2 | 8 | 28 | 29 | 42 | 90 | 180 | 395 |
|---|---|---|---|---|---|---|---|---|---|---|
| NP swab for COVID-19 via RT-PCR | X | | | | X | | | | | |
| Blood samples for immunogenicity analysis (antibodies IgG, IgA, neutralization) | X | | | X | X | | X | X | X | X |
| Vaccine administration | | X | | | X | | | | | |
| Post-vaccination assessment (approximately 1 hour post vaccination, includes arm check and vital signs) | | X | | | X | | | | | |
| Medically attended AE assessments | | ← | | | | | → | | | |
| Unsolicited AE assessments | | ← | | | | | → | | | |
| Solicited AEs | | X | X | | X | X | | | | |
| Assessments of NOCD, AESIs (including COVID-19 cases for enhanced disease), and PIMMCs | | ← | | | | | | | | → |
| Serious adverse event assessments | ← | | | | | | | | | → |

Shown are the planned randomization and associated vaccine regimens administered in the trial (Panel A), along with the timing of key trial assessments (Panel B).

#### SUPPLEMENTARY TABLES

##### Table 1. FDA Toxicity Grading Scale for Solicited Local and Systemic Reactions

| | Mild (Grade 1) | Moderate (Grade 2) | Severe (Grade 3) | Potentially Life Threatening (Grade 4) |
|---|---|---|---|---|
| **Local Reaction to Injectable Product** | | | | |
| Pain | Does not interfere with activity | Repeated use of non-narcotic pain reliever > 24 hours or interferes with activity | Any use of narcotic pain reliever or prevents daily activity | Emergency room (ER) visit or hospitalization |
| Erythema/ Redness[a] | 2.5 - 5 cm | 5.1 - 10 cm | > 10 cm | Necrosis or exfoliative dermatitis |
| Induration[b] | 2.5 - 5 cm and does not interfere with activity | 5.1 - 10 cm or interferes with activity | > 10 cm or prevents daily activity | Necrosis |
| Swelling[b] | 2.5 - 5 cm and does not interfere with activity | 5.1 - 10 cm or interferes with activity | > 10 cm or prevents daily activity | Necrosis |
| **Systemic (General)** | | | | |
| Fever[c] (°C)<br>(°F) | 38.0 – 38.4<br>100.4 – 101.1 | 38.5 – 38.9<br>101.2 – 102.0 | 39.0 – 40<br>102.1 – 104 | > 40<br>> 104 |
| Nausea | No interference with activity or 1 – 2 episodes/24 hours | Some interference with activity or > 2 episodes/24 hours | Prevents daily activity, requires outpatient IV hydration | ER visit or hospitalization for hypotensive shock |
| Headache | No interference with activity | Repeated use of nonnarcotic pain reliever > 24 hours or some interference with activity | Significant; any use of narcotic pain reliever or prevents daily activity | ER visit or hospitalization |
| Fatigue | No interference with activity | Some interference with activity | Significant; prevents daily activity | ER visit or hospitalization |
| Muscle pain | No interference with activity | Some interference with activity | Significant; prevents daily activity | ER visit or hospitalization |
| Joint pain | No interference with activity | Some interference with activity | Significant; prevents daily activity | ER visit or hospitalization |
| Chills | No interference with activity | Some interference with activity | Significant; prevents daily activity | ER visit or hospitalization |
| Feverish | No interference with activity | Some interference with activity | Significant; prevents daily activity | ER visit or hospitalization |
| Rash (not at injection site) | No interference with activity | Some interference with activity | Significant; prevents daily activity | ER visit or hospitalization |

[a] In addition to grading the measured local reaction at the greatest single diameter, the measurement should be recorded as a continuous variable.
[b] Induration/Swelling should be evaluated and graded using the functional scale as well as the actual measurement.
[c] Oral temperature; no recent hot or cold beverages or smoking.
The intensity of a solicited adverse event was graded according to the FDA Guidance for Industry: Toxicity Grading Scale for Healthy Adult and Adolescent Volunteers Enrolled in Preventive Vaccine Clinical Trials (September 2007). The grading scale comprises 4 levels: mild (Grade 1), moderate (Grade 2), severe (Grade 3), and potentially life-threatening (Grade 4). The scale was adapted for the protocol.
Reference: Food and Drug Administration. Guidance for industry: Toxicity grading scale for healthy adult and adolescent volunteers enrolled in preventive vaccine clinical trials. US Department of Health and Human Services, Food and Drug Administration, Center for Biologics Evaluation and Research. September 2007. https://www.fda.gov/media/73679/download

**Table 2. DAIDS Adverse Events Intensity Assessment**

| Grade | Intensity | Definition |
|---|---|---|
| Grade 1 | Mild | Symptoms causing no or minimal interference with usual social and functional activities with intervention not indicated. |
| Grade 2 | Moderate | Symptoms causing greater than minimal interference with usual social and functional activities with intervention indicated. |
| Grade 3 | Severe | Symptoms causing inability to perform usual social and functional activities with intervention or hospitalization indicated. |
| Grade 4 | Potentially Life-Threatening | Symptoms causing inability to perform basic self-care functions with intervention indicated to prevent permanent impairment, persistent disability, or death. |
| Grade 5 | | Any adverse event where the outcome is death |

For those adverse events that are not covered in the FDA Guidance for Industry (see Table 1) the rating scale from DAIDS (Table for Grading the Severity of Adult and Pediatric Adverse Events, Version 2.1) was used.
Reference: Division of AIDS (DAIDS) Table for Grading the Severity of Adult and Pediatric Adverse Events. Corrected Version 2.1. Division of AIDS, National Institute of Allergy and Infectious Diseases, National Institutes of Health, US Department of Health and Human Services. July 2017.
https://rsc.niaid.nih.gov/sites/default/files/daidsgradingcorrectedv21.pdf
