## Supplementary material for "Phase I study of a SARS-CoV-2 mRNA vaccine PTX-COVID19-B": Clinical Protocol

### Clinical Study Protocol

### A Phase I, First-in-Human, Observer-Blinded, Randomized, Placebo Controlled, Ascending Dose Study to Evaluate the Safety, Tolerability, and Immunogenicity of PTX-COVID19-B Vaccine in Healthy Seronegative Adults Aged 18-64

Protocol Number: PRO-CL-001

Version 5.0

Date: 14 February 2021

| | |
|---|---|
| Sponsor: | Providence Therapeutics Holdings Inc.<br>661 University Avenue, Suite 1300<br>Toronto, ON M5G 0B7 Canada<br>Telephone: 416-597-3486 |
| Contract Research Organization: | ICON Clinical Research, LLC<br>820 West Diamond Avenue, Suite 100<br>Gaithersburg, MD 20878 USA<br>Telephone: (301) 944-6800<br>Fax: (215) 699-6288 |

**Confidentiality Statement**

This document and the information contained herein or attached hereto ("Confidential Material") are confidential and proprietary to Providence Therapeutics Holdings Inc. This Confidential Material should be viewed only by those individuals or companies that have been given prior authorization to do so by Providence Therapeutics Holdings Inc. ("Authorized Users"). This Confidential Material should not be made available in any form to any person or company, other than the Authorized Users and their respective employees or associates on a need-to-know basis, without the prior written consent from Providence Therapeutics Holdings Inc.

### 1 SYNOPSIS

| | |
|---|---|
| **Protocol Title:** | A Phase I First-in-Human, Observer-Blinded, Randomized, Placebo-Controlled, Ascending Dose Study to Evaluate the Safety, Tolerability, and Immunogenicity of PTX-COVID19-B Vaccine in Healthy Seronegative Adults Aged 18-64 |
| **Study Phase:** | Phase I |
| **Objectives:** | • To evaluate the safety and tolerability of 2 doses of PTX-COVID19-B vaccine in healthy seronegative adults 18-64 years of age<br>• To evaluate the immunogenicity of 2 doses of PTX-COVID19-B vaccine in healthy seronegative adults 18-64 years of age |
| **Endpoints:** | Safety and tolerability and reactogenicity:<br>• Occurrence of events during the follow-up after each vaccination will be analyzed using both the Per Protocol (PP) and Safety Populations:<br>o Vital signs and administration site reactions (eg, arm check evaluations including pain, tenderness, erythema/redness, induration/swelling) during the follow-up after each vaccination<br>o Daily solicited adverse events (AEs; eg, fever, chills, nausea/vomiting, diarrhea, headache, fatigue, myalgia) through the second day post each vaccination.<br>• Overall safety will be analyzed using both the Safety and Per-Protocol Populations.<br>o Unsolicited adverse events from Day 1 through Day 42<br>o Medically attended AEs (Day 1 through Day 42), new onset chronic disease (NOCD), serious adverse events [SAEs], adverse events of special interest [AESIs], and potential immunemediated medical conditions (PIMMCs) from Day 1 through to Day 395 (approximately 1 year after the last vaccination)<br>o Findings from targeted physical examinations, vital sign assessments, and clinical safety laboratory testing<br>Immune Response:<br>• Immunogenicity analysis using the Safety, Per Protocol, and mITT Populations. Analysis using the mITT Population will not be performed if it differs from the PP Population by ≤ 5% of the subjects for each of the treatment groups:<br>o Antibodies (anti-COVID-19 immunoglobulin (Ig), IgG, IgA [enzyme-linked immunosorbent assay]; and anti-COVID-19 neutralization titer assays)<br>o Cell-mediated immunity using blood/peripheral blood mononuclear cells (PBMCs [Flow Cytometry Assay]) |
| **Study Design:** | This is a Phase I, first-in-human, observer-blinded, randomized, placebo -controlled, ascending dose study to evaluate the safety, tolerability, and immunogenicity of PTX-COVID19-B vaccine in healthy seronegative adults aged 18 to -64. The study is designed with dose-escalations and will be performed in seronegative adult subjects without evidence of recent of exposure to Severe Acute Respiratory Syndrome (SARS)-CoV-2 or viral respiratory disease not identified as influenza or respiratory syncytial virus (RSV) (febrile or lower respiratory tract infection).<br>This study will be performed as follows:<br>• 2 doses 4 weeks apart of PTX-COVID19-B intramuscular (IM) vaccine or placebo<br>o Cohort 1: 20 healthy subjects 18 to 64 years of age administered 16 µg PTX-COVID19-B IM vaccine or placebo; 5 sentinel subjects |

(4 PTX-COVID19-B:1 placebo), followed by remaining 11 PTX-COVID19-B:4 placebo (cohort expansion)

  - Assessment of the sentinel cohort will include safety and tolerability follow-up assessment (including reactogenicity, solicited and unsolicited AEs, and safety laboratory evaluations) through the third day post vaccination, and an independent Safety Review Committee (iSRC) will decide on cohort expansion and dose escalation as applicable. If no Study Halting Rules are met, the next dose level will begin enrollment.
  - ~14 days between Cohort 1 (at least 7 days of AE and safety data) and Cohort 2 Day 1 for iSRC review

- Cohort 2: 20 healthy subjects 18 to 64 years of age administered 40 µg PTX-COVID19-B IM vaccine or placebo; 5 sentinel subjects (4 PTX-COVID19-B:1 placebo), followed by remaining 11 PTX-COVID19-B:4 placebo (cohort expansion)
  - Assessment of the sentinel cohort will include a safety and tolerability follow-up assessment (including reactogenicity, solicited and unsolicited AEs, and safety laboratory evaluations) through the third day post vaccination, and a iSRC will decide on cohort expansion and dose escalation as applicable. If no Study Halting Rules are met, the next dose level will begin enrollment.
  - ~14 days between Cohort 2 (at least 7 days of AE and safety data) and Cohort 3 Day 1 for iSRC review
- Cohort 3: 20 healthy subjects 18 to 64 years of age administered 100 µg PTX-COVID19-B IM vaccine or placebo; 5 sentinel subjects (4 PTX-COVID19-B:1 placebo), followed by remaining 11 PTX-COVID19-B:4 placebo (cohort expansion)
  - Assessment of the sentinel cohort will include a safety and tolerability follow-up assessment (including reactogenicity, solicited and unsolicited AEs, and safety laboratory evaluations) through the third day post vaccination, and an iSRC will decide on cohort expansion and dose escalation as applicable.

Subjects will visit the clinical site for screening visit (Days -21 to -1) and on Days 1, 8, 28 ± 2, 42 ± 2, 90 ± 3, 180 ± 5, and 395 - 14. Safety phone calls to subjects will be performed on Days 2 and 29. Screening procedures will include informed consent, evaluation of entry criteria, demographics, height, weight, body mass index, medical and surgical histories, safety evaluations (including SAE evaluations, recording prior medications and procedures, physical examinations, vital sign assessments, clinical blood and urine samples, nasopharyngeal [NP] swab with the option for the clinic to confirm eligibility via an additional point-of-care test), and blood samples for immunogenicity analysis (antibodies). On Day 1, randomization will be performed (overall 15:5 investigational vaccine:placebo. On Day 1 and Day 28, eligibility will be confirmed and safety evaluations, immunogenicity and cell mediated immunity blood sampling, paper diary training and distribution, and vaccine or placebo administration will be performed. Safety evaluations, blood and urine samples for safety evaluations, and blood samples for immunogenicity analysis and cell mediated immunity will be performed through the end of study visit. Assessments of unsolicited AEs will be performed on Day 1 through Day 42. Medically-attended AE assessments will be performed when subjects are in-house on Days 1 through 42. Assessments of NOCDs, AESIs (including COVID-19 cases for enhanced disease), and PIMMCs will be performed at visits from Day 1 through the end of study. The SAEs will be assessed throughout the study.

Vaccine and placebo will be prepared by an unblinded site pharmacist and administered by IM injection in the upper arm deltoid muscle of the non-dominant side at the clinical research site by unblinded CPU personnel. Subjects will be observed for immediate AEs and/or reactogenicity for approximately 1 hour after administration of vaccine. Subjects will be provided with a Diary Card and will be trained to record specifically-elicited systemic and local symptoms daily, as well as any additional AEs, during the follow-up period after each vaccination. Subjects will be requested to take a photo of completed Diary Card and text/email the photo to the site to ensure close oversight of reactions.

The duration of the study is approximately 14 months for each subject.

Interim Data Review:

The iSRC will convene between all cohorts to review available safety data (through the third day post-vaccination for the sentinel group and at least 7 days post vaccination of expanded cohort; for assessment AE and safety data between Cohorts) to ensure that there are no safety concerns and that none of the Study Halting Rules have been met before recommending that the study enroll the next cohort or study phase.

At approximately 2 weeks after the last subject receives their last dose, (Day 42) there will be an interim review and analysis of all data collected for safety and immunogenicity endpoints by the study team. The available data will be cleaned, and the database will be frozen and unblinded. The study team will analyze the available data for safety and immunogenicity endpoints, and then a clinical study report will be written for data from Day 1 through Day 42.. This unblinded review will be used for dose selection for the Phase 2/3 study.

Study Halting Rules:

The occurrence and confirmation of 1 or more of the following findings will result in suspension of further enrollment and vaccine administration pending urgent review (within 1 week) of the safety data by the iSRC.

1. One or more subjects experiences an SAE assessed as possibly related, probably related, or related to vaccine without a plausible alternative explanation.
2. One or more subjects have generalized urticaria within 3 days of vaccination for which there is no alternative plausible explanation.
3. One or more subjects develop a Grade 4 local reaction for which there is no alternative plausible explanation.
4. One or more subjects experience laryngospasm, bronchospasm, or anaphylaxis after vaccine administration considered related to the vaccine.
5. One or more subjects develop a fever > 40°C/104°F post vaccination that is assessed as possibly related, probably related, or related to vaccine without a plausible alternative explanation.
6. Two or more subjects within a cohort, or 2 or more subjects across cohorts, experience any Grade 3 or higher abnormality in the same laboratory parameter determined by the investigator or medical monitor as clinically significant and that are assessed as possibly related, probably related, or related to vaccine without a plausible alternative explanation.

   Note: An unblinded statistician or designee will assist with determining whether Study Halting Rule #6 is met.

| | |
|---|---|
| | 7. Two or more subjects across cohorts experience a Grade 3 or higher AE, AESI, or medically-attended AE of the same preferred term (as categorized by Medical Dictionary for Regulatory Activities [MedDRA]) assessed as possibly related, probably related, or related to vaccine without a plausible alternative explanation.<br>8. If no subject experiences a Grade 3 or higher AE, AESI, or medically attended AE by the third day post-vaccination, dosing may move forward; otherwise, dosing will be halted while the event is followed to resolution.<br>9. Death due to SARS Cov-2 infection in a study subject who was on the PTX-COVID19-B arm of the study. |
| **Study Rationale:** | There is currently a shortage of vaccines to prevent infections of SARS-CoV-2 or effective antiviral vaccines to treat COVID19. Given the crisis of COVID-19 infections in multiple countries and the fast expansion of the disease, rapid development of an effective vaccine is of the utmost importance. |
| **Subject Selection Criteria:** | <u>Inclusion Criteria:</u><br>Subjects must meet all inclusion criteria to be eligible for study participation. In addition, racial and ethnic minorities will be sought to obtain a diverse study population.<br>1. Subject has read, understood, and signed the informed consent form.<br>2. Healthy adult males and females 18 to 64 years of age, inclusive, at screening<br>3. Seronegative to SARS-CoV-2 and reverse transcription-polymerase chain reaction (RT-PCR)-negative at screening, without evidence of recent of exposure or viral respiratory disease not identified as influenza or RSV (febrile or lower respiratory tract infection)<br>4. Body mass index of $\geq 18$ and $\leq 30$ kg/m$^2$ at screening<br>5. Must be in general good health before study participation with no clinically relevant abnormalities that could interfere with study assessments.<br>6. Women of childbearing potential (WOCBP) and men whose sexual partners are WOCBP must be able and willing to use at least 1 highly effective method of contraception (i.e., including hysterectomy, bilateral salpingectomy, and bilateral oophorectomy, hormonal oral [in combination with male condoms with spermicide], transdermal, implant, or injection, barrier [i.e., condom, diaphragm with spermicide]; intrauterine device; vasectomized partner [6 months minimum], clinically sterile partner; or abstinence) during the study.<br>• A female subject is considered a WOCBP after menarche and until she is in a postmenopausal state for 12 consecutive months (without an alternative medical cause) or otherwise permanently sterile.<br>• Subjects not of childbearing potential are not required to use any other forms of contraception during the study. Non-childbearing potential is defined as subject confirmed:<br>- Surgical sterilization (e.g., bilateral oophorectomy, bilateral salpingectomy, bilateral occlusion by cautery [Essure System® is not acceptable], hysterectomy, or tubal ligation)<br>- Postmenopausal (defined as permanent cessation of menstruation for at least 12 consecutive months prior to screening); if postmenopausal status is unclear, pregnancy tests will be performed prior to vaccinations.<br>7. Women of childbearing potential must have a negative pregnancy test before each vaccination. If menopausal status is unclear, a pregnancy test is required. |

8. Must be able to attend all visits (scheduled and unscheduled, as applicable) for the duration of the study and comply with all study procedures, including daily completion of the Diary Card (Appendix A) after each injection.

Exclusion Criteria:

Subjects will not be eligible for study participation if they meet any of the exclusion criteria, or will be discontinued at the discretion of the investigator if they develop any of the exclusion criteria during the study.

1. History of an acute or chronic medical condition including dementia that, in the opinion of the investigator, would render vaccination unsafe or would interfere with the evaluation of responses.
2. History of any medical conditions that place subjects at higher risk for severe illness due to SARS-CoV-2 will be excluded including:
   - Chronic kidney disease
   - COPD (chronic obstructive pulmonary disease)
   - Heart conditions, such as heart failure, coronary artery disease, or cardiomyopathies
   - Any Immunocompromised state including from transplantation, history immunodeficiency, HIV, immunosuppressive drug intake.
   - Sickle cell disease
   - Current smoker or history of >5 pack/years of smoking.
   - Type 2 diabetes mellitus

   Subjects with history of any of the following conditions might be at an increased risk of complications from Covid 19 and will be excluded:

   - Asthma (moderate-to-severe)
   - Cerebrovascular disease (affects blood vessels and blood supply to the brain)
   - Cystic fibrosis
   - Hypertension or high blood pressure
   - Neurologic conditions, such as dementia
   - Liver disease
   - Pulmonary fibrosis (having damaged or scarred lung tissues)
   - Thalassemia (a type of blood disorder)
   - Type 1 diabetes mellitus
3. History of ongoing clinical condition or medication or treatments that may adversely affect the immune system.
4. Individuals who are seropositive or RT-PCR positive for SARS-CoV-2, including prior to a second dose of PTX-COVID19-B vaccine.
5. Individuals who are at increased risk of exposure to SARS-CoV-2 (e.g., healthcare workers, emergency responders).
6. Close contact of anyone known to have SARS-CoV-2 infection within 30 days prior to vaccine administration.
7. Living in a group setting or group care facility (e.g., dormitory, assisted living or nursing home).
8. Individuals with any elevated (Grade 1 or higher) laboratory test assessed as clinically significant for age/sex by the investigator at screening.
9. Individuals with any elevated for age/sex (Grade 1 or higher) liver function enzyme at screening, regardless of the appraisal of clinical significance (one

retest permitted). The criteria for excluding subjects with elevated liver enzymes are as follows:

- Alkaline phosphatase, alanine aminotransferase, aspartate aminotransferase, or gamma-glutamyl transferase > 1.5 × upper limit of normal (ULN)
- Total bilirubin > 1.5 × ULN

10. Active neoplastic disease (excluding nonmelanoma skin cancer that was successfully treated) or a history of any hematological malignancy. “Active” is defined as having received treatment within the past 5 years.
11. Long-term (> 2 weeks) use of oral or parenteral steroids or high-dose inhaled steroids (> 800 µg/day of beclomethasone dipropionate or equivalent) within 6 months before screening (nasal and topical steroids are allowed).
12. History of autoimmune, inflammatory disease, or PIMMCs (Appendix B).
13. Women currently pregnant, lactating, or planning a pregnancy between enrollment and 181 days after randomization.
14. History of Guillain-Barré Syndrome or any degenerative neurology disorder.
15. History of anaphylactic-type reaction to any injected vaccines.
16. Known or suspected hypersensitivity to 1 or more of the components of the vaccine.
17. History of alcohol abuse, illicit drug use, physical dependence to any opioid, or any history of drug abuse or addiction within 12 months of screening.
18. Acute illness or fever (temperature >37.5C) within 3 days before study enrollment (enrollment may be delayed for full recovery if acceptable to the investigator).
19. Individuals currently participating or planning to participate in a study that involves an experimental agent (vaccine, drug, biologic, device, or medication); or who have received an experimental agent within 1 month (3 months for immunoglobulins) before enrollment in this study; or who expect to receive another experimental agent during participation in this study.
20. Receipt of immunoglobulin or another blood product within the 3 months before enrollment in this study or those who expect to receive immunoglobulin or another blood product during this study.
21. Individuals who intend to donate blood within 6 months after the first vaccination and within 30 days post the last donation from Visit 1.
22. Individuals using prescription medications for prophylaxis of SARS-CoV-2.
23. Individuals who plan to receive another vaccine within the first 3 months of the study except influenza vaccine which should not be given within 2 weeks of vaccine.
24. Receipt of any other SARS-CoV-2 or other experimental coronavirus (Middle East Respiratory Syndrome, SARS etc.) vaccine at any time prior to or during the study.
25. Receipt of any investigational vaccine or investigational drug within 1 month of enrollment and through the end of the study (1 year after the last vaccination).
26. Plan to travel outside Canada from enrollment through Day 42.
27. History of surgery or major trauma within 12 weeks of screening, or surgery planned during the study.
28. Significant blood loss (> 400 mL) or has donated 1 or more units of blood or plasma within 6 weeks prior to study participation.

| | |
|---|---|
| | 29. Strenuous activity or significant alcohol intake (as assessed by the investigator) within 72 hours prior to safety laboratory sample collection.<br>30. Positive urine drugs of abuse screen.<br>31. Positive screen for human immunodeficiency virus-1 and -2 antibodies, hepatitis B surface antigen, or hepatitis C virus antibody.<br>32. Involved in the planning or conduct of this study.<br>33. Unwilling or unlikely to comply with the requirements of the study.<br>34. Subjects is an employee, contractor, or friend or relative of any employee of sponsor, CRO, study site, or site affiliate.<br>35. Subjects oximetry is <90% |
| **Vaccine, Dose, and Route of Administration:** | PTX-COVID19-B messenger ribonucleic acid Humoral Vaccine, injectable IM 0.2 mg/mL 0.5 mL solution, multiple dose levels as presented in Section 5.1 |
| **Reference Vaccine, Dose and Route of Administration:** | Control (Placebo) vaccine (saline), injectable IM solution 0.0 mg/mL 0.5 mL |
| **Planned Sample Size:** | No formal sample size calculation was performed.<br>A total of 60 adult subjects will be enrolled. |
| **Statistical Analysis:** | Analysis Populations:<br>Safety Population: All subjects who provide consent, are randomized, and receive any amount of vaccine/placebo. The Safety Population will be used for all safety analyses and analysis of immunogenicity and will be analyzed as actually treated.<br>Per Protocol Population: Includes all subjects in the Safety Population who receive the assigned doses of the vaccine/placebo according to protocol, have serology results, and have no major protocol deviations affecting the primary immunogenicity outcomes, as determined by the Sponsor before database lock and unblinding. The PP Population will be the primary population used for the analysis of safety endpoints.<br>Modified Intent-to-Treat Population: The mITT Population will be used for the analysis of immunogenicity endpoints. Analysis using the mITT Population will not be performed if it differs from the PP Population by ≤ 5% of the subjects for each of the treatment groups.<br><br>Immunogenicity Analysis:<br>Immunogenicity data for each cohort and study phase will be listed and summarized by phase, cohort, and time point using appropriate descriptive statistics.<br><br>Safety Analysis:<br>Vital signs, clinical laboratory tests, and physical examination findings will be listed and summarized by, cohort, treatment (as applicable) and time point using appropriate descriptive statistics.<br>The number and percentage of subjects reporting any treatment-emergent adverse event (TEAE) or reactogenicity will be summarized by study phase and cohort and tabulated by system organ class and preferred term (coded using MedDRA). The TEAEs will be further classified by severity and relationship and for SAEs, medically-attended AEs, NOCD, PIMMCs and AESIs.<br>Additionally, number of subjects who became infected after vaccination and whether being vaccinated made the disease less or more severe will be presented. |

#### LIST OF FIGURES

Figure 5.1 Study Design Schematic ....... 26

### 2 SCHEDULE OF ASSESSMENTS AND PROCEDURES

#### Table 2.1 Schedule of Assessments and Procedures

| | Screening | Treatment Period | | | | | | | | End of study [a] |
|---|---|---|---|---|---|---|---|---|---|---|
| **Study Procedure** **Day** | **-21 to -1** | **1 [b]** | **2** | **8** | **28** | **29** | **42** | **90** | **180** | **395** |
| **Window allowance (Days)** | | | | | **± 2** | | **± 2** | **± 3** | **± 5** | **-14** |
| Clinic visit | X | X | | X | X | | X | X | X | X |
| Telephone call (safety check) [c] | | | X | | | X | | | | |
| Informed consent | X | | | | | | | | | |
| Inclusion/exclusion criteria | X | X | | | | | | | | |
| Demographics | X | | | | | | | | | |
| Height, body weight, BMI | X [d] | | | | | | | | | |
| Medical and surgical history | X | | | | | | | | | |
| Complete physical examination | X | | | | | | | | | |
| Symptom-directed, targeted physical examination | | X | | X | X | | X | X | X | X |
| Vital signs [e] | X | X [f] | | X | X [f] | | X | X | X | X |
| Clinical laboratory tests [g] | X | | | X | X | | X | X | | |
| Serology (HIV-1 and -2, HBsAg, HCV) | X | | | | | | | | | |
| Pregnancy test (women of child-bearing potential) | X | X | | | X | | | | | |
| Urine drugs of abuse | X | | | | | | | | | |
| NP swab for COVID-19 via RT-PCR [h] | X | | | | X | | | | | |
| Blood samples for immunogenicity analysis (antibodies IgM, IgG, IgA, neutralization) | X | | | X | X | | X | X | X | X |
| Blood/PBMC samples for cell-mediated immunity (pre-dose as applicable) | | X | | X | X | | X | X | X | X |
| Randomization | | X [i] | | | | | | | | |

#### Table 2.1 Schedule of Assessments and Procedures

| | Screening | Treatment Period | | | | | | | | End of study [a] |
|---|---|---|---|---|---|---|---|---|---|---|
| **Study Procedure** **Day** | **-21 to -1** | **1 [b]** | **2** | **8** | **28** | **29** | **42** | **90** | **180** | **395** |
| Training of subjects on diary completion | | X | | | X | | | | | |
| Distribute paper diary | | X | | | X | | | | | |
| Review paper diary | | | | X | | | X | | | |
| Predose, confirm eligibility for vaccination | | X | | | X | | | | | |
| Vaccine administration [j] | | X | | | X | | | | | |
| Post-vaccination assessment (approximately 1 hour post vaccination, includes arm check and vital signs) | | X | | | X | | | | | |
| Medically-attended AE assessments [k] | | X | | X | X | | X | | | |
| Unsolicited AE assessments | X | | | | | | | | | |
| Solicited AEs [l] | | X | X | | X | X | | | | |
| Assessments of NOCD, AESIs (including COVID-19 cases for enhanced disease), and PIMMCs [h] | | X | | X | X | | X | X | X | X |
| Serious adverse event assessments | X | | | | | | | | | |
| Prior and concomitant medications/procedures/other vaccinations [m] | X | | | | | | | | | |

Abbreviations: AEs = adverse events; AESI = adverse event of special interest; BMI = body mass index; COVID-19 = coronavirus disease 2019; HBsAg = hepatitis B surface antigen; HCV = hepatitis C virus; HIV = human immunodeficiency virus; Ig = immunoglobulin; IM = intramuscular; NOCD = new onset chronic disease; NP = nasopharyngeal; PBMC = peripheral blood mononuclear cell; PIMMC = potential immune-mediated medical condition; RT-PCR = reverse transcription-polymerase chain reaction

a. Subjects who withdraw from the study early will have end-of-study procedures performed at the time of discontinuation.
b. Study entry assessments conducted on Day 1 will be used to reconfirm a subject's eligibility for enrollment into the study.
c. All subjects will have safety calls approximately 24 hours after each of the vaccinations. Trained site staff will inquire about the presence of solicited and unsolicited AEs and remind subjects to keep their diary cards up to date.
d. BMI will be calculated using the height obtained at screening.

e. Vital signs will be measured after the subject has been resting quietly in a seated position for at least 5 minutes. Vital signs include systolic and diastolic blood pressures, heart rate, respiratory rate, and oral temperature (between 35ºC and37.5ºC)-. Pulse oximetry will be performed prior to each vaccination (Pulse oximetry must be >90%). Unscheduled visits for COVID-like illness will require pulse oximetry in addition to standard vital signs. Temperature at all visits is to be between 35°C and 37.5°C

f. Vital signs on Days 1 and 28 will be collected before vaccination and approximately 1 hour after vaccination (before subjects are discharged from the clinic).

g. Hematology, chemistry, and urinalysis. Refer to Table 8.1 for a detailed list of clinical laboratory test parameters. As applicable, on vaccination days, laboratory samples will be collected prior to vaccination.

h. Serial NP swabs (RT-PCR) will be performed on subjects who report either potential exposure to SARS-CoV-2 (ie, a close contact tests positive for SARS-CoV-2) or symptoms of SARS-CoV-2 infection. This testing will occur every 2 weeks for a total of 4 weeks (if there are 2 consecutive negative tests) or until negative after a positive result. Positive results will be reported as an AE and subjects will be followed for disease severity, duration, and outcome. A negative NP swab test result is required prior to the second vaccination. In addition, the clinical site has the option to confirm eligibility via an additional point-of-care test.

i. Subjects will be randomized prior to vaccination.

j. Subjects will receive IM injection of investigational vaccine or placebo in the upper arm deltoid muscle of the non-dominant side according to the randomization. All subjects will remain at the study site for at least 60 minutes after vaccination for safety monitoring. An arm check and vital signs will be done at approximately 1 hour post injection.

k. Medically-attended AEs will be assessed from the time of the first study vaccination on Day 1 until Day 42 during clinic visits. Observations for asymptomatic disease are required prior to the second vaccination.

l. Solicited AEs through the third day post each vaccination.

m. Any medication taken by a subject from 30 days prior to the screening visit through Day 42 and the reason for its use will be documented. After Day 42, concomitant medications will be documented if associated with an SAE, AESI, medically-attended AE, NOCD, or PIMMC.

NOTES:

Unscheduled visits may be performed at any time for COVID-like illness and NP swab. If positive, subjects will be referred for standard of care and followed for severity, duration, and outcome.

In the event multiple procedures are required to be conducted at the same nominal time point, the timing of blood sample collections will take priority over all other scheduled activities. Blood samples must be performed prior to vaccinations.

### LIST OF ABBREVIATIONS

| Abbreviation | Definition |
|---|---|
| AAV | adeno-associated viruses |
| ACE-2 | angiotensin-converting enzyme 2 |
| AE | adverse event |
| AESI | AE of special interest |
| ALB | albino |
| BMI | body mass index |
| CDC | United States Centers for Disease Controls |
| CoV | coronavirus |
| COVID-19 | coronavirus disease 2019 |
| CPU | clinical pharmacology unit |
| CRA | clinical research associate |
| CRO | contract research organization |
| DILI | drug-induced liver injury |
| eCRF | electronic case report form |
| ELISA | enzyme-linked immunosorbent assay |
| EOS | end of study |
| GCP | Good Clinical Practice |
| GLP | Good Laboratory Practice |
| HBsAg | hepatitis B surface antigen |
| HCV | hepatitis C virus |
| HEK | human embryonic kidney |
| HIV | human immunodeficiency virus |
| ICD | International Classification of Disease |
| ICF | informed consent form |
| ICH | International Council for Harmonisation |
| ICU | intensive care unit |
| IEC | independent ethics committee |
| Ig | immunoglobulin |
| IM | intramuscular |
| IRB | institutional review board |
| iSRC | independent Safety Review Committee |
| LNP | lipid nanoparticle |
| MedDRA | Medical Dictionary for Regulatory Activities |
| MERS | Middle East Respiratory Syndrome |
| mITT | modified Intent-to-Treat |
| mRNA | messenger ribonucleic acid |
| N | number |
| NOCD | new onset chronic disease |
| NP | nasopharyngeal |
| PBMC | peripheral blood mononuclear cell |
| PIMMC | potential immune-mediated medical condition |

| | |
|---|---|
| PP | per protocol |
| RNA | ribonucleic acid |
| RSV | respiratory syncytial virus |
| RT-PCR | reverse transcription-polymerase chain reaction |
| SAE | serious adverse event |
| SAP | statistical analysis plan |
| SARS | Severe Acute Respiratory Syndrome |
| SARS-CoV-2 | Severe Acute Respiratory Syndrome coronavirus 2 |
| SOP | standard operating procedure |
| SRM | study reference manual |
| SSP | study-specific procedure |
| SUSAR | serious and unexpected suspected adverse reaction |
| TEAE | treatment-emergent adverse event |
| Th1 | T helper 1 |
| ULN | upper limit of normal |
| USA | United States of America |
| WHO | World Health Organization |
| WOCBP | women of childbearing potential |

#### STUDY ADMINISTRATIVE STRUCTURE

| | |
|---|---|
| Sponsor: | Providence Therapeutics Holdings Inc.<br>661 University Avenue, Suite 1300<br>Toronto, ON M5G 0B7 Canada |
| Sponsor Responsible Medical Officer and Contact: | Dr Piyush Patel Chief Medical Officer |
| Investigator and Clinical Pharmacology Unit: | Dr. Ben Lasko<br>Manna Research (Toronto)<br>2291 Kipling Avenue, Unit 117B<br>Toronto, Ontario, M9W 4L6, Canada<br><br>Telephone: (416) 740-2895 |
| Medical Monitor: | Piyush Patel, MD, FRCP<br>Telephone: (416) 419-8300 |
| Local Safety Laboratory for Hematology, Chemistry, Urinalysis, Serology, Drugs of Abuse Testing, HbA1c, and Pregnancy Testing: | LifeLabs Medical Laboratories<br>100 International Boulevard<br>Etobicoke, ON M9W 6J6 Canada<br>Telephone: (877) 849-3637 |
| Central Laboratory (Nasopharyngeal Swab for COVID-19 Testing via RT-PCR; Immunogenicity Analysis of anti-COVID-19 Titer Assays [IgG, IgM, and IgA by ELISA], HIV test): | Cirion Biopharma Research Inc.<br>3150 Delaunay, Laval (Quebec)<br>Canada, H7L 5E1 |
| Central Laboratory (Immunogenicity Analysis of anti-COVID-19 Neutralizing Antibody Titer Assay; Cell-mediated Immunity/PBMC Evaluations [Flow Cytometry, Enzyme Linked Immunospot Assay]): | Cirion Biopharma Research Inc.<br>3150 Delaunay, Laval (Quebec)<br>Canada, H7L 5E1<br>Telephone: 1-866-688-2474 |
| Clinical Research Organization: | ICON Clinical Research, LLC<br>820 West Diamond Avenue, Suite 100<br>Gaithersburg, MD 20878 USA<br>Telephone: 301-944-6800 |

### 3 INTRODUCTION AND BACKGROUND

#### 3.1 Introduction

The Vaccine Product, PTX-COVID19-B mRNA Humoral Vaccine, is intended for prevention of COVID-19 in a general population. PTX-COVID19-B mRNA Humoral Vaccine is composed of LNP containing an mRNA that encodes for the full-length S protein of SARS-CoV-2 to elicit a neutralizing immune response against the virus. The mRNA is the drug substance and the LNP containing the mRNA is the drug product. The lipid components of the LNP, cholesterol, distearoylphosphatidylcholine, PEG2000-C-DMA, and 3D-P-DMA encapsulate the mRNA and deliver it into the cells via cellular membrane and endosomal membrane fusion. The mRNA enters the cytosol, and it is transcribed and translated to express the S protein. The mRNA is an ~4.2kb sequence encoding for 1273 amino acid polypeptide, with a natural N-terminal signal peptide for entry into the endosomal pathway. The polypeptide will be eventually be anchored in the plasma membrane of cells that take up the LNP. PTX-COVID19-B mRNA Humoral Vaccine is preservative-free and will be supplied as a sterile parenteral in an aqueous cryoprotectant buffer for IM injection for vaccination. Additional information may be found in the CTA briefing document.

To date, 7 coronaviruses capable of human infection have been identified, including SARS-CoV, MERS CoV, and this newly identified CoV (SARS-CoV-2). These 3 viruses, when in the human population, are significant public health risks showing high fatality rates (John Hopkins 2020). Coronavirus (SARS-CoV-2) infection causes a newly spread viral disease (COVID-19), with no specific highly-effective therapy available. With a world-wide high infection rate, effective treatment is an unmet need. Originating in Wuhan (China), the SARS-CoV-2 virus spread rapidly to 235 countries and currently (14 Oct 2020) has been confirmed in over 38 million cases and caused over 1.08 million deaths according to the WHO (2020). On March 11, 2020, the WHO assessed COVID-19 as a pandemic. In Canada, 240,010 cases have been confirmed with 10,208 deaths and spread to all provinces and territories (WHO 2020). Cases and deaths in Canada have continued to climb. To help patients affected by the COVID-19 pandemic, Providence Therapeutics is proposing to evaluate a COVID-19 mRNA vaccine (PTX-COVID19-B) that is designed to induce immunity against COVID-19 through neutralizing antibodies against the S protein of SARS-CoV-2, likely interacting with the receptor binding domain (RBD).

#### 3.2 Background and Summary of Findings to Date - Nonclinical Experience

In a rapid response to the COVID-19 global crisis, Providence re-focused the vast majority of its efforts, which had been focused on personalized cancer vaccines, to making a prophylactic vaccine for SARS-CoV-2, the causative agent of COVID-19. Providence first organized several meetings with experts in the fields of immunology, virology, structural biology, and vaccinology to derive a plan for moving forward rapidly. Providence started executing that plan in April 2020. The first step in making an mRNA vaccine was antigen selection. In collaboration with experts, Providence designed the sequences for multiple different antigens based on the S protein of SARS-CoV-2. The production of the mRNA encoding the antigens is complete and the multiple forms of vaccine candidates were tested.

Providence worked with the laboratory of a leading Canadian virologist, Mario Ostrowski, at the University of Toronto. Dr. Ostrowski has access to patients in the greater Toronto area who have recovered from COVID-19. Dr. Ostrowski had already collected blood and isolated immune cells from 11 patients and continues to collect more. His laboratory isolated and cultured the SARS-CoV-2 virus and has a Biosafety Level 3 facility for performing experiments with viral infection. Providence used these materials and facilities to characterize the human immune response to SARS-CoV-2 and to the vaccine candidates. The initial vaccine designs were produced at research scale and delivered to Dr. Ostrowski's laboratory for testing in human cells to prove they induce the production of the encoded proteins. It was demonstrated that the Providence mRNA can instruct human cells to make the SARS-CoV-2 S protein, or the RBD fragment of the S protein, and that these proteins are recognized by antibodies in the sera of a recovered COVID-19 patient or by a commercially available SARS-CoV-2 S protein monoclonal antibody.

Larger batches of these 3 vaccine candidates were made, formulated, and tested in mice in June 2020 to prove they could produce a neutralizing antibody response. These experiments involved injecting mice intramuscularly with 20 µg of the vaccine candidates twice at 3-week intervals and determining which candidates were most effective at causing the mice to produce antibodies that could neutralize SARS-CoV-2 from being able to infect human cells in culture. The lead candidate from the first experiment, PTX-COVID19-B (LNP formulated full-length SARS-CoV-2 S protein), was then tested in the same dosing regimen with doses of either 1 or 10 µg. Induction of T cell responses against the S protein were seen almost equally at both dose levels, while neutralizing antibody levels were much higher in the animals from the 10 µg dose cohort. Further characterization of the antibody isotypes and cytokine production indicated a Th1-induced response upon vaccination with PTX-COV19-B. Additionally, 2 different models were used in order to attempt to see whether PTX-COVID19-B could prevent animals from being infected with SARS-CoV-2. The first was a mouse model, in which the mice were first immunized with PTX-COVID19-B, then infected with an AAV strain that induced the expression of human ACE-2 (the SARS-Cov_2 receptor) in the airways of the mice before the mice were infected with a clinical isolate of SARS-CoV-2 by intranasal administration. All mice (N = 12/group) immunized with 20 or 4 µg of PTX-COVID19-B were completely protected from infection, as determined by having undetectable amounts of SARS-CoV-2 RNA in their lungs 4 days post infection, while 11 of the 12 mock immunized mice had easily detectable SARS-CoV-2 RNA. Of the 12 mice immunized with 1 µg of PTX-COVID19-B, 2 had detectable SARS-CoV-2 RNA while 9 did not, thus there was a dose response effect.

In discussions with the leading non-clinical CRO in Canada, Charles River Laboratories, Providence designed an experiment to test immunogenicity and local tolerability in mice. The data generated further supported the Providence clinical trial application, as there were no significant safety issues.

Significant non-clinical studies are summarized in Table 3.1.

**Table 3.1 Summary of Significant Non-clinical Studies in Support of COVID-19 mRNA Humoral Vaccine**

| **Study Description (Dose Levels)** | **Test System** | **Summary of Results** |
|---|---|---|
| Production of SARS-CoV-2 S protein in response to vaccine | HEK293 cells and recovered patient sera | Patient sera did react to cells treated with the vaccine, but not untreated cells |
| Production of neutralizing antibodies (20 µg) | C57BL/6 mice | Vaccines encoding three different forms of the S protein tested, both full-length S protein vaccines produced high titers, wild-type S chosen as candidate |
| Production of neutralizing antibodies (1 and 10 µg) | C57BL/6 mice | Ability of candidate vaccine to produce high titers of neutralizing antibodies confirmed, activation of T cells and a TH1 biased response also demonstrated |
| Immunogenicity and local tolerability (4 and 20 µg) | BALB/C mice | Tests performed prior to Phase 1 clinical study:<br>6-Week (q3wX2) IM Injection Vaccine and Toxicity Study Of mRNA/LNP in the BALB/C Mouse (non-GLP, study currently ongoing) |
| Mouse AAV6-hACE2 challenge model (20 µg) | C57BL/6 mice | Mice given two injections of vaccine 3 weeks apart, 15 days after second dose mice are infected intranasally with virus encoding hACE2 (receptor for SARS-CoV-2) and 6 days later challenged with SARS-Cov-2 to determine if vaccine protected from infection (non-GLP, study currently ongoing) |
| Hamster challenge model (20 µg) | Syrian golden hamsters | Hamsters given two injections of vaccine 3 weeks apart, 21 days after second dose hamsters are challenged with SARS-Cov-2 to determine if vaccine protected from infection (non-GLP, study currently ongoing) |

Abbreviations: ALB = albino; GLP = Good Laboratory Practice; HEK = human embryonic kidney; IM = intramuscular; LNP = lipid nanoparticle; mRNA = messenger ribonucleic acid; SARS-CoV-2 = Severe Acute Respiratory Syndrome coronavirus 2; Th1 = T helper 1

#### 3.3 Study Rationale

##### 3.3.1 Dose Rationale

Given the large number of doses required for the Canadian population, Providence proposes to develop a multi-dose vaccine presentation, where 1 vial would contain sufficient vaccine product to fill multiple syringes for administration to multiple recipients. This approach would help to address issues of limited fill/finish capacity, storage of vials, and distribution of the vaccine.

There is no preservative in the vaccine product formulation; therefore, once a vial is opened, it must be used within 6 hours of opening.

The initial trial will be a first-in-human Phase I trial with 3 dose levels of 16, 40, and 100 µg. Two injections of the vaccine will be administered at intervals of 4 weeks.

Based on the nonclinical studies conducted thus far, the results demonstrated that PTX-COVID19-B vaccine can be effective at doses as low as 1 µg and nearly maximally effective by a dose of 4 µg. In addition, doses as high as 20 µg PTX-COVID19-B vaccine did not produce any significant safety signals in mice (CRL5003561).

Furthermore, PTX-COVID19-B works through the same mechanism and chemically is very similar to the vaccines of Pfizer/BioNTech (ClinicalTrials.gov Identifier: NCT04368728) and Moderna (ClinicalTrials.gov Identifier: NCT04470427). These companies tested doses over a similar range in their Phase I studies and selected dose levels of 30 µg and 100 µg respectively for their Phase III trials. Based on nonclinical studies and doses tested on similar vaccine products, Providence selected 16 µg, 40 µg and 100 µg as appropriate dose levels for the Phase I study as per study protocol, PRO-CL-001.

##### 3.3.2 Study Design Rationale

This first-in-human clinical study will be performed in healthy subjects who are SARS-CoV-2 seronegative and RT-PCR negative and are without evidence of recent exposure or viral respiratory disease that has not been identified as influenza or respiratory syncytial virus (febrile or lower respiratory tract infection). The investigational vaccine will be administered in approximately 60 healthy subjects 18-64 years of age. Three cohorts of 20 subjects each will include sentinel groups of 5 subjects randomized 4:1 investigational vaccine:placebo, followed by a cohort expansion phase of 15 subjects randomized 11:4 to investigational vaccine:placebo. Assessment of the sentinel cohort will include safety and tolerability follow-up assessment (including reactogenicity, solicited and unsolicited AEs, and safety laboratory evaluations) through the third day post each vaccination, and the iSRC will decide on cohort expansion and dose escalation.

Subjects will be recruited from 1 clinical research site with vaccine experience and access to all age groups in Canada.

At 14 days post-second dose, there will be a review of all available data collected for safety and immunogenicity endpoints to determine the appropriate dose, plan, and study design for moving forward.

The duration of the study is approximately 14 months for each subject.

### 4 OBJECTIVES

#### 4.1 Objectives

The study objectives are as follows:

- To evaluate the safety and tolerability of 2 doses of PTX-COVID19-B vaccine in healthy seronegative adults 18-64 years of age
- To evaluate the immunogenicity of 2 doses of PTX-COVID19-B vaccine in healthy seronegative adults 18-64 years of age

#### 4.2 Endpoints

Safety and tolerability:

- Occurrence of events during the follow-up after each vaccination will be analyzed using both the PP and Safety Populations:
    - Vital signs and administration site reactions (e.g., arm check evaluations including pain, tenderness, erythema/redness, induration/swelling) during the follow-up after each vaccination
    - Daily solicited adverse events (e.g., fever, chills, nausea/vomiting, diarrhea, headache, fatigue, myalgia) through the second day post each vaccination.
- Overall safety will be analyzed using both the Safety and Per Protocol Populations.
    - Unsolicited adverse events from Day 1 through Day 42
    - Medically attended AEs (Day 1 through Day 42), NOCD, SAEs, AESIs, and PIMMCs from Day 1 through to Day 395 (approximately 1 year after the last vaccination)
    - Findings from targeted physical examinations, vital sign assessments, and clinical safety laboratory testing

Immune Response:

- Immunogenicity analysis using the Safety, Per Protocol, and mITT Populations. Analysis using the mITT Population will not be performed if it differs from the PP Population by ≤ 5% of the subjects for each of the treatment groups:
    - Antibodies (anti-COVID-19 immunoglobulin (Ig), IgG, IgA [ELISA]; and anti-COVID-19 neutralization titer assays)
    - Cell-mediated immunity using blood/PBMCs (Flow Cytometry, Enzyme Linked Immunospot Assay)

### 5 STUDY DESIGN

#### 5.1 Study Design and Overview

This is a Phase I, first-in-human, observer-blinded, randomized, placebo-controlled, ascending dose study to evaluate the safety, tolerability, and immunogenicity of PTX-COVID19-B vaccine in healthy seronegative adults aged 18-64. The study is designed with and dose-escalations and will be performed in seronegative adult subjects without evidence of recent of exposure to SARS-CoV-2 or viral respiratory disease not identified as influenza or RSV (febrile or lower respiratory tract infection).

This study will be performed as follows:

- 2 doses 4 weeks apart of PTX-COVID19-B IM vaccine or placebo
    - Cohort 1: 20 healthy subjects 18 to 64 years of age administered 16 µg PTX-COVID19-B IM vaccine or placebo; 5 sentinel subjects (4 PTX-COVID19-B:1 placebo), followed by remaining 11 PTX-COVID19-B:4 placebo (cohort expansion)
        - Assessment of the sentinel cohort will include a safety and tolerability follow-up assessment (including reactogenicity, solicited and unsolicited AEs, and safety laboratory evaluations) through the third day post vaccination, and an iSRC will decide on cohort expansion and dose escalation as applicable. If no Study Halting Rules are met (Section 6.7.2), the next dose level will begin enrollment.
        - ~14 days between Cohort 1 (at least 7 days of AE and safety data) and Cohort 2 Day 1 for iSRC review
    - Cohort 2: 20 healthy subjects 18 to 64 years of age administered 40 µg PTX-COVID19-B IM vaccine or placebo; 5 sentinel subjects (4 PTX-COVID19-B:1 placebo), followed by remaining 11 PTX-COVID19-B:4 placebo (cohort expansion)
        - Assessment of the sentinel cohort will include a safety and tolerability follow-up assessment (including reactogenicity, solicited and unsolicited AEs, and safety laboratory evaluations) through the third day post vaccination, and an iSRC will decide on cohort expansion and dose escalation as applicable. If no Study Halting Rules are met (Section 6.7.2), the next dose level will begin enrollment.
        - ~14 days between Cohort 2 (at least 7 days of AE and safety data) and Cohort 3 Day 1 for iSRC review
    - Cohort 3: 20 healthy subjects 18 to 64 years of age administered 100 µg PTX-COVID19-B IM vaccine or placebo; 5 sentinel subjects

(4 PTX-COVID19-B:1 placebo), followed by remaining 11 PTX-COVID19-B:4 placebo (cohort expansion)

- Assessment of the sentinel cohort will include a safety and tolerability follow-up assessment (including reactogenicity, solicited and unsolicited AEs, and safety laboratory evaluations) through the third day post vaccination, and an iSRC will decide on cohort expansion and dose escalation as applicable.

Subjects will visit the clinical site for screening visit (Days -21 to -1) and on Days 1, 8, 28 ± 2, 42 ± 2, 90 ± 3, 180 ± 5, and 395 - 14. Safety phone calls to subjects will be performed on Days 2 and 29. Screening procedures will include informed consent, evaluation of entry criteria, demographics, height, weight, BMI, medical and surgical histories, safety evaluations (including SAE evaluations, recording prior medications and procedures, physical examinations, vital sign assessments, clinical blood and urine samples, NP swab with the option for the clinic to confirm eligibility via an additional point-of-care test), and blood samples for immunogenicity analysis (antibodies). On Day 1, randomization will be performed (overall 15:5 investigational vaccine:placebo). On Day 1 and Day 28, eligibility will be confirmed and safety evaluations, immunogenicity and cell mediated immunity blood sampling, paper diary training and distribution, and vaccine or placebo administration will be performed. Safety evaluations, blood and urine samples for safety evaluations, and blood samples for immunogenicity analysis and cell mediated immunity will be performed through the end of study visit. Assessments of unsolicited AEs will be performed on Day 1 through Day 42. Medically-attended AE assessments will be performed when subjects are in-house on Days 1 through 42. Assessments of NOCD, AESIs (including COVID-19 cases for enhanced disease), and PIMMCs will be performed at visits from Day 1 through the end of study. Solicited AEs will be assessed through the seventh day post each vaccination. The SAEs will be assessed throughout the study (refer to Table 2.1).

Vaccine and placebo will be prepared by an unblinded site pharmacist and administered by IM injection in the upper arm deltoid muscle of the non-dominant side at the clinical research site by unblinded CPU personnel. Subjects will be observed for immediate AEs and/or reactogenicity for approximately 1 hour after administration of vaccine. Subjects will be provided with a Diary Card (Appendix A) and will be trained to record specifically-elicited systemic and local symptoms daily, as well as any additional AEs, during the follow-up period after each vaccination. Subjects will be requested to take a photo of completed Diary Card and text/email the photo to the site to ensure close oversight of reactions.

A schematic of the study design is presented in Figure 5.1.

**Figure 5.1 Study Design Schematic**

| **Phase 1:** | | | | |
|---|---|---|---|---|
| Subjects 18 to 64 years of age | | | | |
| Cohort 1<br>5 Sentinel Subjects, Safety Review ➡ Remaining 15 Subjects | ~14 Days, ➡ iSRC Meets | Cohort 2<br>5 Sentinel Subjects, Safety Review ➡ Remaining 15 Subjects | ~14 Days, ➡ iSRC Meets | Cohort 3<br>5 Sentinel Subjects, Safety Review ➡ Remaining 15 Subjects |

Abbreviation: iSRC = independent Safety Review Committee

##### 5.1.1 Interim Data Review

The iSRC will convene between all cohorts to review available safety data (through the third day post-vaccination for the sentinel group and at least 7 days post vaccination of expanded cohort; for assessment AE and safety data between Cohorts, refer to Section 5) to ensure that there are no safety concerns and that none of the Study Halting Rules (Section 6.7.2) have been met before recommending that the study enroll the next cohort or study phase.

At approximately 2 weeks after the last subject receives their last dose, (Day 42) there will be an interim review and analysis of all data collected for safety and immunogenicity endpoints by the study team. The available data will be cleaned, and the database will be frozen and unblinded. The study team will analyze the available data for safety and immunogenicity endpoints, and then a clinical study report will be written for data from Day 1 through Day 42.

When longer term follow-up (Day 57 to Day 395) has been completed for all subjects after the end of the study, further reviews and analysis will be performed on all data. An addendum to the clinical study report will also be written.

##### 5.1.2 Duration of Study

Each subject will receive 2 doses of vaccine or placebo, the first on Day 1 and the second on Day 28. The total duration of subject participation is approximately 14 months (including screening, treatment, and follow-up periods).

##### 5.1.3 Definition of Study Completion

End-of-study procedures will be performed as specified in the schedule of assessments (Table 2.1); subjects who withdraw from the study early will have EOS procedures performed at the time of discontinuation. If a subject refuses to return for the EOS blood samples, the safety follow-up may be completed via telephone. Subjects with ongoing clinically significant clinical or laboratory findings will be followed until the finding is resolved or medically stable; reasonable attempts will be made to follow-up with subjects. The subject’s participation in the study will end once all study assessments and follow-up- have been completed.

##### 5.1.4 End of Study

The end of the study is defined as the date when the last subject has completed all study procedures up to and including the EOS/early termination visit as specified in the schedule of assessments (Table 2.1).

### 6 SELECTION AND WITHDRAWAL OF SUBJECTS

#### 6.1 Inclusion Criteria

Subjects must meet all inclusion criteria to be eligible for study participation. In addition, racial and ethnic minorities will be sought to obtain a diverse study population.

1. Subject has read, understood, and signed the informed consent form.
2. Healthy adult males and females 18 to 64 years of age, inclusive, at screening
3. Seronegative to SARS-CoV-2 and RT-PCR negative at screening, without evidence of recent of exposure or viral respiratory disease not identified as influenza or RSV (febrile or lower respiratory tract infection)
4. BMI of $\geq 18$ and $\leq 30$ kg/m$^2$ at screening
5. Must be in general good health before study participation with no clinically relevant abnormalities that could interfere with study assessments.
6. Women of childbearing potential and men whose sexual partners are WOCBP must be able and willing to use at least 1 highly effective method of contraception (i.e., including hysterectomy, bilateral salpingectomy, and bilateral oophorectomy, hormonal oral [in combination with male condoms with spermicide], transdermal, implant, or injection, barrier [i.e., condom, diaphragm with spermicide]; intrauterine device; vasectomized partner [6 months minimum], clinically sterile partner; or abstinence) during the study.
    - A female subject is considered a WOCBP after menarche and until she is in a postmenopausal state for 12 consecutive months (without an alternative medical cause) or otherwise permanently sterile.
    - Subjects not of childbearing potential are not required to use any other forms of contraception during the study. Non-childbearing potential is defined as subject-confirmed:
        - Surgical sterilization (e.g., bilateral oophorectomy, bilateral salpingectomy, bilateral occlusion by cautery [Essure System® is not acceptable], hysterectomy, or tubal ligation)
        - Postmenopausal (defined as permanent cessation of menstruation for at least 12 consecutive months prior to screening); if postmenopausal status is unclear, pregnancy tests will be performed prior to vaccinations.
7. Women of childbearing potential must have a negative pregnancy test before each vaccination. If menopausal status is unclear, a pregnancy test is required.
8. Must be able to attend all visits (scheduled and unscheduled, as applicable) for the duration of the study and comply with all study procedures, including daily completion of the Diary Card (Appendix A) after each injection.

#### 6.2 Exclusion Criteria

Subjects will not be eligible for study participation if they meet any of the exclusion criteria or will be discontinued at the discretion of the investigator if they develop any of the exclusion criteria during the study.

1. History of an acute or chronic medical condition including dementia that, in the opinion of the investigator, would render vaccination unsafe or would interfere with the evaluation of responses.
2. History of any medical conditions that place subjects at higher risk for severe illness due to SARS-CoV-2 will be excluded including:
    - Chronic kidney disease
    - COPD (chronic obstructive pulmonary disease)
    - Heart conditions, such as heart failure, coronary artery disease, or cardiomyopathies
    - Any Immunocompromised state including from transplantation. History of immunodeficiency, HIV, or immunosuppressive drug intake.
    - Pregnancy
    - Sickle cell disease
    - Current smoker or history of >5pack/years of smoking.
    - Type 2 diabetes mellitus

    Subjects with a history of any of the following conditions might be at an increased risk of complications from Covid 19 and will be excluded:
    - Asthma (moderate-to-severe)
    - Cerebrovascular disease (affects blood vessels and blood supply to the brain)
    - Cystic fibrosis
    - Hypertension or high blood pressure
    - Neurologic conditions, such as dementia
    - Liver disease
    - Pulmonary fibrosis (having damaged or scarred lung tissues)
    - Thalassemia (a type of blood disorder)
    - Type 1 diabetes mellitus
3. History of ongoing clinical condition or medication or treatments that may adversely affect the immune system.
4. Individuals who are seropositive or RT-PCR positive for SARS-CoV-2, including prior to a second dose of PTX-COVID19-B vaccine.
5. Individuals who are at increased risk of exposure to SARS-CoV-2 (e.g., healthcare workers, emergency responders).
6. Close contact of anyone known to have SARS-CoV-2 infection within 30 days prior to vaccine administration.

7. Living in a group setting or group care facility (e.g., dormitory, assisted living or nursing home).
8. Individuals with any elevated (Grade 1 or higher) laboratory test assessed as clinically significant for age/sex by the investigator at screening. (Guidance for Industry: Toxicity Grading Scale for Healthy Adult and Adolescent Volunteers Enrolled in Preventive Vaccine Clinical Trials (September 2007), Tables for Laboratory Abnormalities).
9. Individuals with any elevated for age/sex (Grade 1 or higher) liver function enzyme at screening, regardless of the appraisal of clinical significance (one retest permitted). The criteria for excluding subjects with elevated liver enzymes are as follows:
    - Alkaline phosphatase, alanine aminotransferase, aspartate aminotransferase, or gamma-glutamyl transferase > 1.5 × ULN
    - Total bilirubin > 1.5 × ULN
10. Active neoplastic disease (excluding nonmelanoma skin cancer that was successfully treated) or a history of any hematological malignancy. "Active" is defined as having received treatment within the past 5 years.
11. Long-term (> 2 weeks) use of oral or parenteral steroids or high-dose inhaled steroids (> 800 µg/day of beclomethasone dipropionate or equivalent) within 6 months before screening (nasal and topical steroids are allowed).
12. History of autoimmune, inflammatory disease, or PIMMCs (Appendix B).
13. Women currently pregnant, lactating, or planning a pregnancy between enrollment and 181 days after randomization.
14. History of Guillain-Barré Syndrome or any degenerative neurology disorder.
15. History of anaphylactic-type reaction to any injected vaccines.
16. Known or suspected hypersensitivity to 1 or more of the components of the vaccine.
17. History of alcohol abuse, illicit drug use, physical dependence to any opioid, or any history of drug abuse or addiction within 12 months of screening.
18. Acute illness or fever (temperature >37.5C) within 3 days before study enrollment (enrollment may be delayed for full recovery if acceptable to the investigator).
19. Individuals currently participating or planning to participate in a study that involves an experimental agent (vaccine, drug, biologic, device, or medication); or who have received an experimental agent within 1 month (3 months for immunoglobulins) before enrollment in this study; or who expect to receive another experimental agent during participation in this study.

20. Receipt of immunoglobulin or another blood product within the 3 months before enrollment in this study or those who expect to receive immunoglobulin or another blood product during this study.
21. Individuals who intend to donate blood within 6 months after the first vaccination, and within 30 days past the last donation from Visit 1.
22. Individuals using prescription medications for prophylaxis of SARS-CoV-2.
23. Individuals who plan to receive another vaccine within the first 3 months of the study except influenza vaccine which should not be given within 2 weeks of vaccine.
24. Receipt of any other SARS-CoV-2 or other experimental coronavirus (MERS, SARS etc.) vaccine at any time prior to or during the study.
25. Receipt of any investigational vaccine or investigational drug within 1 month of enrollment and through the end of the study (1 year after the last vaccination).
26. Plan to travel outside Canada from enrollment through Day 42.
27. History of surgery or major trauma within 12 weeks of screening, or surgery planned during the study.
28. Significant blood loss (> 400 mL) or has donated 1 or more units of blood or plasma within 6 weeks prior to study participation.
29. Strenuous activity or significant alcohol intake (as assessed by the investigator) within 72 hours prior to safety laboratory sample collection.
30. Positive urine drugs of abuse screen.
31. Positive screen for HIV-1 and HIV-2 antibodies, HBsAg, or HCV antibody.
32. Involved in the planning or conduct of this study.
33. Unwilling or unlikely to comply with the requirements of the study.
34. Subjects is an employee, contractor, or friend or relative of any employee of sponsor, CRO, study site, or site affiliate.
35. Subjects' oximetry is <90%

##### 6.3 Screen Failures

Subjects who are discontinued from the study prior to vaccine administration will be considered screen failures and may be re-screened as long as the subject was not discontinued from the study due to noncompliance with the protocol (e.g., positive urine drugs of abuse screen, etc.). If the subject is re-screened, the subject must be re-consented.

Screen failure data will not be recorded in the eCRF.

##### 6.4 Decision to Administer Vaccination

Prior to administering the vaccination, subjects will have the following procedures/assessments performed or reviewed to confirm continued eligibility:

- Inclusion/exclusion criteria recheck (including absence of acute illness or new medical condition)
- Targeted physical examination
- Vital signs
    - Temperature at all visits must be between 35ºC and 37.5ºC
    - Pulse oximetry must be >90%
- Review of clinical safety laboratory tests to date
- Pregnancy test (as applicable)
- SAE and AE assessment and assessment of medically-attended AEs, NOCD, AESIs, and PIMMCs
- Observations for asymptomatic disease
- Negative NP test
- Check of halting rules (Section 6.7.2)

All assessments must be in agreement with inclusion/exclusion criteria and not clinically significant in order for a subject to receive the second vaccination dose. Subjects presenting for the first vaccination who have any of the above, will be rescheduled to another cohort if appropriate. Subjects presenting for the second vaccination who have any of the above criteria may be rescheduled up to one week later at the discretion of the Principal Investigator.

##### 6.5 Individual Discontinuation Criteria

The occurrence of the following findings in an individual subject may result in early termination from the study.

- Subjects experiences an SAE assessed as possibly related, probably related, or related to vaccine without a plausible alternative explanation.
- Subject has generalized urticaria within 7 days post vaccination without a plausible alternate explanation.
- Subject develops a Grade 4 local reaction for which there is no alternative plausible explanation.
- Subject experiences vaccine-associated enhanced respiratory disease (e.g., laryngospasm, bronchospasm, or anaphylaxis after vaccine administration, considered related to the vaccine).

- Subject develops a fever > 40°C/104°F post vaccination that is assessed as possibly related, probably related, or related to vaccine without a plausible alternative explanation.
- Subject experiences any Grade 3 or higher abnormality in the same laboratory parameter determined by the investigator or medical monitor as clinically significant and that are assessed as possibly related, probably related, or related to vaccine without a plausible alternative explanation.
- Subjects testing positive or develop symptoms related to infection with SARS-CoV2 after the first dose of the vaccine but before the second dose, will not be given any further doses of the study medication but will be followed to the end of the study.

##### 6.6 Subject Withdrawal

Subjects are free to withdraw from the study at any time, for any reason, and without prejudice to further treatment. The investigator may withdraw a subject if, in the investigator's judgment, continued participation would pose unacceptable risk to the subject or to the integrity of the study data. All procedures for early withdrawal must be completed. Reasons for withdrawal may include:

- AE
- SAE that is possibly related, probably related, or related, without a plausible alternative explanation
- Lost to follow-up
- Physician decision
- Pregnancy
- Protocol deviation
- Study terminated by sponsor
- Withdrawal by subject
- Death
- Other

Subjects who are withdrawn for reasons other than AEs may be replaced.

In the event of a subject's withdrawal, the investigator will promptly notify the Sponsor and will make every effort to complete the EOS assessments. All withdrawn subjects with ongoing clinically significant clinical or laboratory findings will be followed until the finding is resolved or medically stable; reasonable attempts will be made to follow-up with subjects.

Subjects may be withdrawn from the study if they meet specific Study Halting Rules (Section 6.7.2).

#### 6.7 Independent Safety Review Committee, Study Halting Rules, and Study Termination

##### 6.7.1 Independent Safety Review Committee

Safety reviews will be performed by the iSRC according to the details established at the start of the study and provided in the iSRC charter. The iSRC will consist of, at a minimum, 2 physicians with experience in vaccines and 1 statistician.

Within cohorts with sentinel subjects, the iSRC will review available safety and tolerability (reactogenicity, solicited and unsolicited AEs, and safety laboratory evaluations) data through approximately the third day post each vaccination prior to the enrollment of the remaining cohort subjects.

Cohort dose continuation/escalation will only occur if the iSRC determines that the current dose was safe and well tolerated and no study halting rules were met. The decision to allow dose continuation/escalation into the next cohort and/or age group will be documented and this decision, along with the safety information, will be communicated to the site.

The iSRC will also meet on an ad hoc basis if any of the Study Halting Rules are met (Section 6.7.2) based on new or emerging data. If a Study Halting Rule is met, study vaccinations will pause and the iSRC will proceed with an unblinded review of available safety data. After the meeting, the iSRC will make 1 or more of the following recommendations:

- All vaccinations should resume
- Specific cohorts should be discontinued while other cohorts should resume vaccinations
- No vaccinations should resume
- Modifications to study conduct (e.g., additional safety or laboratory assessments)

The blinded medical monitor may also assess cumulative safety information for this study per the Medical Monitoring Plan and advise the Sponsor of any of the safety findings (Section 6.7.2) have occurred.

The iSRC will also review unblinded safety data for any subject who dies or requires ICU admission due to SARS-CoV-2 infection. If the subject had received active vaccine, the iSRC may decide whether a study halt for vaccine-enhanced disease is required based on a review of all available clinical and preclinical safety and immunogenicity data.

##### 6.7.2 Study Halting Rules

The occurrence and confirmation of 1 or more of the following findings will result in suspension of further enrollment and vaccine administration pending urgent review (within ~1 week) of the safety data by the iSRC.

1. One or more subjects experiences an SAE assessed as possibly related, probably related, or related to vaccine without a plausible alternative explanation.
2. One or more subjects have generalized urticaria within 3 days of vaccination for which there is no alternative plausible explanation.
3. One or more subjects develop a Grade 4 local reaction for which there is no alternative plausible explanation.
4. One or more subjects experience laryngospasm, bronchospasm, or anaphylaxis after vaccine administration considered related to the vaccine.
5. One or more subjects develop a fever > 40°C/104°F post vaccination that is assessed as possibly related, probably related, or related to vaccine without a plausible alternative explanation.
6. Two or more subjects within a cohort, or 2 or more subjects across cohorts, experience any Grade 3 or higher abnormality in the same laboratory parameter determined by the investigator or medical monitor as clinically significant and that are assessed as possibly related, probably related, or related to vaccine without a plausible alternative explanation.

   Note: An unblinded statistician or designee will assist with determining whether Study Halting Rule #6 is met.
7. Two or more subjects across cohorts experience a Grade 3 or higher AE, AESI, or medically -attended AE of the same preferred term (as categorized by MedDRA) assessed as possibly related, probably related, or related to vaccine without a plausible alternative explanation.
8. Death due to SARS Cov-2 infection in a study subject who was on the PTX-COVID19-B arm of the study.
9. If no subject experiences a Grade 3 or higher AE, AESI, or medically -attended AE by the third day post-vaccination, dosing may move forward; otherwise, dosing will be halted while the event is followed to resolution.

###### 6.7.3 Study Termination

The study may be terminated at any time by the sponsor if serious side effects occur, if potential risks to study participants are identified, if the investigator does not adhere to the protocol, or if, in the sponsor’s judgment, there are no further benefits to be achieved from the study. If clinical development of the vaccine is discontinued, the sponsor will inform all investigators/institutions and regulatory authorities.

### 7 TREATMENT OF SUBJECTS

#### 7.1 Identity of Vaccine

The vaccine product, PTX-COVID19-B mRNA Humoral Vaccine, is a sterile formulation of mRNA (drug substance) in aqueous buffer that is administered by IM injection in the upper arm deltoid muscle of the non-dominant side. Physiochemically, the product is a dispersion of nucleic acid/lipid nanoparticles with an average size of approximately 70 to 90 nm.

A description of the vaccine and control (placebo) is presented in Table 7.1.

**Table 7.1 Vaccine and Placebo**

| Vaccine | Dosage Form | Dose | Manufacturer |
|---|---|---|---|
| PTX-COVID19-B mRNA Humoral Vaccine | Injectable solution, IM vaccine | 0.2 mg/mL 0.5 mL, at dose levels presented in Section 5.1 | Genevant Sciences Corporation |
| Control (Placebo) Vaccine (0.9% Normal Saline) | Injectable solution, IM vaccine | 0.0 mg/mL 0.5 mL | Commercially available source |

Abbreviations: IM = intramuscular; mRNA = messenger ribonucleic acid

Vaccine and placebo will be prepared by an unblinded site pharmacist. Vaccine will be sourced by Providence Therapeutics Holdings Inc. Saline may be sourced by the clinical site. Instructions for dilutions will be included in a Pharmacy Manual for the site.

#### 7.2 Treatments Administered

This study will initially be a first-in-human Phase I study of 3 dose levels (16, 40, and 100 µg). Two injections of the vaccine will be administered at an interval of 4 weeks. (refer to Section 5.1).

Detailed direction for vaccine preparation will be provided in a Pharmacy Manual.

#### 7.3 Method of Assigning Subjects to Treatment Groups

ICON’s unblinded statistician will prepare the randomization scheme in accordance with its SOPs and the randomization plan, which reflect GCP standards. Refer to Section 10.3 for a description of randomization methods. Eligible subjects will be assigned to cohorts according to age and the list of subject randomization assignments.

#### 7.4 Measurements of Treatment Compliance

All vaccine doses will be prepared and administered by delegated and trained staff at the clinical site. Details regarding dose preparation and dosing, including the dose administered and the date and time of dosing, will be recorded.

#### 7.5 Vaccine Storage, Accountability, and Retention

##### 7.5.1 Storage Conditions

The vaccine and placebo control storage temperatures will be presented in the Pharmacy Manual. Stability testing in ongoing.

The investigator will ensure that all the vaccine components are stored per the instructions accompanying the vaccine. In addition, the investigator will ensure that vaccine components are handled in accordance with the Health Canada regulations concerning the storage and administration of investigational products.

Refer to the SRM for additional vaccine component storage instructions.

##### 7.5.2 Vaccine Accountability and Retention

The investigator must ensure that all vaccine supplies are kept in a secure locked area with access limited to those authorized by the investigator. The investigator must maintain accurate records of the receipt of all vaccine/placebo shipped by Providence Therapeutics Holdings Inc. or their representative, including but not limited to the date received, lot number, expiration date, amount received, and the disposition of all vaccine. Current preparation and dispensing records will also be maintained including the date and amount of: complete vaccine dispensed, and the subject receiving the vaccine. All remaining vaccine not required by regulations to be held by the clinical site must be returned to Providence Therapeutics Holdings Inc. or their representative immediately after the study is completed.

#### 7.6 Packaging and Labeling

##### 7.6.1 Vaccine

The vaccine product is presented as a 0.2 mg/mL, 2 mL fill in a 3 mL United States Pharmacopeia / European Pharmacopeia Type I borosilicate glass vial with a fluoro-resin laminated bromobutyl rubber stopper and an aluminum overseal with a red, plastic, flip-off cap.

##### 7.6.2 Placebo

The placebo is commercially available sodium chloride 0.9%.

#### 7.7 Blinding of Treatment Assignment

ICON’s unblinded statistician will maintain the randomization code in a secure location with controls to prevent unauthorized access, including the computer program written to generate the randomization, randomization codes, program log, seed number used by the program, copy of the randomization plan along with approval documentation as appropriate, and the write-protected electronic storage medium.

ICON’s unblinded statistician will provide the randomization list to the site’s unblinded pharmacists. If the placebo and vaccine are not identical in appearance, the staff member administering the doses will be unblinded and will not participate in any subject assessments. In order to preserve the blind, vaccine will be prepared and administered in a manner that masks the content for both the subject and any observers. Investigators, site staff, subjects, CRO, and the sponsor will remain blinded to individual subjects' treatment assignment for the duration of the study until the study unblinding has been authorized.

###### 7.7.1 Unblinding of Treatment Assignment for Adverse Event/Safety Reasons

Should an SAE or other circumstance require that the blind be broken to ensure subject safety, the investigator must immediately notify the medical monitor and/or sponsor. ICON/designee will notify the sponsor as soon as possible and will release the treatment assignment of the subject to the appropriate personnel in accordance with the applicable SSP.

If the treatment assignment is unblinded by the site pharmacist or any party other than ICON, the investigator must notify ICON in writing and document the course of events in the source records. Any subject for whom the treatment code is prematurely released will be withdrawn from the study.

###### 7.7.2 Unblinding for Interim Analysis

At approximately 5 weeks after the last subject receives their last dose, there will be additional interim reviews by the study team and analysis of all data collected for safety and immunogenicity endpoints. The available data will be unblinded, cleaned, the database will be frozen, and a clinical study report will be written.

##### 7.8 Concomitant Medications and Procedures and Other Restrictions

###### 7.8.1 Concomitant Medications and Procedures

Any medication taken by a subject from 30 days prior to the screening visit through Day 42 and the reason for its use will be documented. After Day 42, concomitant medications will be documented if associated with an SAE, AESI, medically-attended AE, NOCD, or PIMMC.

Previous prescription medication for mild stable chronic health disorders (e.g., hypertension or hypercholesterolemia) or maintenance (e.g., contraception) will be allowed at the discretion of the investigator as long as the subject has been on a stable dose with no disease exacerbations for at least 3 months. Subjects should continue their regular maintenance medications throughout study participation unless instructed to discontinue or change by the investigator or their personal physician. If concomitant medications are discontinued by a subject during study conduct the subject should notify the investigator. Subjects should refrain from starting new medications, modifying current concomitant medications, or receiving vaccinations during study participation unless prescribed by the investigator for treatment of specific clinical events.

Subjects should not receive any vaccine within 1 month before screening except for influenza vaccine that should not be given within 14 days of study vaccine.

Subjects should not be using prescription medications for the prophylaxis of SARS-CoV-2.

Subjects should not participate or plan to participate in a study that involves an experimental agent (vaccine, drug, biologic, device, or medication); or should not have received an experimental agent within 1 month before enrollment in this study; or who expect to receive another experimental agent during participation in this study.

###### 7.8.2 Other Restrictions

Subjects will be instructed to adhere to the following restrictions:

- Strenuous activity (as assessed by the investigator) is prohibited from 72 hours prior to the days safety laboratory blood will be collected
- Subjects are not permitted to consume alcohol for 72 hours prior to each vaccination and should refrain from consumption of alcohol until 48 hours post vaccination.
- Subjects should refrain from blood donation for 6 months post vaccination.
- Subjects should not participate or plan to participate in a study that involves an experimental agent (vaccine, drug, biologic, device, or medication); or should not have received an experimental agent within 1 month (3 months for immunoglobulins) before enrollment in this study; or who expect to receive another experimental agent during participation in this study.

### 8 STUDY ASSESSMENTS AND PROCEDURES

Subjects will undergo study procedures and assessments at time points specified in the schedule of assessments (Table 2.1).

#### 8.1 Pandemic Safety Procedures

Appropriate measures will be taken to ensure the safety of both study staff and study subjects including social distancing and appropriate personal protective equipment.

#### 8.2 Medical and Surgical History

The investigator or designee will collect a complete medical and surgical history at screening. As applicable, self-reported medical and surgical history will be collected at clinical site visits to determine if any changes have occurred since screening. Any ongoing chronic conditions must be stable with medication changes and no exacerbations in past 3 months and no hospitalizations in past 6 months.

#### 8.3 Demographic Characteristics

Demographic characteristics including sex, age, race, and ethnicity will be recorded.

#### 8.4 Physical Measurements

Height (cm) and body weight (kg) without shoes will be recorded. Body mass index will be calculated using the height obtained at screening.

#### 8.5 Immunogenicity Assessments

Blood samples or immunogenicity analysis will be collected as per Table 2.1. Immunogenicity assessments will include the following:

- Antibodies:
  - Anti-COVID-19 , IgG, IgA by ELISA
  - Anti-COVID-19 neutralizing antibody titer assays
- Cell-mediated Immunity/PBMC Evaluations (Flow Cytometry,
- Additional assays may be conducted for research purposes.

Immunogenicity sample collection, processing, and shipping details will be outlined in a separate SRM.

#### 8.6 Safety Assessments

##### 8.6.1 Adverse Events

Subjects will be monitored for solicited and unsolicited AEs according to Section 9.

If a subject experiences respiratory symptom (e.g., primary respiratory fever or initially presented symptoms) or has a significant exposures to someone with confirmed SARS-CoV2 infection, the subject should notify the clinical site for a visit to collect NP swabs for RT-PCR virus detection. If positive, the subject will be referred for standard of care and followed for severity, duration (number of days from date of first symptom[s] onset to 2 days without symptoms), and outcome.

For SAEs, medically-attended AEs, NOCDs, PIMMCs, and AESIs, the clinical site will attempt to obtain medical records from the subjects' care providers.

##### 8.6.2 Reactogenicity Assessments

Subjects will keep a Diary Card to track reactogenicity and other safety assessments for 7 days after each vaccination. Reactogenicity solicited events include, generalized symptoms such as headache, fatigue, muscle pain, joint pain, nausea, chills, and fever, and localized reactions including injection site pain, bruising, redness, and swelling. Subjects will be asked to record oral temperature with their own thermometer if they feel any of the above symptoms and record in the Diary Card.

##### 8.6.3 Laboratory Tests

A certified laboratory will be utilized to process and provide results for the clinical safety laboratory tests listed in Table 8.1. The baseline laboratory test results for clinical assessment for a particular test will be defined as the last measurement prior to the first dose of vaccine.

Subjects will refrain from strenuous exercise for 48 hours prior to laboratory sample collection and from alcohol for 72 hours prior to laboratory sample collection. Subjects will be non-fasted at screening; if glucose test result is outside the normal range then one fasted repeat test may be performed.

During screening, if a subject has an out-of-range value for a clinical safety laboratory parameter that the investigator believes is not clinically significant or the investigator does not believe is correct (e.g., lab or specimen processing error), but the investigator wants to confirm with a repeat laboratory test, a single repeat is allowed to confirm the initial result.

Additional safety laboratory tests may be conducted as needed by the investigator to evaluate subject safety.

**Table 8.1 Clinical Laboratory Tests**

| Hematology | Chemistry | Urinalysis |
|---|---|---|
| HbA1C (calculation) | Alanine aminotransferase | Blood |
| Hemoglobin | Albumin | Glucose |
| Platelet count (estimate not acceptable) | Alkaline phosphatase | Ketones |
| Red blood cell count | Aspartate aminotransferase | Leukocyte esterase |
| White blood cell count with total and differential: | Gamma-Glutamal Transferase | Microscopic analysis (performed if blood, leukocytes, or protein are present) |
| Neutrophils | Blood urea nitrogen | pH |
| Lymphocytes | Creatinine | Protein |
| Monocytes | Direct bilirubin | Specific gravity |
| Basophils | Glucose | |
| Eosinophils | Total bilirubin | |
| | Total protein | |

For any laboratory test value outside the reference range that the investigator considers clinically significant during the on-study period (i.e., following either vaccine administration), the investigator will:

- Repeat the test to verify the out-of-range value and clinical significance.
- Follow the out-of-range value until the value returns to normal or baseline, or until the value is deemed stable and not clinically significant by the investigator.
- Record as an AE any laboratory test value that is confirmed by repeat and the investigator considers clinically significant, requires a subject to be discontinued from the study, requires a subject to receive treatment, or requires a change or discontinuation of the vaccine (if applicable).

##### 8.6.4 Other Tests

The following tests will be performed:

- Urine drugs of abuse (at a minimum, opiates, cocaine, phencyclidine, amphetamines, benzodiazepines, and methadone) screens
  - During the screening period, urine drugs of abuse screens may not be repeated for eligibility
  - An alcohol breathalyzer test will be performed at screening
- Serology tests (i.e., HIV-1 and HIV-2 antibodies, HBsAg, and HCV antibody, and any confirmatory tests performed at the discretion of the investigator)
- Urine pregnancy test (females only)
- Immunogenicity analysis and cell mediated immunity (refer to Section 8.5)

- NP swab for COVID-19 RT-PCR (the clinic also has the option to confirm eligibility via an additional point-of-care test)

###### 8.6.5 Vital Signs

Vital signs assessments will include systolic and diastolic blood pressure (mmHg), heart rate (bpm), respiratory rate (breaths per minute), and oral temperature (°C). Pulse oximetry (%) will be performed prior to each vaccination and if applicable for unscheduled visits for COVID-like illness. Vital signs will be measured after the subject has been resting quietly in a seated position for at least 5 minutes. Any clinically significant abnormal vital sign assessment requires at least one repeat measurement. Any subject with a temperature of <35C or >37.5 or pulse oximetry of <90% after repeat measurement, will not be vaccinated and will be rescheduled at the discretion of the investigator.

A vital sign abnormality that is considered clinically significant initially and on confirmation, requires a subject to be discontinued from the study, requires a subject to receive treatment, or requires discontinuation from the vaccine (if applicable) will be recorded as an AE.

###### 8.6.6 Physical Examination

Complete physical examinations (excluding genital, rectal, and breast examinations [unless indicated]) will be performed at screening. Targeted physical examinations that are symptom-directed will be performed at other time points as per Table 2.1. Abnormal findings will be documented in the subject's eCRF.

An abnormal physical examination finding that is considered clinically significant and requires the subject to be discontinued from the study, requires the subject to receive treatment, or requires discontinuation of the vaccine (if applicable) will be recorded as an AE.

###### 8.6.7 Appropriateness of Safety Assessments

Safety evaluations selected for this study are typical of those for this subject population and utilize widely accepted measures.

###### 8.6.8 Resuscitation

The site will have readily available resuscitation equipment including but not limited to epinephrine, oxygen, automated defibrillator, and intravenous fluids and catheters for volume expansion. A physician (or appropriately trained personnel such as a paramedic) must be physically present and available for all injections and for 60 minutes after each vaccination for all patients.

### 9 ADVERSE EVENTS

An AE is defined as any untoward medical occurrence in a subject administered a pharmaceutical product during the course of a clinical investigation. An AE can therefore be any unfavorable and unintended sign, symptom, or disease temporally associated with the use of a vaccine, whether or not thought to be related to the vaccine.

Medically-attended AEs are AEs for which the subject has received medical attention by medical personnel, or in an emergency room, or which led to hospitalization. For each reported AE, the investigator will ask the subject if such medical attention has been received. In addition, the investigator will assess each medically-attended AE if it constitutes a NOCD, defined as a medically-attended AE that was:

- absent at baseline,
- not resolved at EOS
- required continuous medical care or attention.

Assessments of unsolicited AEs will be performed on Day 1 through Day 42. Medically-attended AE assessments will be performed when subjects are in-house on Days 1 through 42. Assessments of NOCD, AESIs (including COVID-19 cases for enhanced disease), and PIMMCs will be performed from Day 1 through the end of study as per Table 2.1. The SAEs will be assessed throughout the study (refer to Table 2.1). Adverse events that are identified at the last assessment visit (or the early termination visit) as specified in the protocol must be recorded on the AE eCRF with the status of the AE noted. All events that are ongoing at this time will be recorded as ongoing on the eCRF. All (both serious and nonserious) AEs must be followed until they are resolved or medically stable, or until reasonable attempts to determine resolution of the event are exhausted. The investigator should use his/her discretion in ordering additional tests as necessary to monitor the resolution of such events.

The procedures specified in Section 9.4 are to be followed for reporting SAEs.

#### 9.1 Recording Adverse Events

Adverse events are to be recorded on the AE page of the eCRF. The following information will be recorded:

- Assessment of whether or not the AE is an SAE, medically-attended AE, NOCD, PIMMC, or AESI (Section 9.2.1)
- Assessment of AE intensity (Section 9.2.2)
- Assessment of AE relationship to vaccine (Section 9.2.4)
- Action taken - dose not changed, vaccine dose 2 not given, not applicable, or unknown, as applicable
- Outcome - recorded as fatal, not recovered/not resolved, recovered/resolved, recovered/resolved with sequelae, recovering/resolving, or unknown, as applicable
- Solicited AEs from the diary card will be assumed to be possibly related due to temporal association. Local reactions will be assumed to be definitely related unless an alternative plausible cause is identified.

#### 9.2 Assessment of Adverse Events

The investigator will assess each AE except solicited events for seriousness, intensity, and relationship to vaccine.

##### 9.2.1 Serious Adverse Events, Medically Attended Adverse Events, and New Onset Chronic Disease, Adverse Events of Special Interest, and Potential Immune-mediated Conditions

The investigator is responsible for determining whether an AE meets the definition of an SAE, medically-attended AE, NOCD, AESI, or PIMMC per the timing presented in Table 2.1.

An SAE is any AE that results in any of the following outcomes:

- Death
- A life-threatening adverse drug experience
- Inpatient hospitalization or prolongation of existing hospitalization
- A persistent or significant disability/incapacity
- A congenital anomaly/birth defect

Important medical events that may not result in death, be life threatening, or require hospitalization may be considered an SAE when, based upon appropriate medical judgment, they may jeopardize the subject and may require medical or surgical intervention to prevent any of the outcomes listed in this definition. Examples of such medical events include allergic bronchospasm requiring intensive treatment in an emergency room or at home, blood dyscrasias or convulsions that do not result in inpatient hospitalization, or the development of drug dependency or drug abuse.

Medically-attended AEs are defined as hospitalization, an emergency room visit, or an otherwise unscheduled visit to or from medical personnel for any reason except for a health maintenance visit for a routine physical.

New onset chronic diseases are defined as any new ICD diagnosis (per current WHO International Statistical Classification of Diseases and Related Health Problems) that is applied to the subject during the course of the study, after receipt of the study agent, that is expected to continue for at least 3 months and requires continued health care intervention.

Adverse events of special interest are defined in Section 9.2.2 and a list of PIMMCs is provided in Appendix B.

Note: SAEs, medically-attended AEs, NOCDs, AESIs, and PIMMCs require immediate reporting to the medical monitor and/or sponsor. Refer to Section 9.4 for details.

##### 9.2.2 Adverse Events of Special Interest and Serious and Unexpected Suspected Adverse Reactions

An AESI is any AE that a regulatory authority has mandated be reported on an expedited basis, regardless of the seriousness, expectedness, or relatedness of the AE to the administration of investigational product. A SUSAR is considered “unexpected” if it is not listed in the Investigator Brochure or is not listed at the specificity or severity that has been observed; or, if an Investigator Brochure is not required or available, is not consistent with the risk information described in the general investigational plan or elsewhere in the current application.

To date, no AESIs have been identified specifically for this vaccine. However, subjects who test positive for SARS-CoV-2, present with autoimmune disease, or are diagnosed with a PIMMC (Appendix B) should be reported to ICON Pharmacovigilance (Section 9.4) as an AESI or SUSAR. The subject will be referred to their primary care provider for COVID-19 standard of care. The AE will be followed appropriately, including illness duration, severity, and outcome.

##### 9.2.3 Intensity

The intensity of an AE will be graded according to Guidance for Industry: Toxicity Grading Scale for Healthy Adult and Adolescent Volunteers Enrolled in Preventive Vaccine Clinical Trials (September 2007). The grading scale is comprised of 4 levels: mild (Grade 1), moderate (Grade 2), severe (Grade 3), and potentially life-threatening (Grade 4). For those AEs that are not covered in the Guidance for Industry (above) the rating scale from DAIDS (Table for Grading the Severity of Adult and Pediatric Adverse Events, Version 2.1) will be used (Table 9.1).

**Table 9.1 Division of AIDS Adverse Events Intensity Assessment**

| Grade | Intensity | Definition |
|---|---|---|
| Grade 1 | Mild | Symptoms causing no or minimal interference with usual social and functional activities with intervention not indicated. |
| Grade 2 | Moderate | Symptoms causing greater than minimal interference with usual social and functional activities with intervention indicated. |
| Grade 3 | Severe | Symptoms causing inability to perform usual social and functional activities with intervention or hospitalization indicated. |
| Grade 4 | Potentially Life-Threatening | Symptoms causing inability to perform basic self-care functions with intervention indicated to prevent permanent impairment, persistent disability, or death. |
| Grade 5 | | Any adverse event where the outcome is death |

##### 9.2.4 Relationship to Vaccine

The relationship of an AE to the vaccine should be determined by the investigator according to the following criteria:

- Not related: The event is most likely produced by other factors such as the subject’s clinical condition, intercurrent illness, or concomitant drugs, and does not follow a known response pattern to the vaccine, or the temporal relationship of the event to vaccine administration makes a causal relationship unlikely
- Related: The event follows a reasonable temporal sequence from the time of drug administration, and/or follows a known response pattern to the vaccine, and cannot be reasonably explained by other factors such as the subject's clinical condition, intercurrent illness, or concomitant drugs

#### 9.3 Management and Discontinuation Due to Adverse Events

Any subject who experiences an AE may be withdrawn at any time from the study at the discretion of the investigator. Subjects withdrawn from the study due to an AE, whether serious or nonserious, may be followed by the investigator until the clinical outcome of the AE is determined. The AE(s) should be noted on the appropriate eCRFs, the subject’s progress should be followed until the AE is resolved or medically stable as determined by the investigator, and the sponsor notified. Subjects testing positive or develop symptoms related to infection with SARS-CoV2 during the study will be followed for severity, duration, and outcome, and will be followed for safety and immune response to the end of the study period. Subjects with ongoing or chronic symptoms attributable to COVID-19 at the end of the study, will be referred to their regular physician for follow up. Subjects testing positive or develop symptoms related to infection with SARS-CoV2 after the first dose of the vaccine but before the second dose, will not be given any further doses of the study medication but will be followed to the end of the study.

#### 9.4 Reporting Serious Adverse Events, Medically-Attended Adverse Events, New Onset Chronic Disease, Adverse Events of Special Interest, and Potential Immune-Mediated Medical Conditions

In the event of any SAE, MAAE, NOCD, AESI, or PIMMC reported or observed during the study, whether or not attributable to the study vaccine, site personnel will report it immediately by telephone to the sponsor, medical monitor, AND SAE hotline at ICON (ICON Pharmacovigilance) (SAE/AESI/pregnancy should be reported to or **Fax to** +44 (0) 208 100 5005, not to SAE hotline) in accordance with procedures described in the SRM and/or SSP. Site personnel will follow up with a written report on the next working day:Report Forms will be provided to the CRU to assist in collecting, organizing, and reporting SAEs, MAAEs, NOCDs, PIMMCs, or AESIs and follow-up information.

All SAEs, MAAEs, NOCDs, PIMMCs, and AESIs should be followed to their resolution, with documentation provided to the sponsor/ICON on a follow-up Report Form.

#### 9.5 Pregnancy

Pregnancies will be captured if they occur in female subjects from the time the subject is first exposed to the vaccine until end of study. Pregnancies in the sexual partners of male subjects will be captured from the time the subject is first exposed to the vaccine until 90 days after last exposure to the vaccine.

Female subjects must be instructed to inform the study investigator immediately if they become pregnant during the study.

The investigator must report any pregnancy to the medical monitor, sponsor, and ICON Pharmacovigilance within 1 business day of becoming aware of the pregnancy per pregnancy reporting procedures described in the SRM. The subject must be immediately discontinued from further treatment with vaccine. An uncomplicated pregnancy will not be considered an AE or SAE; however, all pregnancies will be followed through birth and 3 months post-delivery.

Any congenital abnormalities in the offspring of a subject who received vaccine will be reported as an SAE. The outcome of any pregnancy and the presence or absence of any congenital abnormality will be recorded in the source documentation and reported to the sponsor and ICON Pharmacovigilance.

#### 9.6 Drug-induced Liver Injury

Subjects will be monitored for signs of DILI. Vaccine (dose 2) will be withheld in the event of potential DILI.

Potential events of DILI will be defined as meeting all of the following criteria (as specified in the Health Canada Guidance Document: Pre-market Evaluation of Hepatotoxicity in Health Products, 2012):

- Alanine aminotransferase or aspartate aminotransferase > 3 × ULN
- Total bilirubin > 2 × ULN without initial findings of cholestasis (elevated serum alkaline phosphatase)
- No other reason can be found to explain the combination of laboratory value increases (e.g., acute viral hepatitis; alcoholic and autoimmune hepatitis; hepatobiliary disorders; nonalcoholic steatohepatitis; cardiovascular causes; concomitant treatments)

Potential events of DILI will be reported as SAEs (Section 9.4). All subjects with potential DILI will be closely followed until abnormalities return to normal or baseline or until reasonable attempts to determine resolution of the event are exhausted.

### 10 STATISTICAL CONSIDERATIONS

The statistical analysis will be conducted following the principles as specified in ICH Topic E9 (CPMP/ICH/363/96).

All statistical analyses will be described in a separate SAP.

The statistical evaluation will be performed using SAS® software version 9.4 or higher (SAS Institute, Cary, NC). All data will be listed, and summary tables will be provided as applicable. No formal significance testing will be performed. Summary statistics will be presented by cohort, timepoint, treatment (as applicable) and phase. For continuous variables, data will be summarized with the number of subjects, mean, standard deviation, median, minimum, maximum, and 95% confidence interval by treatment group. For categorical variables, data will be tabulated with the number and proportion of subjects for each category by treatment group.

The database will be locked and unblinded after the data have been monitored for all subjects at approximately 5 weeks after the last subject receives their last dose and then again after all subjects complete the study.

The final analysis of the safety and immunogenicity data collected up to approximately 1 year after the last vaccination will be performed after the study is completed. An addendum to the initial CSR will be prepared with data from final analysis. Details of the statistical analyses, methods, and data conventions will be described in the SAP.

#### 10.1 Sample Size Calculation

No formal sample size calculation was performed.

A total of 60 adult subjects will be enrolled and randomized per Section 10.3.

Rare AEs are not demonstrable in a clinical study of this size; however, the probabilities of observing one or more AEs given various true event rates are presented in Table 10.1. With the assumption that all enrolled subjects will likely complete immunizations and safety visits in this relatively short duration study, the following statistical considerations apply. With 15 subjects in each dose group, the chance of observing at least 1 AE of probability ≥ 20% is approximately 97%. Therefore, if no AEs of a given type occur in a dose cohort, it is relatively certain that they will occur in < 20% of people once the vaccine is implemented. With 45 subjects across the 3 Phase 1dosing cohorts, the chance of observing at least 1 AE of probability ≥ 5% is at least 90%. Therefore, if no AEs of a given type occur across the combined doses, it is relatively certain that any dosage-independent event will occur in < 5% of people once the vaccine is implemented.

**Table 10.1 Probability of Observing an Adverse Event for Various Event Rates**

| N | "True" Event Rate | Probability of Observation (%) | N | "True" Event Rate | Probability of Observation (%) |
|---|---|---|---|---|---|
| 15 | 0.1% | 1.5 | 45 | 0.1% | 4.4 |
| | 0.5% | 7.2 | | 0.5% | 20.2 |
| | 1.0% | 14.0 | | 1.0% | 36.4 |
| | 2.0% | 26.1 | | 2.0% | 59.7 |
| | 3.0% | 36.7 | | 3.0% | 74.6 |
| | 4.0% | 45.8 | | 4.0% | 84.1 |
| | 5.0% | 53.7 | | 5.0% | 90.1 |
| | 10.0% | 79.4 | | 10.0% | 99.1 |
| | 15.0% | 91.3 | | 15.0% | 99.9 |
| | 20.0% | 96.5 | | 20.0% | >99.9 |

#### 10.2 Analysis Populations

Safety Population: All subjects who provide consent, are randomized, and receive any amount of vaccine/placebo. The Safety Population will be used for all safety analyses will be analyzed as actually treated.

Per Protocol Population: Includes all subjects in the Safety Population who receive the assigned doses of the vaccine/placebo according to protocol, have serology results, and have no major protocol deviations affecting the primary immunogenicity outcomes, as determined by the Sponsor before database lock and unblinding. The Safety Population will be the primary population used for the analysis of safety endpoints.

Modified Intent-to-Treat Population: The mITT Population will be used for the analysis of immunogenicity endpoints. Analysis using the mITT Population will not be performed if it differs from the PP Population by ≤ 5% of the subjects for each of the treatment groups.

#### 10.3 Randomization

On Day 1, randomization will be performed. Three cohorts of 20 subjects each will include sentinel groups of 5 subjects randomized in 4:1 ratio for investigational vaccine:placebo, followed by a cohort expansion phase of 15 subjects randomized in 11:4 ratio for investigational vaccine:placebo.

The randomization list containing a list of vaccine or placebo assignment will be prepared by ICON's randomization team.

#### 10.4 Endpoints

Refer to Section 4.2.

#### 10.5 Immunogenicity Analysis

#### 10.6 Immunogenicity data for each cohort and study phase will be listed and summarized by cohort, and time point (Table 2.1) using descriptive statistics as mentioned in Section 10 along with geometric mean and 95% confidence interval. Safety Analysis

Vital signs, clinical laboratory tests, and physical examination findings will be listed and summarized by cohort, treatment (as applicable), and time point (Table 2.1) using appropriate descriptive statistics.

The number and percentage of subjects reporting any TEAE or reactogenicity will be summarized by study phase and cohort and tabulated by system organ class and preferred term (coded using MedDRA). The TEAEs will be further classified by severity and relationship and for SAEs, medically-attended AEs, NOCD, PIMMCs, and AESIs.

Additionally, number of subjects who became infected after vaccination and whether being vaccinated made the disease less or more severe will be presented.

### 11 ACCESS TO SOURCE DATA/DOCUMENTS

The investigator will provide direct access to source data and documents for individuals conducting study-related monitoring, audits, IRB/IEC review, and regulatory review. The investigator must inform the study subject that his/her study-related records may be reviewed by the above individuals without violating the subject's privacy of personal health information in compliance with regulations.

By signing this protocol, the investigator affirms to the sponsor that the investigator will maintain, in confidence, information furnished to him or her by the sponsor and will divulge such information to the IRB/IEC under an appropriate understanding of confidentiality with such board.

### 12 QUALITY CONTROL AND QUALITY ASSURANCE

Sponsor/ICON will implement and maintain quality control and quality assurance procedures with written SOPs to ensure the study is conducted and data are generated, documented, and reported in compliance with the protocol, GCP, and applicable regulatory requirements.

#### 12.1 Conduct of Study

This study will be conducted in accordance with the provisions of the Declaration of Helsinki and all revisions thereof (Tokyo 2004), and in accordance with ICH E6 Guidelines on GCP (CPMP/ICH/135/95). Specifically, this study is based on adequately performed laboratory and animal experimentation; the study will be conducted under a protocol reviewed by an IRB/IEC; the study will be conducted by scientifically and medically qualified persons; the benefits of the study are in proportion to the risks; the rights and welfare of the subjects will be respected; the physicians conducting the study do not find the hazards to outweigh the potential benefits; and each subject will give his or her written, informed consent before any protocol-driven tests or evaluations are performed.

The investigator may not deviate from the protocol without a formal protocol amendment having been established and approved by an appropriate IRB/IEC, except when necessary to eliminate immediate hazards to the subject or when the change(s) involve only logistical or administrative aspects of the study and are approved by the sponsor. Any deviation may result in the subject having to be withdrawn from the study, and may render that subject nonevaluable.

##### 12.1.1 Protocol Deviations

A protocol deviation is defined as any intentional or unintentional change to, or noncompliance with, the approved protocol procedures or requirements. Deviations may result from the action or inaction of the subject, investigator, or site staff.

At the outset of the study, a process for defining and handling protocol deviations will be established. This will include determining which violations will be designated “key”, requiring immediate notification to the medical monitor and/or sponsor. The investigator is responsible for seeing that any known protocol deviations are recorded and handled as agreed.

#### 12.2 Protocol Amendments

Only the sponsor may modify the protocol. Amendments to the protocol will be made only after consultation and agreement between the sponsor and the investigator. All amendments that have an impact on subject risk or the study objectives, or require revision of the ICF, must receive approval from the IRB/IEC prior to their implementation.

#### 12.3 Monitoring of Study

The investigator will permit the CRA to review study data as frequently as is deemed necessary to ensure data are being recorded in an adequate manner and protocol adherence is satisfactory.

The investigator will provide access to medical records for the monitor to verify eCRF entries. The investigator is expected to cooperate with the sponsor or a designee in ensuring the study adheres to GCP requirements.

The investigator may not recruit subjects into the study until the sponsor or a designee has conducted a site initiation visit (in-person or via teleconference) to review the protocol and eCRF in detail.

### 13 ETHICS

#### 13.1 Institutional Review Board/Independent Ethics Committee Approval

##### 13.1.1 Ethics Review Prior to Study

The investigator will ensure that the protocol and ICF are reviewed and approved by the appropriate IRB/IEC prior to the start of any study procedures. The IRB/IEC will be appropriately constituted and will perform its functions in accordance with Health Canada regulations, ICH GCP guidelines, and local requirements as applicable.

##### 13.1.2 Ethics Review of Other Documents

The IRB/IEC will approve all protocol amendments (except for sponsor-approved logistical or administrative changes), written informed consent documents and document updates, subject recruitment procedures, written information to be provided to the subjects, available safety information, information about payment and compensation available to subjects, the investigator’s curriculum vitae and/or other evidence of qualifications, and any other documents requested by the IRB/IEC and regulatory authority as applicable.

#### 13.2 Written Informed Consent

The nature and purpose of the study will be fully explained to each subject. The subjects must be given ample time and opportunity to inquire about details of the study, to have questions answered to their satisfaction, and to decide whether to participate. Written informed consent must be obtained from each subject prior to any study procedures being performed.

### 14 DATA HANDLING AND RECORD KEEPING

#### 14.1 Data Reporting and Case Report Forms

##### 14.1.1 Case Report Forms

The investigator will be provided with eCRFs, and will ensure all data from subject visits are promptly entered into the eCRFs in accordance with the specific instructions given. The investigator must sign the eCRFs to verify the integrity of the data recorded.

##### 14.1.2 Laboratory Data

A list of the normal ranges for all laboratory tests to be undertaken forms part of the documentation to be collated prior to study start. If a central laboratory has been selected to conduct any or all tests, it is essential all samples be analyzed at that laboratory. The investigator must maintain source documents such as laboratory reports and complete history and physical examination reports.

##### 14.1.3 Retention of Source Documents

The investigator must maintain source documents such as laboratory reports, x-rays, consultation reports, and complete history and physical examination reports.

#### 14.2 Retention of Essential Documents

The study essential documents must be maintained as specified in the ICH guidelines for GCP and the applicable regulatory requirements. The investigator/institution should take measures to prevent accidental or premature destruction of these documents.

Essential documents should be retained until at least 2 years after the last approval of a marketing application in an ICH region and until there are no pending or contemplated marketing applications in an ICH region or until at least 2 years have elapsed since the formal discontinuation of clinical development of the vaccine. These documents should be retained for a longer period; however, if required by the applicable regulatory requirements or by an agreement with the sponsor. It is the responsibility of the sponsor to inform the investigator/institution as to when these documents no longer need to be retained.

### 15 ADMINISTRATIVE INFORMATION

#### 15.1 Financing and Insurance

Financing and insurance will be addressed in a separate agreement between the sponsor and the investigator.

#### 15.2 Publication Policy

The sponsor will retain ownership of all data. All proposed publications based on this study will be subject to sponsor's approval requirements.

PTX-COVID19-B Vaccine Investigator's Brochure. Providence Therapeutics Holdings. 02 December 2020.

National Heart, Lung, and Blood Institute, Calculate Your Body Mass Index. Available at: https://www.nhlbi.nih.gov/health/educational/lose_wt/BMI/bmi-m.htm. Last accessed 15 October 2020.

WHO Coronavirus Disease (COVID-19) Dashboard. Available at: https://covid19.who.int/. Last accessed 14 October 2020.

WHO International Statistical Classification of Diseases and Related Health Problems. Available at: https://www.who.int/classifications/icd/en/. Last accessed 15 October 2020.

Centers for Disease Control and Prevention. Coronavirus Disease 2019 (COVID-19). Available at https://www.cdc.gov/coronavirus/2019-ncov/need-extra-precautions/people-with-medical-conditions.html?CDC_AA_refVal=https%3A%2F%2Fwww.cdc.gov%2Fcoronavirus%2F2019-

ncov%2Fneed-extra-precautions%2Fgroups-at-higher-risk.html. Last accessed 16 December 2020.

#### APPENDIX A DIARY CARD EXAMPLE

Example Diary Card may be adjusted to add additional subject information and/or clinical site information.

VACCINE REACTION AND ADVERSE EVENT DIARY

**Section 1: Diary - Reactogenicity Days 1-8**

My Subject ID number is: ____________

| **Date:** | | | | | | | | |
|---|---|---|---|---|---|---|---|---|
| **Day of immunization►** / **Symptom** | Evening of Dose | 1 Day after | 2 Days after | 3 Days after | 4 Days after | 5 Days after | 6 Days after | 7 Days after |
| **Body temperature** **Celsius (C) or Fahrenheit (F)** | | | | | | | | |
| **Fill in today's column by entering the worst grade/severity for each symptom that you have had in the last day (24 hours). Symptom grades are defined at the bottom of the page.** | | | | | | | | |
| **Generalized Symptoms** | | | | | | | | |
| Headache | | | | | | | | |
| Fatigue | | | | | | | | |
| Muscle Pain | | | | | | | | |
| Joint pain | | | | | | | | |
| Nausea | | | | | | | | |
| Chills | | | | | | | | |
| Feverish | | | | | | | | |
| Rash (not at injection site) | | | | | | | | |
| **Local Reactions** | | | | | | | | |
| Pain | | | | | | | | |
| Erythema / redness (in mm) | | | | | | | | |
| Induration (in mm) | | | | | | | | |
| Swelling (in mm) | | | | | | | | |
| **No Symptoms of Any Kind – Tick this box** | | | | | | | | |
| **In addition, tick the box in today's column if your answer to the question below is YES.** | | | | | | | | |
| **Any other new symptom(s)? Y or N. If yes, describe in section 2.** | | | | | | | | |

| | |
|---|---|
| **Grade 0 =** | I didn't have it at all. |
| **Grade 1 =** | I noticed it, but it didn't interfere with my usual activities at all. |
| **Grade 2 =** | I had it, and it was bad enough to prevent a significant part of my usual activities. |
| **Grade 3 =** | I had it, and it prevented most or all of my normal activities, or I had to see a doctor for prescription medicine. |
| **Grade 4 =** | I had to visit the ER or was hospitalized because of the specific symptom |

| VACCINE REACTION AND ADVERSE EVENT DIARY | | | | | |
|---|---|---|---|---|---|
| **Section 2:**<br>**Other Adverse Events Day 1 until next on-site visit**<br><br>Please describe any other symptoms experienced beyond those listed above. Please describe symptom or diagnosis, severity, start and stop day and if you saw a health care profession and severity below | | | | | |
| **Symptom or Diagnosis or if visited a Health Care Provider** | **Did you visit a care provider** | **Start Date** | **Stop Date** | **Severity[1]** | **Comments, including any medicine prescribed or taken to reduce the symptoms** |

| [1]**Please use these qualitative definitions to grade your symptoms** | |
|---|---|
| Grade 1 = | I noticed it, but it didn't interfere with my usual activities at all. |
| Grade 2 = | I had it, and it was bad enough to prevent a significant part of my usual activities. |
| Grade 3 = | I had it, and it prevented most or all of my normal activities, or I had to see a doctor for prescription medicine. |
| Grade 4 = | I had it and visited the ER or was hospitalized for it. |

**PLEASE BRING THIS DOCUMENT TO THE CLINIC AT YOUR NEXT VISIT. If you have any symptoms that rate Grade 3 or Grade 4, please call the clinic immediately. The clinic may ask you to send a picture of this diary via text or email.**

**Study Site Contact Information:**

| | |
|---|---|
| **Principal Investigator:** | **Ben Lasko, MD** |
| **Telephone:** | **416-740-2895**<br>**416-556-4399 (24hr-Emergency)** |
| **Study Coordinator:** | ______________________________ |
| **Email address:** | ______________________________ |

### APPENDIX B POTENTIAL IMMUNE-MEDIATED MEDICAL CONDITIONS

**Gastrointestinal Disorders**

- Autoimmune pancreatitis
- Celiac disease
- Crohn's disease
- Microscopic colitis
- Ulcerative colitis
- Ulcerative proctitis

**Liver Disorders**

- Autoimmune cholangitis
- Autoimmune hepatitis
- Primary biliary cirrhosis
- Primary sclerosing cholangitis

**Metabolic Diseases**

- Addison's disease
- Autoimmune hypophysitis
- Autoimmune thyroiditis (including Hashimoto thyroiditis)
- Diabetes mellitus type I
- Grave's or Basedow's disease

**Musculoskeletal Disorders**

- Antisynthetase syndrome
- Dermatomyositis
- Juvenile chronic arthritis (including Still's disease)
- Mixed connective tissue disorder
- Polymyalgia rheumatic
- Polymyositis
- Psoriatic arthropathy
- Relapsing polychondritis
- Rheumatoid arthritis
- Scleroderma, including diffuse systemic form and CREST (alcinosis, Raynaud's phenomenon, esophageal dysmotility, sclerodactyly, and telangiectasia) syndrome
- Spondyloarthritis, including ankylosing spondylitis, reactive arthritis (Reiter's Syndrome), and undifferentiated spondyloarthritis
- Systemic lupus erythematosus
- Systemic sclerosis

**Neuroinflammatory Disorders**

- Acute disseminated encephalomyelitis, including site-specific variants (eg, noninfectious encephalitis, encephalomyelitis, myelitis, radiculomyelitis)
- Cranial nerve disorders, including paralyses/paresis (eg, Bell’s palsy)
- Guillain-Barré syndrome, including Miller Fisher syndrome and other variants
- Immune-mediated peripheral neuropathies and plexopathies, including chronic inflammatory demyelinating polyneuropathy, multifocal motor neuropathy and polyneuropathies associated with monoclonal gammopathy
- Multiple sclerosis
- Narcolepsy
- Optic neuritis
- Transverse myelitis
- Myasthenia gravis, including Eaton-Lambert syndrome

**Skin Disorders**

- Alopecia areata
- Autoimmune bullous skin diseases, including pemphigus, pemphigoid and dermatitis herpetiformis
- Cutaneous lupus erythematosus
- Erythema nodosum
- Morphoea
- Lichen planus
- Psoriasis
- Sweet’s syndrome
- Vitiligo

**Vasculitides**

- Large vessels vasculitis including: giant cell arteritis such as Takayasu's arteritis and temporal arteritis
- Medium sized and/or small vessels vasculitis including the following:
  - Polyarteritis nodosa
  - Kawasaki’s disease
  - Microscopic polyangiitis
  - Wegener’ granulomatosis
  - Behcet’s syndrome
  - Leukocytoclastic vasculitis
  - Henoch-Schonlein purpura
  - Necrotizing vasculitis and antineutrophil cytoplasmic antibody (ANCA) positive vasculitis (type unspecified)
  - Churg-Strauss syndrome (allergic granulomatous angiitis)
  - Buerger’s disease (thromboangiitis obliterans)

**Others**

- Antiphospholipid syndrome
- Autoimmune hemolytic anemia
- Autoimmune glomerulonephritis (including IgA nephropathy, glomerulonephritis rapidly progressive, membranous glomerulonephritis, membranoproliferative glomerulonephritis, and mesangioproliferative glomerulonephritis)
- Autoimmune myocarditis/cardiomyopathy
- Autoimmune neutropenia
- Autoimmune pancytopenia
- Autoimmune thrombocytopenia
- Goodpasture syndrome
- Idiopathic pulmonary fibrosis
- Pernicious anemia
- Polyglandular autoimmune syndrome
- Raynaud's phenomenon
- Sarcoidosis
- Sjögren's syndrome
- Stevens-Johnson syndrome
- Uveitis

### 17 SIGNATURES

**Protocol Number: PRO-CL-001**

**Protocol Title: A Phase I, First-in-Human, Observer-Blinded, Randomized, Placebo-Controlled, Ascending Dose Study to Evaluate the Safety, Tolerability, and Immunogenicity of PTX-COVID19-B Vaccine in Healthy Seronegative Adults Aged 18-64**

**Providence Therapeutics Holdings Inc. Signatures**

This clinical study protocol has been reviewed and approved by Providence Therapeutics Holdings Inc.

| | |
|---|---|
| | 10 MAR 2021 |
| Jared Davis, President | Date |

**Protocol Number: PRO-CL-001**

**Protocol Title: A Phase I, First-in-Human, Observer-Blinded, Randomized, Placebo-Controlled, Ascending Dose Study to Evaluate the Safety, Tolerability, and Immunogenicity of PTX-COVID19-B Vaccine in Healthy Seronegative Adults Aged 18-64**

#### Investigator Signature

I agree to conduct the aforementioned study according to the terms and conditions of the protocol, GCP guidelines, and all other applicable local and regulatory requirements. All information pertaining to the study will be treated in a confidential manner.

________________________________________
Site Name

________________________________________
Print Name

________________________________________ ________________
Signature Date
