## Supplementary material for "Phase I study of a SARS-CoV-2 mRNA vaccine PTX-COVID19-B": Statistical Analysis Plan

### A PHASE I, FIRST-IN-HUMAN, OBSERVER-BLINDED, RANDOMIZED, PLACEBO-CONTROLLED, ASCENDING DOSE STUDY TO EVALUATE THE SAFETY, TOLERABILITY, AND IMMUNOGENICITY OF PTX-COVID19-B VACCINE IN HEALTHY SERONEGATIVE ADULTS AGED 18-64

SAP Version 1.0
Final
Date: 19 MAR 2021

for

Protocol No. PRO-CL-001

Submitted to:
Providence Therapeutics Holdings Inc.
661 University Avenue, Suite 1300
Toronto, ON M5G 0B7 Canada

Prepared by:
ICON Clinical Research, LLC
820 West Diamond Avenue, Suite 100
Gaithersburg, MD 20878

DocUUID : 37b099b4-2512-4dbf-8238-d258ccb127ba

### SIGNATURES

#### ICON Early Phase Services, LLC Signature Page

**ICON Author:**

Shuchi Joshi
03 Apr 2021 00:31:003+0000
REASON: I approve this document as author.
e0ddc4f6-173f-436a-9e21-5abf1441f0b8

Shuchi Joshi Date
Senior Statistician II, Biostatistics

**ICON Reviewers:**

Geethalakshmi Gunasekaran
02 Apr 2021 02:57:035+0000
REASON: I approve this document
db176dec-b675-4a8e-8604-b1ce75c558d1

Geethalakshmi Gunasekaran, Date
Principal Biostatistician, Biostatistics

Amy Craven
05 Apr 2021 14:08:009+0000
REASON: I approve this document
89c2758b-e52d-44a7-9569-a280f526fd7d

Amy Craven Date
Senior SAS Programmer II, IEP

Uma Arumugam
09 Apr 2021 12:56:003+0000
REASON: I approve this document
d76df923-b741-4eb3-8c04-561fecf38f81

Uma Arumugam Date
Medical Director, Clinical R&D, IEP

DocUUID : 37b099b4-2512-4dbf-8238-d258ccb127ba

### CLIENT SIGNATURE PAGE

#### Client Approvals:

Piyush Patel Piyush Patel
09 Apr 2021 12:44:053+0000
REASON: I approve this document
a8eccfa8-76aa-4ec8-b1ad-7650750ed318

Dr. Piyush Patel Date
Chief Medical Officer
Providence Therapeutics

DocUUID : 37b099b4-2512-4dbf-8238-d258ccb127ba

### LIST OF ABBREVIATIONS

| Abbreviation | Definition |
|---|---|
| AE | Adverse Event |
| AESI | AE of special interest |
| ALT | Alanine aminotransferase |
| ALP | Alkaline Phosphatase |
| AST | Aspartate aminotransferase |
| ATC | Anatomical Therapeutic Chemical |
| BMI | Body Mass Index |
| CI | Confidence Interval |
| COVID-19 | Coronavirus Disease 2019 |
| CSR | Clinical Study Report |
| CV | Coefficient of Variation |
| DILI | Drug-Induced Liver Injury |
| eCRF | Electronic Case Report Form |
| ELISA | Enzyme-Linked Immunosorbent Assay |
| HBsAg | Hepatitis B Surface Antigen |
| HCV | Hepatitis C Virus |
| HIV | Human Immunodeficiency Virus |
| ICD | International Classification of Disease |
| ICH | International Council for Harmonisation |
| Ig | Immunoglobulin |
| IM | Intramuscular |
| iSRC | Independent Safety Review Committee |
| LS | Least Square |
| MAAEs | Medically attended adverse events |
| MedDRA | Medical Dictionary for Regulatory Activities |
| mITT | Modified Intent-to-Treat |
| mRNA | messenger Ribonucleic acid |
| N | Number |
| NOCD | New Onset Chronic Disease |
| NP | Nasopharyngeal |
| PBMC | Peripheral Blood Mononuclear Cell |
| PIMMC | Potential Immune-Mediated Medical Condition |
| PN | Preferred Name |
| PP | Per Protocol |
| PT | Preferred Term |
| RSV | Respiratory Syncytial Virus |
| RT-PCR | Reverse Transcription-Polymerase Chain Reaction |
| SAE | Serious Adverse Event |
| SAP | Statistical Analysis Plan |
| SARS-CoV-2 | Severe Acute Respiratory Syndrome coronavirus 2 |
| SD | Standard Deviation |

DocUUID : 37b099b4-2512-4dbf-8238-d258ccb127ba

| | |
|---|---|
| SOC | System Organ Class |
| SRM | Study Reference Manual |
| SSP | Study-Specific Procedure |
| TEAE | Treatment-Emergent Adverse Event |
| ULN | Upper Limit of Normal |
| WHO | World Health Organization |

DocUUID : 37b099b4-2512-4dbf-8238-d258ccb127ba

DocUUID : 37b099b4-2512-4dbf-8238-d258ccb127ba

### 1. INTRODUCTION

This statistical analysis plan (SAP) is consistent with the statistical methods section of the study protocol (Version 5.0, dated 14 February 2021) and includes additional detail of the safety, tolerability and immunogenicity summaries to be included in the clinical study report (CSR).

The analysis plan may change due to unforeseen circumstances. Any changes post SAP finalization will be documented in the CSR.

DocUUID : 37b099b4-2512-4dbf-8238-d258ccb127ba

### 2. STUDY OBJECTIVES AND ENDPOINTS

#### 2.1 Objective

The study objectives are as follows:

- To evaluate the safety and tolerability of 2 doses of PTX-COVID19-B vaccine in healthy seronegative adults 18-64 years of age
- To evaluate the immunogenicity of 2 doses of PTX-COVID19-B vaccine in healthy seronegative adults 18-64 years of age

#### 2.2 Endpoint

The following are the study endpoint.

Safety and tolerability:

- Occurrence of events during the follow-up after each vaccination will be analyzed using both the Per Protocol (PP) and Safety Populations:
    - Vital signs and administration site reactions (eg, arm check evaluations including pain, tenderness, erythema/redness, induration/swelling) during the follow-up after each vaccination
    - Daily solicited adverse events (eg, fever, chills, nausea/vomiting, diarrhea, headache, fatigue, myalgia) through the second day post each vaccination.
- Overall safety will be analyzed using both the Safety and PP Populations.
    - Unsolicited adverse events from Day 1 through Day 42
    - Medically attended adverse events (AE) (Day 1 through Day 42), new onset chronic disease (NOCD), serious adverse events (SAE), adverse events of special interest (AESI) and potential immune-mediated medical condition (PIMMC) from Day 1 through to Day 395 (approximately 1 year after the last vaccination)
    - Findings from targeted physical examinations, vital sign assessments, and clinical safety laboratory testing

Immune Response:

- Immunogenicity analysis using the Safety, PP and mITT Populations. Analysis using the mITT Population will not be performed if it differs from the PP Population by ≤ 5% of the subjects for each of the treatment groups:
    - Antibodies (anti-COVID-19 immunoglobulin [Ig], IgG, IgA [ELISA]; and anti-COVID-19 neutralization titer assays)

DocUUID : 37b099b4-2512-4dbf-8238-d258ccb127ba

- Cell-mediated immunity using blood/ peripheral blood mononuclear cell (PBMC) (Flow Cytometry, Enzyme Linked Immunospot Assay)

DocUUID : 37b099b4-2512-4dbf-8238-d258ccb127ba

DocUUID : 37b099b4-2512-4dbf-8238-d258ccb127ba

- ~14 days between Cohort 2 (at least 7 days of AE and safety data) and Cohort 3 Day 1 for iSRC review

- Cohort 3: 20 healthy subjects 18 to 64 years of age administered 100 µg PTX-COVID19-B IM vaccine or placebo; 5 sentinel subjects (4 PTX-COVID19-B:1 placebo), followed by remaining 11 PTX-COVID19-B:4 placebo (cohort expansion)

Occurrence and confirmation of study halting rules will result in suspension of further enrollment and vaccine administration pending urgent review (within ~1 week) of the safety data by the iSRC (Section 6.7.2 of the protocol).

DocUUID : 37b099b4-2512-4dbf-8238-d258ccb127ba

Figure 3-1: Study Schematic

| **Phase 1:** | | | | |
|---|---|---|---|---|
| Subjects 18 to 64 years of age | | | | |
| Cohort 1<br>5 Sentinel Subjects, Safety Review ➡ Remaining 15 Subjects | ~14 Days, ➡ iSRC Meets | Cohort 2<br>5 Sentinel Subjects, Safety Review ➡ Remaining 15 Subjects | ~14 Days, ➡ iSRC Meets | Cohort 3<br>5 Sentinel Subjects, Safety Review ➡ Remaining 15 Subjects |

DocUUID : 37b099b4-2512-4dbf-8238-d258ccb127ba

#### Table 3-1: Schedule of Activities

| | Screening | Treatment Period | | | | | | | | End of study [a] |
|---|---|---|---|---|---|---|---|---|---|---|
| **Study Procedure** **Day** | **-21 to -1** | **1 [b]** | **2** | **8** | **28** | **29** | **42** | **90** | **180** | **395** |
| **Window allowance (Days)** | **-** | | | | **± 2** | | **± 2** | **± 3** | **± 5** | **-14** |
| Clinic visit | X | X | | X | X | | X | X | X | X |
| Telephone call (safety check) [c] | | | X | | | X | | | | |
| Informed consent | X | | | | | | | | | |
| Inclusion/exclusion criteria | X | X | | | | | | | | |
| Demographics | X | | | | | | | | | |
| Height, body weight, BMI | X [d] | | | | | | | | | |
| Medical and surgical history | X | | | | | | | | | |
| Complete physical examination | X | | | | | | | | | |
| Symptom-directed, targeted physical examination | | X | | X | X | | X | X | X | X |
| Vital signs [e] | X | X [f] | | X | X [f] | | X | X | X | X |
| Clinical laboratory tests [g] | X | | | X | X | | X | X | | |
| Serology (HIV-1 and -2, HBsAg, HCV) | X | | | | | | | | | |
| Pregnancy test (women of child-bearing potential) | X | X | | | X | | | | | |
| Urine drugs of abuse | X | | | | | | | | | |
| NP swab for COVID-19 via RT-PCR [h] | X | | | | X | | | | | |
| Blood samples for immunogenicity analysis (antibodies IgM, IgG, IgA, neutralization) | X | | | X | X | | X | X | X | X |
| Blood/PBMC samples for cell-mediated immunity (pre-dose as applicable) | | X | | X | X | | X | X | X | X |
| Randomization | | X [i] | | | | | | | | |
| Training of subjects on diary completion | | X | | | X | | | | | |

DocUUID : 37b099b4-2512-4dbf-8238-d258ccb127ba

| | Screening | Treatment Period | | | | | | | | End of study [a] |
|---|---|---|---|---|---|---|---|---|---|---|
| **Study Procedure** **Day** | **-21 to -1** | **1 [b]** | **2** | **8** | **28** | **29** | **42** | **90** | **180** | **395** |
| Distribute paper diary | | X | | | X | | | | | |
| Review paper diary | | | | X | | | X | | | |
| Predose, confirm eligibility for vaccination | | X | | | X | | | | | |
| Vaccine administration [j] | | X | | | X | | | | | |
| Post-vaccination assessment (approximately 1 hour post vaccination, includes arm check and vital signs) | | X | | | X | | | | | |
| Medically-attended AE assessments [k] | | X | | X | X | | X | | | |
| Unsolicited AE assessments | | ← | ← | ← | ← | ← | → | | | |
| Solicited AEs [l] | | X | X | | X | X | | | | |
| Assessments of NOCD, AESIs (including COVID-19 cases for enhanced disease), and PIMMCs [h] | | X | | X | X | | X | X | X | X |
| Serious adverse event assessments | ← | ← | ← | ← | ← | ← | ← | ← | ← | → |
| Prior and concomitant medications/procedures/other vaccinations [m] | ← | ← | ← | ← | ← | ← | ← | ← | ← | → |

Abbreviations: AEs = adverse events; AESI = adverse event of special interest; BMI = body mass index; COVID-19 = coronavirus disease 2019; HBsAg = hepatitis B surface antigen; HCV = hepatitis C virus; HIV = human immunodeficiency virus; Ig = immunoglobulin; IM = intramuscular; NOCD = new onset chronic disease; NP = nasopharyngeal; PBMC = peripheral blood mononuclear cell; PIMMC = potential immune-mediated medical condition; RT-PCR = reverse transcription-polymerase chain reaction

a. Subjects who withdraw from the study early will have end-of-study procedures performed at the time of discontinuation.

b. Study entry assessments conducted on Day 1 will be used to reconfirm a subject's eligibility for enrollment into the study.

c. All subjects will have safety calls approximately 24 hours after each of the vaccinations. Trained site staff will inquire about the presence of solicited and unsolicited AEs and remind subjects to keep their diary cards up to date.

d. BMI will be calculated using the height obtained at screening.

e. Vital signs will be measured after the subject has been resting quietly in a seated position for at least 5 minutes. Vital signs include systolic and diastolic blood pressures, heart rate, respiratory rate, and oral temperature (between 35ºC and37.5ºC)-. Pulse oximetry will be performed prior to each vaccination (Pulse oximetry must be >90%). Unscheduled visits for COVID-like illness will require pulse oximetry in addition to standard vital signs. Temperature at all visits is to be between 35°C and 37.5°C

DocUUID : 37b099b4-2512-4dbf-8238-d258ccb127ba

f. Vital signs on Days 1 and 28 will be collected before vaccination and approximately 1 hour after vaccination (before subjects are discharged from the clinic).

DocUUID : 37b099b4-2512-4dbf-8238-d258ccb127ba

#### 3.2 Study Population

A total of 60 healthy adult subjects with BMI of ≥ 18 and ≤ 30 kg/m$^2$ at screening will be enrolled.

#### 3.3 Evaluations at Screening and Check-in

Screening will occur between Days -21 to -1 from signing consent. For further details on these procedures of schedule of activities refer to Table 3-1.

#### 3.4 Randomization and Treatment Assignments

On Day 1, randomization will be performed. Three cohorts of 20 subjects each will include sentinel groups of 5 subjects randomized in 4:1 ratio for investigational vaccine (PTX-COVID19-B): placebo, followed by a cohort expansion phase of 15 subjects randomized in 11:4 ratio for investigational vaccine (PTX-COVID19-B): placebo.

The randomization list will be provided as a paper listing of vaccine or placebo assignment prepared by ICON's unblinded Biostatistician.

ICON's unblinded statistician will provide the randomization list to the site's unblinded pharmacists. If the placebo and vaccine are not identical in appearance, the staff member administering the doses will be unblinded and will not participate in any subject assessments. In order to preserve the blind, vaccine will be prepared and administered in a manner that masks the content for both the subject and any observers.

If an SAE or other circumstance require that the blind be broken to ensure subject safety, the investigator must immediately notify the medical monitor and/or sponsor. ICON/designee will notify the sponsor as soon as possible and will release the treatment assignment of the subject. If the treatment assignment is unblinded by the site pharmacist or any party other than ICON, the investigator must notify ICON in writing and document the course of events in the source records. Any subject for whom the treatment code is prematurely released will be withdrawn from the study.

#### 3.5 Determination of Sample Size

No formal sample size calculation was performed. A total of 60 adult subjects will be enrolled in the study.

Rare AEs are not demonstrable in a clinical study of this size; however, the probabilities of observing one or more AEs given various true event rates are presented in Table 3-2. With the assumption that all enrolled subjects will likely complete immunizations and safety visits in this relatively short duration study, the following statistical considerations apply. With 15 subjects in each dose group, the chance of observing at least 1 AE of probability ≥ 20% is approximately 97%. Therefore, if no AEs of a given type occur in a dose cohort, it is relatively certain that they will occur in < 20% of people once the vaccine is implemented. With 45 subjects across the 3

DocUUID : 37b099b4-2512-4dbf-8238-d258ccb127ba

Phase 1 dosing cohorts, the chance of observing at least 1 AE of probability ≥ 5% is at least 90%. Therefore, if no AEs of a given type occur across the combined doses, it is relatively certain that any dosage-independent event will occur in < 5% of people once the vaccine is implemented.

**Table 3-2 Probability of Observing an Adverse Event for Various Event Rates**

| N | "True" Event Rate | Probability of Observation (%) | N | "True" Event Rate | Probability of Observation (%) |
|---|---|---|---|---|---|
| 15 | 0.1% | 1.5 | 45 | 0.1% | 4.4 |
| | 0.5% | 7.2 | | 0.5% | 20.2 |
| | 1.0% | 14.0 | | 1.0% | 36.4 |
| | 2.0% | 26.1 | | 2.0% | 59.7 |
| | 3.0% | 36.7 | | 3.0% | 74.6 |
| | 4.0% | 45.8 | | 4.0% | 84.1 |
| | 5.0% | 53.7 | | 5.0% | 90.1 |
| | 10.0% | 79.4 | | 10.0% | 99.1 |
| | 15.0% | 91.3 | | 15.0% | 99.9 |
| | 20.0% | 96.5 | | 20.0% | >99.9 |

#### 3.6 Study Drug Administration

The vaccine product, PTX-COVID19-B intramuscular (messenger ribonucleic acid [mRNA]) Humoral Vaccine, is a sterile formulation of mRNA (drug substance) in aqueous buffer that is administered by intramuscular (IM) injection in the upper arm deltoid muscle of the non-dominant side. Physiochemically, the product is a dispersion of nucleic acid/lipid nanoparticles with an average size of approximately 70 to 90 nm.

A description of the vaccine and control (placebo) is presented in Table 3-3.

**Table 3-3 Vaccine and Placebo**

| Vaccine | Dosage Form | Dose | Manufacturer |
|---|---|---|---|
| PTX-COVID19-B mRNA Humoral Vaccine | Injectable solution, IM vaccine | 0.2 mg/mL 0.5 mL, at dose levels presented in Section 3.1 | Genevant Sciences Corporation |
| Control (Placebo) Vaccine (0.9% Normal Saline) | Injectable solution, IM vaccine | 0.0 mg/mL 0.5 mL | Commercially available source |

Abbreviations: IM = intramuscular; mRNA = messenger ribonucleic acid

The Phase I study drug will be administrated in 3 dose levels (16, 40, and 100 µg). Two injections of the vaccine will be administered at an interval of 4 weeks.

DocUUID : 37b099b4-2512-4dbf-8238-d258ccb127ba

#### 3.7 Prior/Concomitant Medications and Procedures

Any medication taken by a subject from 30 days prior to the screening visit through Day 42 of the study and the reason for its use will be documented on concomitant medication electronic Case Report Form (eCRF). After Day 42, concomitant medications will be documented if associated with an SAE, AESI, medically-attended AE, NOCD, or PIMMC.

Prior medications are defined as those medications that started and stopped prior to the first dose of study treatment. Concomitant medications are defined as those medications with a start date on or after the first dose of study treatment or started prior to the first dose of study treatment and were continued after the first dose of study treatment.

Information recorded will include start and stop dates and times of medication, dose (unit), frequency, route of administration, and indication.

Concomitant procedures along with start and stop date, type and reason for procedures will be captured on eCRF.

#### 3.8 Drug Administration and Compliance

Each subject will receive 2 doses of vaccine (or placebo), the first on Day 1 and the second on Day 28. The total duration of subject participation is approximately 14 months (including screening, treatment, and follow-up periods).

All vaccine doses will be prepared and administered by delegated and trained staff at the clinical site. Details regarding dose preparation and dosing, including the dose administered and the date and time of dosing, will be recorded.

#### 3.9 Evaluation of Immunogenicity and Sampling Schedule

Blood samples or immunogenicity analysis will be collected as per Table 3-1. Immunogenicity assessments will include the following:

- Antibodies:
  - Anti-COVID-19 Ig, IgG, IgA by ELISA
  - Anti-COVID-19 neutralizing antibody titer assays

- Cell-mediated Immunity/PBMC Evaluations (Flow Cytometry)

- Additional assays may be conducted for research purposes.

The actual date and time of the blood sample collections for antibody (anti-COVID-19 Ig, IgG and IgA by ELISA) and Cell-mediated Immunity/PBMC Evaluations will be recorded in the subject's CRF.

Immunogenicity sample collection, processing, and shipping details will be outlined in a separate study reference manual (SRM).

DocUUID : 37b099b4-2512-4dbf-8238-d258ccb127ba

#### 3.10 Evaluation of Treatment Safety

##### 3.10.1 Adverse Events

An AE is defined as any untoward medical occurrence in a subject administered a pharmaceutical product during the course of a clinical investigation. An AE can therefore be any unfavorable and unintended sign, symptom, or disease temporally associated with the use of a vaccine, whether or not thought to be related to the vaccine.

A treatment-emergent AE (TEAE) is defined as any AE that began or worsened following the start of vaccination administration.

SAEs is defined as any event that results in death, is immediately life threatening, requires inpatient hospitalization or prolongation of existing hospitalization, results in persistent or significant disability/incapacity, results in a congenital anomaly or birth defect or important medical event.

Medically-attended AEs (MAAE) (Day 1 through Day 42) are defined as hospitalization, an emergency room visit, or an otherwise unscheduled visit to or from medical personnel for any reason except for a health maintenance visit for a routine physical.

New onset chronic diseases (NOCD) are defined as any new ICD diagnosis (per current WHO International Statistical Classification of Diseases and Related Health Problems) that is applied to the subject during the course of the study, after receipt of the study agent, that is expected to continue for at least 3 months and requires continued health care intervention.

An AE of Special Interest (AESI) is any AE that a regulatory authority has mandated be reported on an expedited basis, regardless of the seriousness, expectedness, or relatedness of the AE to the administration of investigational product. To date, no AESIs have been identified specifically for this vaccine. However, subjects who test positive for SARS-CoV-2, present with autoimmune disease, or are diagnosed with a PIMMC should be reported to ICON Pharmacovigilance (Section 9.4 of protocol) as an AESI or Suspected Unexpected Serious Adverse Reaction. The AE will be followed appropriately, including illness duration, severity, and outcome.

Potentially Immune-Mediated Medical Conditions (PIMMC) will be defined as any conditions occurring from Day 1 through to Day 395 (approximately 1 year after the last vaccination) due to the theoretical potential for induction of autoimmune diseases. For this study PIMMC will be considered as SAE and will be reported to medical monitor and/or sponsor immediately.

###### 3.10.1.1 Reactogenicity Assessments

Reactogenicity solicited events include, generalized symptoms such as headache, fatigue, muscle pain, joint pain, nausea, chills, and fever, and localized reactions including injection site pain, bruising, redness, and swelling. Subjects will be asked to record oral temperature with their own thermometer if they feel any of the above symptoms and record in the Diary Card.

DocUUID : 37b099b4-2512-4dbf-8238-d258ccb127ba

###### 3.10.1.2 Drug-induced Liver Injury (DILI)

Subjects will be monitored for signs of DILI. Potential events of DILI will be defined as meeting all of the following criteria (as per Health Canada Guidance Document: Pre-market Evaluation of Hepatotoxicity in Health Products, 2012):

- ALT or AST > 3 × Upper limit of normal (ULN)
- Total bilirubin > 2 × ULN without initial findings of cholestasis (elevated serum ALP)
- No other reason can be found to explain the combination of laboratory value increases

##### 3.10.2 Clinical Laboratory Assessments

Laboratory evaluations will be collected for hematology, chemistry and urinalysis parameters. Refer to schedule of events Table 3-1.

Subjects will refrain from strenuous exercise for 48 hours prior to laboratory sample collection and from alcohol for 72 hours prior to laboratory sample collection. Subjects will be non-fasted at screening; if glucose test result is outside the normal range then one fasted repeat test may be performed.

During screening, if a subject has an out-of-range value for a clinical safety laboratory parameter that the investigator believes is not clinically significant or the investigator does not believe is correct (eg, lab or specimen processing error), but the investigator wants to confirm with a repeat laboratory test, a single repeat is allowed to confirm the initial result.

Additional safety laboratory tests may be conducted as needed by the investigator to evaluate subject safety are presented below:

DocUUID : 37b099b4-2512-4dbf-8238-d258ccb127ba

**Table 3-4 Clinical Laboratory Tests**

DocUUID : 37b099b4-2512-4dbf-8238-d258ccb127ba

##### 3.10.5 Medical and Surgical History

Medical and surgical history will be collected at screening. As applicable, self-reported medical and surgical history will be collected at clinical site visits to determine if any changes have occurred since screening. Any ongoing chronic conditions must be stable with medication changes and no exacerbations in past 3 months and no hospitalizations in past 6 months.

##### 3.10.6 Other Safety Assessments

The following tests will be performed:

- Urine drugs of abuse (at a minimum, opiates, cocaine, phencyclidine, amphetamines, benzodiazepines, and methadone) screens
  - During the screening period, urine drugs of abuse screens may not be repeated for eligibility
  - An alcohol breathalyzer test will be performed at screening
- Serology tests (ie, HIV-1 and HIV-2 antibodies, HBsAg, and HCV antibody, and any confirmatory tests performed at the discretion of the investigator)
- Urine pregnancy test (females only)
- Immunogenicity analysis and cell mediated immunity
- Nasopharyngeal swab for COVID-19 RT-PCR (the clinic also has the option to confirm eligibility via an additional point-of-care test)
- Urine Alcohol Breath Test

#### 3.11 Protocol Deviation Reporting

A protocol deviation is defined as any intentional or unintentional change to, or noncompliance with, the approved protocol procedures or requirements. Deviations may result from the action or inaction of the subject, investigator, or site staff.

At the outset of the study, a process for defining and handling protocol deviations will be established. This will include determining which violations will be designated "key", requiring immediate notification to the medical monitor and/or sponsor. The investigator is responsible for seeing that any known protocol deviations are recorded and handled as agreed.

DocUUID : 37b099b4-2512-4dbf-8238-d258ccb127ba

### 4. CHANGES IN THE CONDUCT OF THE STUDY OR PLANNED ANALYSIS

All analyses specified in this SAP are consistent with the final study protocol (Version 5.0, dated 14FEB2021). Any changes in this analysis provided or any additional analysis performed will be documented in CSR.

DocUUID : 37b099b4-2512-4dbf-8238-d258ccb127ba

### 5. QUALITY CONTROL AND QUALITY ASSURANCE METHODS FOR DATA ANALYSIS

Case report forms will be monitored and collected by ICON. All monitored CRFs will be sent to the Data Management group at ICON and processed according to the ICON Study Specific Procedure SSP DM-54890001.01 Data Management Plan (DMP). The DMP describes CRF data processing, edit checks, data query management, medical dictionary coding, SAE reconciliation, data transfers, and data quality review through database lock or any necessary reopening of the database. After database lock, the data will be retrieved from the database using SAS® 9.4 version (or higher).

DocUUID : 37b099b4-2512-4dbf-8238-d258ccb127ba

### 6. STATISTICAL METHODS

#### 6.1 General

The statistical analysis will be conducted following the principles specified in the International Council for Harmonization (ICH) Topic E9 Statistical Principles for Clinical Trials (CPMP/ICH/363/96).

All statistical analyses will be performed using the statistical software SAS GRID Linux/SAS Studio (version 9.4 or higher) and any exceptions will be detailed in the CSR.

All results collected in the database will be presented in listings. Both observed values and change-from-baseline values for each subject will be given where applicable. All continuous data will be listed with the same precision as presented in the database. Subject ID, Cohort, Treatment and study day, will sort data listings.

Unless otherwise noted, continuous immunogenicity variables will be summarized using number of non-missing observations (n), arithmetic mean (mean), standard deviation (SD), median, minimum, maximum and 95% confidence interval (CI). All other continuous variables will be summarized using n, mean, SD, median, minimum and maximum. For categorical variables, the number and percentage (as the percentage of subjects in each category relative to the total number of subjects in each category) will be the default summary presentation. No formal significance testing will be performed.

In the data listings, study day relative to dosing date may be presented. Study day relative to dose will be calculated as: event date – first dose date (+ 1 if event date ≥ first dose date).

Baseline is defined as the last observed value of the parameter of interest prior to first dosing of study vaccine (this includes unscheduled visits). For numerical variables, change from baseline will be calculated as (Post baseline value – Baseline value).

For safety, reactogenicity and immunogenicity summaries, the unscheduled and repeat assessments will not be summarized; however, all results will be included in the data listings.

Disposition, demographics and baseline characteristics and all other safety parameters will be summarized by Cohort and treatment (i.e. PTX-COVID19-B [16 µg], PTX-COVID19-B [40 µg], PTX-COVID19-B [100 µg], Placebo) and Overall.

Immunogenicity and cellular immunity parameters will be summarized by scheduled timepoints for each cohort and treatment.

The database will be locked and unblinded after the data have been monitored for all subjects at approximately 5 weeks after the last subject receives their last dose and then again after all subjects complete the study.

The final analysis of the safety and immunogenicity data collected up to approximately 1 year after the last vaccination will be performed after the study is completed. An addendum to the initial CSR will be prepared with data from final analysis.

DocUUID : 37b099b4-2512-4dbf-8238-d258ccb127ba

#### 6.2 Handling of Dropouts or Missing Data

Missing data will be considered as missing at random and no data imputation will be performed.

##### 6.2.1 Handling of missing/ incomplete dates for Adverse Event

Imputation rules for missing or partial AE start date are defined below:

**If only Day of AE start date is missing:**

If the start date has month and year but day is missing, the first day of the month will be imputed

- If this date is earlier than the first dose date, then the first dose date will be used instead.
- If this date is later than the stop date (possibly imputed), then the stop date will be used instead.

**If Day and Month of AE start date are missing:**

If the start date has year, but day and month are missing, the 1st of January will be imputed

- If this date is earlier than the first dose date, then the first dose date will be used instead.
- If this date is later than the stop date (possibly imputed), then the stop date will be used instead.

**If Year of AE start date is missing:**

If the year of AE start is missing or AE start date is completely missing then imputation will not be done.

**Missing or partial AE stop date:**

- If only Day is missing, the last day of the month will be assumed.
- If Day and Month are both missing, the last day of the year will be assumed.
- If Day, Month and Year are all missing, ‘Ongoing’ status to stop date will be assigned.

If the AE stop date (full or partial) is before the first dose date then the AE should be considered as a pre-treatment AE. Otherwise, the AE will be considered as TEAE.

##### 6.2.2 Handling of missing or partial Prior/Concomitant Medication Dates

Missing or partial medication start date:

a. If only Day is missing, the first day of the month will be assumed.
b. If Day and Month are both missing, the first day of the year will be assumed.
c. If Day, Month and Year are all missing, the day before the first dose date will be assumed.

Missing or partial medication stop date:

DocUUID : 37b099b4-2512-4dbf-8238-d258ccb127ba

a. If only Day is missing, the last day of the month will be assumed.
b. If Day and Month are both missing, the last day of the year will be assumed.
c. If Day, Month and year are all missing, 'Ongoing' status to stop date will be assigned.

#### 6.3 Multiple Comparisons and Multiplicity

There is no multiple comparisons/endpoints.

#### 6.4 Adjustments for covariates

There is no adjustment on covariates in this study.

#### 6.5 Multicenter Studies

This is a single-center study.

#### 6.6 Examination of Subgroups

No subgroup analyses are planned.

#### 6.7 Coding dictionaries

Medical history and AEs will be coded using the Medical Dictionary for Regulatory Activities (MedDRA) Version 23.1 or higher. Medications will be coded using the World Health Organization Drug Dictionary (WHO Global B3 format - SEP 2020 or higher. Medical procedures will not be coded.

#### 6.8 Analysis Sets

##### 6.8.1 Randomized Population

The Randomized Set will consist of subjects who are randomization in to the study. In this population, study drug will be assigned based on the study drug to which subjects were randomized, regardless of which study drug they actually received. This population will be used for disposition summary.

##### 6.8.2 Safety Population

All subjects who provide consent, are randomized, and receive any amount of vaccine/placebo.

The Safety Population will be used for demographic, baseline characteristic, safety analyses will be analyzed as actually treated.

DocUUID : 37b099b4-2512-4dbf-8238-d258ccb127ba

##### 6.8.3 Per Protocol Population

Per protocol (PP) population will consist of all subjects in the Safety Population who receive the assigned doses of the vaccine/placebo according to protocol, have serology results, and have no major protocol deviations affecting the primary immunogenicity outcomes, as determined by the Sponsor before database lock and unblinding.

##### 6.8.4 Modified Intent-to-Treat Population

The mITT population will includes all subjects in the Safety Population who provide any serology data.

The mITT Population will be used for the analysis of immunogenicity endpoints. Analysis using the mITT Population will not be performed if it differs from the PP Population by ≤ 5% of the subjects for each of the treatment groups.

#### 6.9 Subject Accountability

Summaries of analysis populations and subject disposition will be presented by cohort, treatment and overall, will contain the following information:

- Number of subjects in randomized population
- Number and percent of subjects dosed
- Number and percent of subjects who received both doses
- Number and percent of subjects who completed the study
- Number and percent of subjects who discontinued early and reason for early termination
- Number and percent of subjects in the safety, PP and mITT Population.

This summary will be based on randomized population.

Subject disposition and Populations, eligibility criteria satisfaction and consent information will be presented in listings for randomized population.

#### 6.10 Protocol Deviations

All protocol deviations will be listed for safety population.

#### 6.11 Subject Demographics Characteristics

Subject demographics characteristics will be listed and summarized descriptively for all subjects by cohort, treatment and overall. The summary will include the subjects' age at informed consent (in years), gender, race, ethnicity, weight (in kg), height (in cm) and BMI (in kg/m$^2$) using safety population.

#### 6.12 Medical and Surgical History

The medical and surgical history data will be listed for safety population.

DocUUID : 37b099b4-2512-4dbf-8238-d258ccb127ba

#### 6.13 Measurements of Treatment Compliance

Individual subject listing will be provided for eCRF collected exposure data for safety population. In addition, eligibility for vaccination (presenting body temperature) will also be reported in a listing.

#### 6.14 Immunogenicity Analysis

The Safety population will be used for all listings. Summary statistics and analysis corresponding to immunogenicity analysis will be performed for Safety mITT and PP populations. Analysis using the mITT Population will not be performed if it differs from the PP Population by ≤5% of the subjects for each of the treatment groups.

The individual sampling and blood collection for anti-COVID-19 RBD Ig, IgG, IgA and anti-COVID-19 neutralization titer will be listed and summarized by cohort, treatment, and time point using descriptive statistics (n, mean, SD, CV [%], median, minimum, median, maximum geometric mean [GM] and 95% CI).

The individual sampling and subject titer value will be displayed graphically on linear and semi-log scales. Linear and semi-logarithmic plots of the individual titer values by actual sampling time for each cohort will be provided by subject (one subject per page). Plots of mean (SD) titer profiles versus time on linear and semi-log scales will be presented by all cohorts being superimposed on the same plot.

Each natural log-transferred (ln) post first dose assessments of Ig or anti COVID-19 neutralization titer assays will be analyzed using a mixed effects model with treatment as fixed effects and subject as a random effect for each visit. Placebo subjects will be pooled for comparison. Analysis will be performed at 5% level of significance. The geometric Least Square Mean (LSM) ratio of treatment PTX-COVID19-B (Test) vs pooled placebo (Reference) and the corresponding 95% CIs for the ratio will be calculated and presented after back-transformation by exponentiation. Individual treatment geometric LSM will also be presented.

The following example code can be used for statistical analysis. Please note that required modification should be made as necessary as per the data

```
PROC MIXED DATA=DATASET;
BY Visit;
        CLASS Subject Treatment;
        MODEL lnValue = Treatment/ DDFM=KR;
        RANDOM Subject;
        LSMEANS Treatment;
        ESTIMATE ‘PTX-COVID19-B 16 ug vs Placebo’ Treatment -1 1 0 0 / ALPHA=0.05
CL;
        ESTIMATE ‘PTX-COVID19-B 40 ug vs Placebo’ Treatment -1 0 1 0 / ALPHA=0.05
CL;
        ESTIMATE ‘PTX-COVID19-B 100 ug vs Placebo’ Treatment -1 0 0 1 / ALPHA=0.05
CL;
```

DocUUID : 37b099b4-2512-4dbf-8238-d258ccb127ba

RUN;

#### 6.15 Cell-mediated Immunity Analysis

The Safety population will be used for all listings. Summary statistics and corresponding analysis will be performed for Safety, mITT and PP populations. Analysis using the mITT Population will not be performed if it differs from the PP Population by ≤5% of the subjects for each of the treatment groups.

Cell-mediated Immunity/PBMC Evaluations (Flow Cytometry Assay) will be listed and summarized by cohort, treatment and time point using descriptive statistics (n, mean, SD, CV [%], median, minimum, median, maximum, GM and 95% CI).

The individual sampling and subject titer value will be displayed graphically on linear and semi-log scales. Linear and semi-logarithmic plots of the individual titer values by actual sampling time for each cohort will be provided by subject (one subject per page). Plots of mean (SD) titer profiles versus time on linear and semi-log scales will be presented by all cohorts being superimposed on the same plot.

Each natural log-transformed post first dose assessments of value will be analyzed using a mixed effects model with terms and treatment as fixed effects and subject as a random effect for each visit. Placebo subjects will be pooled for comparison. Analysis will be performed at 5% level of significance. The geometric Least Square Mean (LSM) ratios of treatment PTX-COVID19-B (Test) vs placebo (Reference) and the corresponding 95% CIs for the ratios will be calculated and presented after back-transformation by exponentiation. Individual treatment geometric LSM will also be presented.

The following example code can be used for statistical analysis. Please note that required modification should be made as necessary as per the data

```
PROC MIXED DATA=DATASET;
BY Visit;
        CLASS Subject Treatment;
        MODEL lnValue = Treatment/ DDFM=KR;
        RANDOM Subject;
        LSMEANS Treatment;
        ESTIMATE 'PTX-COVID19-B 16 ug vs Placebo' Treatment -1 1 0 0 / ALPHA=0.05
CL;
        ESTIMATE 'PTX-COVID19-B 40 ug vs Placebo' Treatment -1 0 1 0 / ALPHA=0.05
CL;
        ESTIMATE 'PTX-COVID19-B 100 ug vs Placebo' Treatment -1 0 0 1 / ALPHA=0.05
CL;

RUN;
```

DocUUID : 37b099b4-2512-4dbf-8238-d258ccb127ba

#### 6.16 Safety Analyses

All Safety analysis will be analyzed for the Safety and PP populations, however, listings will be presented for Safety population only.

All safety data will be summarized by cohort, treatment (as applicable), and time point. No statistical tests will be performed.

##### 6.16.1 Adverse Events

All AE summaries will include treatment emergent AEs. Treatment-emergent AEs are those which occur or worsen after the start of study vaccination, Day 1 to Day 42.

A causally related AE is defined as any AE that is assessed by the Investigator as relationship to treatment.

All AEs will be coded by primary system organ class (SOC) and preferred term (PT) according to the Medical Dictionary for Regulatory Activities (MedDRA) Version 23.1 or higher and presented by subject in data listings.

The overall incidence of TEAEs (number and percentage of subjects) and event (number of events) will be summarized by cohort, treatment and overall. It includes, TEAEs, MAAEs and AESIs up to Day 42; severity of TEAEs, MAAEs and AESIs up to Day 42; relation to treatment of TEAEs, MAAEs and AESIs up to Day 42; TEAEs leading to study discontinuation; SAEs, MAAEs, NOCDs and PIMMCs through Day 395; life-threatening SAE and SAEs resulting in death.

The TEAEs will be summarized and tabulated at both the subject (n [%] of subjects) and event (number of events) level:

- TEAEs by SOC and PT
- MAAEs by SOC and PT
- NOCDs by SOC and PT
- PIMMCs by SOC and PT
- TEAEs by SOC, PT and severity
- MAAEs by SOC, PT and severity
- AESIs by SOC, PT and severity
- TEAEs by SOC, PT and relationship
- MAAEs by SOC, PT and relationship
- AESIs by SOC, PT and relationship
- SAEs by SOC and PT

For the incidence at the subject level by SOC and PT, if a subject experiences more than 1 event within the same SOC and PT, only 1 occurrence will be included in the incidence.

DocUUID : 37b099b4-2512-4dbf-8238-d258ccb127ba

For the incidence at the subject level by SOC, PT, and severity, if a subject experiences more than 1 event within the same SOC and PT, only the most severe occurrence will be included in the incidence.

For the incidence at the subject level by SOC, PT, and severity, if a subject experiences more than 1 event within the same SOC and PT, only the most severe occurrence will be included in the incidence.

Any SAEs, AEs with outcome of death, or AEs resulting in discontinuation of study or study drug will be listed separately.

###### 6.16.1.1 Reactogenicity

Vaccine Reaction and Reactogenicity will be collected through diary card and will be listed.

Following summaries will be provided by cohort, treatment and overall based on safety and PP populations:

- The number and percentage of subjects with at least one solicited systemic events and site reactions during 7- day follow-up after each vaccination by severity.

- The number and percentage of subjects with at least one solicited systemic events and site reactions during 7- day follow-up after each vaccination by day of immunization and severity.

Individual listing will be provided for solicited systemic events and site reactions at each collected reactogenicity days.

##### 6.16.2 Clinical Laboratory Assessments

Observed values and change from baseline for each parameter of continuous clinical laboratory values (hematology, chemistry and urinalysis) will be summarized by cohort, treatment and visit/time points using descriptive statistics.

Shift from baseline to post-baseline laboratory findings in normal range criteria will also be summarized.

A listing of all clinical laboratory data for each subject at each visit will be presented. Clinically significant laboratory values that are outside the normal ranges will be flagged and presented in a separate listing.

##### 6.16.3 Vital Signs

Observed values and change from baseline for each parameter of continuous vital sign parameters (blood pressure, oral temperature, heart rate, respiratory rate and oral temperature) summarized by cohort, treatment and visit/time points using descriptive statistics.

DocUUID : 37b099b4-2512-4dbf-8238-d258ccb127ba

Pulse oximetry will be performed prior to each vaccination and will be listed in listings.

All vital signs data will be listed individually by each subject. Incidence of clinically significant values will be flagged in the listing.

##### 6.16.4 Physical Examinations

Abnormal physical examination findings will be presented in subject listing.

##### 6.16.5 Prior and Concomitant Medications

Prior and Concomitant medications will be coded using the WHO Drug Dictionary (Version WHO Global B3 format - SEP 2020 or higher) and classified according to anatomical therapeutic chemical code (ATC) levels. All prior and concomitant medications data will be listed by ATC 2 classification and preferred name (PN).

A separate listing will be provided for concomitant procedures.

##### 6.16.6 Other Observations Related to Safety

The following additional other safety data information will be provided in a separate listings.

- Serology tests result (ie, HIV-1 and HIV-2 antibodies, HBsAg, and HCV antibody)
- Nasopharyngeal swab (RT-PCR) data
- Urine drug screen, alcohol breath, serology tests results will be listed.
- Blood and urine pregnancy testing in female subjects will be provided in subjects listing.

#### 6.17 Planned Interim Analysis

At 14 days post-second dose, there will be a review of all available data collected for safety and immunogenicity endpoints to determine the appropriate dose, plan, and study design for moving forward.

The iSRC will convene between all cohorts to review available safety data to ensure that there are no safety concerns and that none of the Study Halting Rules have been met before recommending that the study enroll the next cohort.

At approximately 14 days after subject receives their last dose, (Day 42) there will be an interim reviews by the study team and analysis of all data collected till Day 42 for each subject for safety and immunogenicity endpoints.

The available data will be cleaned, and the database will be locked. The interim analysis will be done by the study team and unblinded for the data till Day 42 for each subject. Further visits data will not be considered in the interim analysis. A CSR will be written for data from Day 1 through Day 42.

DocUUID : 37b099b4-2512-4dbf-8238-d258ccb127ba

When longer term follow-up (Day 57 to Day 395) has been completed for all subjects after the end of the study, further reviews and analysis will be performed on all data. An addendum to the clinical study report will also be written.

#### 6.18 General Conventions for Tables, Listings and Figures

Tables and listings will be presented in landscape mode with minimum of 3/4” bound edge margin and 3/8” other margins on 8.5” x 11” paper.

Times new roman font size of no less than 8 point will be used for tables and listings.

A source line will be included on the bottom of each page of all tables and listings. It will contain the SAS code program name and the run date and time.

Each variable is recorded to a specific number of decimal places. If the raw data is presented with varying precision, then the least precise value will be considered as the data precision.

For summary tables, unless otherwise specified, the number of decimal places provided in the tables and listings will be based on the accuracy of the least accurate value in the raw data as follows:

| | |
|---|---|
| n | integer |
| Arithmetic mean | 1 decimal place more than the least accurate number in the raw data |
| SD | 2 decimal place more than the least accurate number in the raw data |
| CV(%) | 2 decimal places |
| Geometric mean | 1 decimal place more than the least accurate number in the raw data |
| Median | 1 decimal place more than the least accurate number in the raw data |
| Minimum | same number of decimal places as raw data |
| Maximum | same number of decimal places as raw data |
| Confidence interval | same number of decimals as the associated statistic |
| Percentage | 1 decimal place |

DocUUID : 37b099b4-2512-4dbf-8238-d258ccb127ba

### 7. LIST OF TABLES, FIGURES, AND LISTINGS

| Table/Figure Number | Table/Figure Name | Interim Analysis |
|---|---|---|
| **Section 14.1** | **Demographic and Subject Characteristics Data Summaries** | |
| Table 14.1.1 | Summary of Subject Disposition (Randomized Population) | X |
| Table 14.1.2 | Analysis Populations | X |
| Table 14.1.3 | Demographic and Baseline Characteristics (Safety Population) | X |
| **Section 14.2** | **Immunogenicity Data Summaries** | |
| Table 14.2.1.1.1 | Statistical Analysis of Anti-COVID-19 RBD Immunoglobulin G by Time point (PP Population) | X |
| Table 14.2.1.1.2 | Statistical Analysis of Anti-COVID-19 RBD Immunoglobulin G by Time point (Safety Population) | X |
| Table 14.2.1.1.3 | Statistical Analysis of Anti-COVID-19 RBD Immunoglobulin G by Time point (mITT Population) | X |
| Table 14.2.1.2.1 | Statistical Analysis of Total Anti S-Protein Immunoglobulin G by Time point (PP Population) | X |
| Table 14.2.1.2.2 | Statistical Analysis of Total Anti S-Protein Immunoglobulin G by Time point (Safety Population) | X |
| Table 14.2.1.2.3 | Statistical Analysis of Total Anti S-Protein Immunoglobulin G by Time point (mITT Population) | X |
| Table 14.2.1.3.1 | Statistical Analysis of Immunoglobulin A by Time point (PP Population) | |
| Table 14.2.1.3.2 | Statistical Analysis of Immunoglobulin A by Time point (Safety Population) | |
| Table 14.2.1.3.3 | Statistical Analysis of Immunoglobulin A by Time point (mITT Population) | |
| Table 14.2.1.4.1 | Statistical Analysis of Anti-COVID-19 Neutralization Titer by Time point (PP Population) | X |
| Table 14.2.1.4.2 | Statistical Analysis of Anti-COVID-19 Neutralization Titer by Time point (Safety Population) | X |
| Table 14.2.1.4.3 | Statistical Analysis of Anti-COVID-19 Neutralization Titer by Time point (mITT Population) | X |
| Table 14.2.1.5.1 | Statistical Analysis of CD4+ T-cells Values by Time point (PP Population) | |
| Table 14.2.1.5.2 | Statistical Analysis of CD4+ T-cells Values by Time point (Safety Population) | |
| Table 14.2.1.5.3 | Statistical Analysis of CD4+ T-cells Values by Time point (mITT Population) | |
| Table 14.2.1.6.1 | Statistical Analysis of CD8+ T-cells Values by Time point (PP Population) | |

DocUUID : 37b099b4-2512-4dbf-8238-d258ccb127ba

| | | |
|---|---|---|
| Table 14.2.1.6.2 | Statistical Analysis of CD8+ T-cells Values by Time point (Safety Population) | |
| Table 14.2.1.6.3 | Statistical Analysis of CD8+ T-cells Values by Time point (mITT Population) | |
| Table 14.2.1.7.1 | Statistical Analysis of Peripheral Blood Mononuclear Cell Results by Time point (PP Population) | |
| Table 14.2.1.7.2 | Statistical Analysis of Peripheral Blood Mononuclear Cell Results by Time point (Safety Population) | |
| Table 14.2.1.7.3 | Statistical Analysis of Peripheral Blood Mononuclear Cell Results by Time point (mITT Population) | |
| Table 14.2.1.8.1 | Statistical Analysis of Flow Cytometry Results by Time point (PP Population) | |
| Table 14.2.1.8.2 | Statistical Analysis of Flow Cytometry Results by Time point (Safety Population) | |
| Table 14.2.1.8.3 | Statistical Analysis of Flow Cytometry Results by Time point (mITT Population) | |
| Table 14.2.2.1.1 | Summary of Anti-COVID-19 RBD Immunoglobulin G by Time point (PP Population) | X |
| Table 14.2.2.1.2 | Summary of Anti-COVID-19 RBD Immunoglobulin G by Time point (Safety Population) | X |
| Table 14.2.2.1.3 | Summary of Anti-COVID-19 RBD Immunoglobulin G by Time point (mITT Population) | X |
| Table 14.2.2.2.1 | Summary of Total Anti S-Protein Immunoglobulin G by Time point (PP Population) | X |
| Table 14.2.2.2.2 | Summary of Total Anti S-Protein Immunoglobulin G by Time point (Safety Population) | X |
| Table 14.2.2.2.3 | Summary of Total Anti S-Protein Immunoglobulin G by Time point (mITT Population) | X |
| Table 14.2.2.3.1 | Summary of Immunoglobulin A by Time point (PP Population) | |
| Table 14.2.2.3.2 | Summary of Immunoglobulin A by Time point (Safety Population) | |
| Table 14.2.2.3.3 | Summary of Immunoglobulin A by Time point (mITT Population) | |
| Table 14.2.2.4.1 | Summary of Anti-COVID-19 Neutralization Titer by Time point (PP Population) | X |
| Table 14.2.2.4.2 | Summary of Anti-COVID-19 Neutralization Titer by Time point (Safety Population) | X |
| Table 14.2.2.4.3 | Summary of Anti-COVID-19 Neutralization Titer by Time point (mITT Population) | X |
| Table 14.2.2.5.1 | Summary of CD4+ T-cells Values by Time point (PP Population) | |

DocUUID : 37b099b4-2512-4dbf-8238-d258ccb127ba

| | |
|---|---|
| Table 14.2.2.5.2 | Summary of CD4+ T-cells Values by Time point (Safety Population) |
| Table 14.2.2.5.3 | Summary of CD4+ T-cells Values by Time point (mITT Population) |
| Table 14.2.2.6.1 | Summary of CD8+ T-cells Values by Time point (PP Population) |
| Table 14.2.2.6.2 | Summary of CD8+ T-cells Values by Time point (Safety Population) |
| Table 14.2.2.6.3 | Summary of CD8+ T-cells Values by Time point (mITT Population) |
| Table 14.2.2.7.1 | Summary of Peripheral Blood Mononuclear Cell Results by Time point (PP Population) |
| Table 14.2.2.7.2 | Summary of Peripheral Blood Mononuclear Cell Results by Time point (Safety Population) |
| Table 14.2.2.7.3 | Summary of Peripheral Blood Mononuclear Cell Results by Time point (mITT Population) |
| Table 14.2.2.8.1 | Summary of Flow Cytometry Results by Time point (PP Population) |
| Table 14.2.2.8.2 | Summary of Flow Cytometry Results by Time point (Safety Population) |
| Table 14.2.2.8.3 | Summary of Flow Cytometry Results by Time point (mITT Population) |
| Figure 14.2.1.1 | Plot of Mean (SD) Immunogenicity Variables (PP Set)<br>note: linear and semi-log |
| Figure 14.2.1.2 | Plot of Mean (SD) Immunogenicity Variables (Safety Set)<br>note: linear and semi-log |
| Figure 14.2.1.3 | Plot of Mean (SD) Immunogenicity Variables (mITT Set)<br>note: linear and semi-log |
| **Section 14.3** | **Safety Data Summaries** |
| **Section 14.3.1** | **Displays of Adverse Events** |
| Table 14.3.1.1.1 | Overall Summary of Treatment-Emergent Adverse Events (Safety Population) |
| Table 14.3.1.1.2 | Overall Summary of Treatment-Emergent Adverse Events (PP Population) |
| Table 14.3.1.2.1 | Summary of Treatment-Emergent Adverse Events by System Organ Class and Preferred Term (Safety Population) |
| Table 14.3.1.2.2 | Summary of Treatment-Emergent Adverse Events by System Organ Class and Preferred Term (PP Population) |
| Table 14.3.1.3.1 | Summary of Treatment-Emergent Adverse Events by Severity by System Organ Class and Preferred Term (Safety Population) |

DocUUID : 37b099b4-2512-4dbf-8238-d258ccb127ba

| | |
|---|---|
| Table 14.3.1.3.2 | Summary of Treatment-Emergent Adverse Events by Severity by System Organ Class and Preferred Term (PP Population) |
| Table 14.3.1.4.1 | Summary of Medically Attended Adverse Events by Severity, System Organ Class and Preferred Term (Safety Population) |
| Table 14.3.1.4.2 | Summary of Medically Attended Adverse Events by Severity, System Organ Class and Preferred Term (PP Population) |
| Table 14.3.1.5.1 | Summary of Adverse Event of Special Interest by Severity, System Organ Class and Preferred Term (Safety Population) |
| Table 14.3.1.5.2 | Summary of Adverse Event of Special Interest by Severity, System Organ Class and Preferred Term (PP Population) |
| Table 14.3.1.6.1 | Summary of Treatment-Emergent Adverse Events by Relationship, System Organ Class and Preferred Term (Safety Population) |
| Table 14.3.1.6.2 | Summary of Treatment-Emergent Adverse Events by Relationship, System Organ Class and Preferred Term (PP Population) |
| Table 14.3.1.7.1 | Summary of Medically Attended Adverse Events by Relationship, System Organ Class and Preferred Term (Safety Population) |
| Table 14.3.1.7.2 | Summary of Medically Attended Adverse Events by Relationship, System Organ Class and Preferred Term (PP Population) |
| Table 14.3.1.8.1 | Summary of Adverse Event of Special Interest by Relationship, System Organ Class and Preferred Term (Safety Population) |
| Table 14.3.1.8.2 | Summary of Adverse Event of Special Interest by Relationship, System Organ Class and Preferred Term (PP Population) |
| Table 14.3.1.9.1 | Summary of Serious Adverse Events by System Organ Class and Preferred Term (Safety Population) |
| Table 14.3.1.9.2 | Summary of Serious Adverse Events by System Organ Class and Preferred Term (PP Population) |
| Table 14.3.1.10.1 | Summary of New Onset Chronic Disease by System Organ Class and Preferred Term (Safety Population) |
| Table 14.3.1.10.2 | Summary of New Onset Chronic Disease by System Organ Class and Preferred Term (PP Population) |
| Table 14.3.1.11.1 | Summary of Potential Immune-Mediated Medical Condition by System Organ Class and Preferred Term (Safety Population) |
| Table 14.3.1.11.2 | Summary of Potential Immune-Mediated Medical Condition by System Organ Class and Preferred Term (PP Population) |

DocUUID : 37b099b4-2512-4dbf-8238-d258ccb127ba

| | |
|---|---|
| Table 14.3.3.1 | Summary of Solicited Local and Systemic Adverse Site Reactions within 7 days after Each Dose (Safety Population) |
| Table 14.3.3.2 | Summary of Solicited Local and Systemic Adverse Site Reactions within 7 days after Each Dose by Day of Immunization and Severity (Safety Population) |
| **Section 14.3.2** | **Listings of Deaths, Other Serious and Certain Significant Adverse Events** |
| Table 14.3.2.1 | Listing of Adverse Events with Outcome of Death (Safety Population) |
| Table 14.3.2.2 | Listing of Serious Adverse Events (Safety Population) |
| Table 14.3.2.3 | Listing of Adverse Events Leading to Study or Study Drug Discontinuation (Safety Population) |
| **Section 14.3.4** | **Abnormal Laboratory Values** |
| Table 14.3.4 | Listing of Abnormal Laboratory Results (Safety Population) |
| **Section 14.3.5** | **Additional Safety Data Summaries** |
| Table 14.3.5.1 | Observed and Change from Baseline for Clinical Laboratory Parameters (Safety Population) |
| Table 14.3.5.2 | Observed and Change from Baseline for Clinical Laboratory Parameters (PP Population) |
| Table 14.3.5.3 | Shift Assessment of Categorical Change of Laboratory Results from Baseline to each Post-Baseline Visit (Safety Population) |
| Table 14.3.5.4 | Shift Assessment of Categorical Change of Laboratory Results from Baseline to each Post-Baseline Visit (PP Population) |
| Table 14.3.5.5 | Observed and Change from Baseline Vital Signs (Safety Population) |
| Table 14.3.5.6 | Observed and Change from Baseline Vital Signs (PP Population) |
| **Listing Number** | **Listing Name** |
| **Section 16.2.1** | **Discontinued Subjects** |
| Listing 16.2.1 | Subject Disposition (Randomized Population)<br>Note: Including consent information |
| **Section 16.2.2** | **Protocol Deviations** |
| Listing 16.2.2 | Protocol Deviations (Safety Population) |
| **Section 16.2.3** | **Subjects Excluded from Analysis** |
| Listing 16.2.3 | Analysis Populations |
| **Section 16.2.4** | **Demographic Data** |
| Listing 16.2.4.1 | Demographics Characteristics (Safety Population)<br>Note: include height, weight and BMI |
| Listing 16.2.4.2 | Eligibility Criteria Satisfaction (Safety Population) |
| Listing 16.2.4.3 | Medical and Surgical History (Safety Population) |

DocUUID : 37b099b4-2512-4dbf-8238-d258ccb127ba

| | | |
|---|---|---|
| Listing 16.2.4.4 | Prior and Concomitant Medications (Safety Population) | |
| Listing 16.2.4.5 | Concomitant Procedures (Safety Population) | |
| **Section 16.2.5** | **Compliance Data** | |
| Listing 16.2.5.1 | Study Drug Dosing (Safety Population) | |
| Listing 16.2.5.2 | Eligibility for Vaccination (Safety Population) | |
| **Section 16.2.6** | **Individual Immunogenicity Response Data** | |
| Listing 16.2.6 | Individual Titer Values (Safety Population)<br>Note: Anti-COVID-19 Ig, IgG, IgA, anti-COVID-19 neutralization titer assays, PBMC, Flow Cytometry results | X |
| Figure 16.2.6.1 | Individual Plot of Immunogenicity Variables (PP Set)<br>note: linear and semi-log | |
| Figure 16.2.6.2 | Individual Plot of Immunogenicity Variables (Safety Set)<br>note: linear and semi-log | |
| Figure 16.2.6.3 | Individual Plot of Immunogenicity Variables (mITT Set)<br>note: linear and semi-log | |
| **Section 16.2.7** | **Adverse Event Listings** | |
| Listing 16.2.7.1 | Adverse Events (Safety Population) | |
| Listing 16.2.7.2.1 | Reactogenicity Diary Card - Generalized Symptoms (Safety Population) | |
| Listing 16.2.7.2.2 | Reactogenicity Diary Card - Local Reactions (Safety Population) | |
| Listing 16.2.7.3 | Vaccine Reaction and Adverse Event Diary (Safety Population) | |
| **Section 16.2.8** | **Individual Laboratory Measurements by Subject** | |
| Listing 16.2.8.1 | Normal Ranges for Laboratory Data | |
| Listing 16.2.8.2 | Clinical Laboratory Data by Category (Safety Population)<br>Note: except serology and pregnancy test | |
| Listing 16.2.8.3 | Urine Drug Screen Results (Safety Population) | |
| Listing 16.2.8.4 | Alcohol Breath Test Results (Safety Population) | |
| Listing 16.2.8.5 | Serology Test Results (Safety Population) | |
| Listing 16.2.8.6 | Pregnancy Test Results (Safety Population) | |
| Listing 16.2.8.7 | Post-Vaccination Assessment (Safety Population) | |
| Listing 16.2.8.8 | Vital Signs (Safety Population) | |
| Listing 16.2.8.9 | Abnormal Physical Examination Findings (Safety Population) | |
| Listing 16.2.8.10 | Nasopharyngeal Swab for RT-PCR Results (Safety Population) | |
| Listing 16.2.8.11 | Telephone Contact (Safety Population) | |
| Listing 16.2.8.12 | Comments (Safety Population) | |

DocUUID : 37b099b4-2512-4dbf-8238-d258ccb127ba
